# Vaccine hesitancy for COVID-19 explored in a phenomic study of 259 socio-cognitive-behavioural measures in the UK-REACH study of 12,431 UK healthcare workers

**DOI:** 10.1101/2021.12.08.21267421

**Authors:** I Chris McManus, Katherine Woolf, Christopher A Martin, Laura B Nellums, Anna L Guyatt, Carl Melbourne, Luke Bryant, Amit Gupta, Catherine John, Martin D Tobin, Sue Carr, Sandra Simpson, Bindu Gregary, Avinash Aujayeb, Stephen Zingwe, Rubina Reza, Laura J Gray, Kamlesh Khunti, Manish Pareek, On behalf of the UK-REACH Study Collaborative Group

## Abstract

**Background:** Vaccination is key to successful prevention of COVID-19 particularly nosocomial acquired infection in health care workers (HCWs). ‘Vaccine hesitancy’ is common in the population and in HCWs, and like COVID-19 itself, hesitancy is more frequent in ethnic minority groups. UK-REACH (United Kingdom Research study into Ethnicity and COVID-19 outcomes) is a large-scale study of COVID-19 in UK HCWs from diverse ethnic backgrounds, which includes measures of vaccine hesitancy. The present study explores predictors of vaccine hesitancy using a ‘phenomic approach’, considering several hundred questionnaire-based measures.

**Methods:** UK-REACH includes a questionnaire study encompassing 12,431 HCWs who were recruited from December 2020 to March 2021 and completed a lengthy online questionnaire (785 raw items; 392 derived measures; 260 final measures). Ethnicity was classified using the Office for National Statistics’ five (ONS5) and eighteen (ONS18) categories. Missing data were handled by multiple imputation. Variable selection used the *islasso* package in *R*, which provides standard errors so that results from imputations could be combined using Rubin’s rules. The data were modelled using path analysis, so that predictors, and predictors of predictors could be assessed. Significance testing used the Bayesian approach of Kass and Raftery, a ‘very strong’ Bayes Factor of 150, N=12,431, and a Bonferroni correction giving a criterion of p<4.02 × 10^−8^ for the main regression, and p<3.11 × 10^−10^ for variables in the path analysis.

**Results:** At the first step of the phenomic analysis, six variables were direct predictors of greater vaccine hesitancy: Lower pro-vaccination attitudes; no flu vaccination in 2019-20; pregnancy; higher COVID-19 conspiracy beliefs; younger age; and lower optimism the roll-out of population vaccination. Overall 44 lower variables in total were direct or indirect predictors of hesitancy, with the remaining 215 variables in the phenomic analysis not independently predicting vaccine hesitancy. Key variables for predicting hesitancy were belief in conspiracy theories of COVID-19 infection, and a low belief in vaccines in general. Conspiracy beliefs had two main sets of influences:

i. Higher Fatalism, which was influenced a) by high external and chance locus of control and higher need for closure, which in turn were associated with neuroticism, conscientiousness, extraversion and agreeableness; and b) by religion being important in everyday life, and being Muslim.
ii. receiving information via social media, not having higher education, and perceiving greater risks to self, the latter being influenced by higher concerns about spreading COVID, greater exposure to COVID-19, and financial concerns.

There were indirect effects of ethnicity, mediated by religion. Religion was more important for Pakistani and African HCWs, and less important for White and Chinese groups. Lower age had a direct effect on hesitancy, and age and female sex also had several indirect effects on hesitancy.

**Conclusions:** The phenomic approach, coupled with a path analysis revealed a complex network of social, cognitive, and behavioural influences on SARS-Cov-2 vaccine hesitancy from 44 measures, 6 direct and 38 indirect, with the remaining 215 measures not having direct or indirect effects on hesitancy. It is likely that issues of trust underpin many associations with hesitancy. Understanding such a network of influences may help in tailoring interventions to address vaccine concerns and facilitate uptake in more hesistant groups.

**Funding:** UKMRI-MRC and NIHR

## Introduction

Successful prevention of COVID-19 requires not only effective vaccines and effective vaccination programmes against SARS-Cov-2 (^a^: see footnote a),, but also, “it needs people who believe in [vaccines]”^1^. As Stephen Reicher, a member of the UK Government’s Independent Scientific Pandemic Insights Group on Behaviours (SPI-B) put it:

> “A vaccine solves nothing. It is people getting vaccinated that will affect the disease and its transmission. So, we need to address issues of vaccine hesitancy, why hesitancy is so much greater in some groups than others, how conspiracy theories gain traction, and how to impact all of these if the vaccine is to play its part.” ^2^

Relatively little is known about the social, cognitive and behavioural underpinnings of COVID-19 vaccine hesitancy in the population ^3^. Even less is known about hesitancy in healthcare workers (HCWs). A simple expectation might be that vaccine hesitancy should be rare, given HCWs’ professional and scientific training, and their experiences of patient care in the COVID-19 pandemic^4^. However, in our previous, interim, analysis of data from the United Kingdom Research study into Ethnicity and COVID-19 outcomes in Healthcare workers (UK-REACH)^5^, 23% of HCWs were COVID-19 vaccine hesitant, including 3% who were vaccine refusing. COVID-19 vaccine hesitancy was more common among younger and HCWs, and Black Caribbean (54.2%), Mixed White and Black Caribbean (38.1%), Black African (34.4%), Chinese (33.1%), Pakistani (30.4%), and White Other (28.7%) ethnic groups ^5^. Hesitancy was less likely in those with pro-vaccine attitudes in general ^6^ and more common in those with COVID-19 conspiracy beliefs ^7 5^. Our previous, interim, analysis did not further explore predictors of hesitancy, or the predictors of those predictors. A qualitative study of hesitancy in UK-REACH of 164 HCWs as a part of UK-REACH identified a number of influences, noting particularly, “(mis)trust, including historical (mis)trust based on experiences of racism and discrimination … [as] important factor[s] in determining vaccine attitudes” ^8^.

The present analysis uses a *phenomic approach* to look at the association of a wide range of measures with vaccine hesitancy in the HCWs in UK-REACH, with the aim of informing and developing interventions for reducing hesitancy and increasing vaccine uptake, particularly with the advent in Autumn 2021 in the UK of booster doses for COVID-19 as well as influenza vaccination for those most at risk, including HCWs. In November 2021 NHS England also announced that COVID-19 vaccination would become mandatory for all NHS staff in England.

### Phenomics

Lewontin described the phenome as “the actual physical manifestation of the organism including its morphology, physiology, *and behavior*”[our emphasis] ^9^ and it is now used in a variety of contexts, such as the Human Phenome Project ^10^, and Phenome-wide Association Scans (PheWAS) ^11^, sometimes in a narrower sense of phenotypes related to genotypes ^12^, and also in broad terms ^13^. The term phenomics was first used in 1996 by Steve Garan ^14^. A clear example of phenomics, albeit not called as such, but which inspired the methodology of the present study, used UK Biobank data to assess five-year mortality in relation to 655 varied measures of demographics, health and lifestyle ^15^. We will refer here to the ‘socio-cognitive-behavioural’ phenome since most of our measures are questionnaire-based, are broadly behavioural in type, and at individual rather than group level^16^

### Hesitancy, HCWs and UK-REACH

HCWs have been prioritised within many vaccination programmes globally, but several studies have found evidence of hesitancy to COVID-19 vaccines even within HCW populations ^5 17–19^. Understanding and reducing hesitancy among HCWs is of particular importance because of increased impact of COVID-19 on HCW health, increased risk of nosocomial transmission ^20 21^ and influence of HCW vaccine attitudes on patient and population vaccine uptake ^22^. There is also particular concern about greater vaccine hesitancy and lower vaccine uptake among HCWs from ethnic minority groups ^23 24^. Individuals from ethnic minority backgrounds are at greater risk from COVID-19 ^25 26 27^ and UK National Health Service (NHS) staff are more ethnically diverse than the general population^28^. UK-REACH collected data from December 2020 to March 2021, at the peak of the UK’s second wave of the COVID-19 pandemic, and at the start of the UK vaccination programme. UK-REACH deliberately over-sampled HCWs from ethnic minority backgrounds, and used a wide-ranging questionnaire to assess the social-cognitive-behavioural *phenome* of COVID-19 vaccine hesitancy in HCWs.

### Defining Vaccine Hesitancy

We used the World Health Organisation (WHO) of vaccine hesitancy as refusal or delay in acceptance of vaccine(s) ^29^. Individuals vary from complete refusal of all vaccines to refusal or delay in uptake of a particular vaccine. Low vaccine uptake can also result from systemic failures preventing access to vaccines, including shortages or poorly located vaccine clinics. This study focuses on vaccine hesitancy, aiming to describe and understand the behavioural, attitudinal, social, cultural, and work-related factors that contribute to hesitancy in accepting COVID-19 vaccines in HCWs in the UK, especially among those from ethnic minority groups.

The WHO Working Group on Vaccine Hesitancy provides two main frameworks for understanding and addressing vaccine hesitancy (WHO, 2014). The “Three C’s” model has three broad dimensions that influence hesitancy: **confidence** (trust in safety and efficacy of vaccines, in reliability and competence of health services and HCWs, and in policy-makers deciding on the needed vaccines); **complacency** (perceived risks to self and society of the disease being vaccinated against); and **convenience** (ease of access and perceived quality of vaccination services) ^22 30–32^.

The WHO’s more complex Hesitancy Determinants Matrix considers **contextual influences** (the social, historic, systemic, political and economic contexts of a vaccination programme, including the communication and media environment), **individual and group influences** (including family/friend/community experiences of vaccination and social norms around vaccination), **and vaccine/vaccination-specific issues** (including the mode of vaccination administration and the ways in which the behaviours of healthcare professionals influence hesitancy). In addition, good communication and information provision are regarded as key to increased acceptance and reduced hesitancy.

## Methods

### Overview

We consider baseline questionnaire data from the consented, longitudinal cohort (work package 2) of UK-REACH ^33^, which was administered online from 4^th^ December 2020 to 8^th^ March 2021. Our previous, interim, analysis ^5^ had utilised interim data downloaded 19^th^ February 2021. We summarise the methods below, with further details given in the Supplementary Information, in the study protocol ^33^ and in the interim analysis ^5^.

### Study population and sampling

Any HCWs, including clinical and ancillary workers, in a UK healthcare setting aged 16 or over, including those registered with one of seven main healthcare regulatory bodies were eligible to participate. The records and registration information of the regulators were used as a sampling frame. See Supplementary Information for further details ^b^.

### Recruitment and data collection

Regulatory bodies sent email invitations and two reminders to 1,052,875 HCWs, providing a link to the study registration page. In addition, 21 NHS Hospital Trusts publicised the questionnaire to their staff and invited staff by email, and open links to the questionnaire were also advertised on social media and in newsletters. After registering, participants accessed an online consent form and the online questionnaire administered via REDCap ^34^.

### Primary outcome measure

The primary outcome measure was COVID-19 vaccine hesitancy, described in Supplementary Figure 1^c^, from the previous, interim, analysis ^5^. The outcome measure comprises a combination of two versions of the vaccine hesitancy question. The first vaccination question (VQ1) was administered between 4^th^ and 20^th^ Dec 2020 and asked “When a vaccine becomes available, would you wish to receive it?” To reflect the rapid roll-out of the vaccines among UK HCWs, VQ1 was replaced on 21^st^ December 2020 with VQ2, which asked: “Have you had, or are you going to have, a vaccination against COVID-19?”, with branching logic for the possible responses. Statistical models included a binary dummy variable, ‘hesitancy question type’, to take account of the different question sets, VQ1 and VQ2.

### Predictors

The questionnaire included 780 raw items on a wide range of social, cognitive and behavioural questions: work and job roles and attitudes to work; exposure to COVID-19; ethnicity, culture and religion; education; home environment; physical and mental health; specific experience with COVID-19 and national lockdowns; and individual difference measures including attitudes, values and personality (see Table 1). Many items were parts of logic chains, and were not encountered by all participants, and others consisted of raw items on multi-item scales. Some measures were derived from raw items, with some derived variables including missing variable indicators. Participants could skip questions or select “prefer not to answer” (PNTA).

**Table 1:**
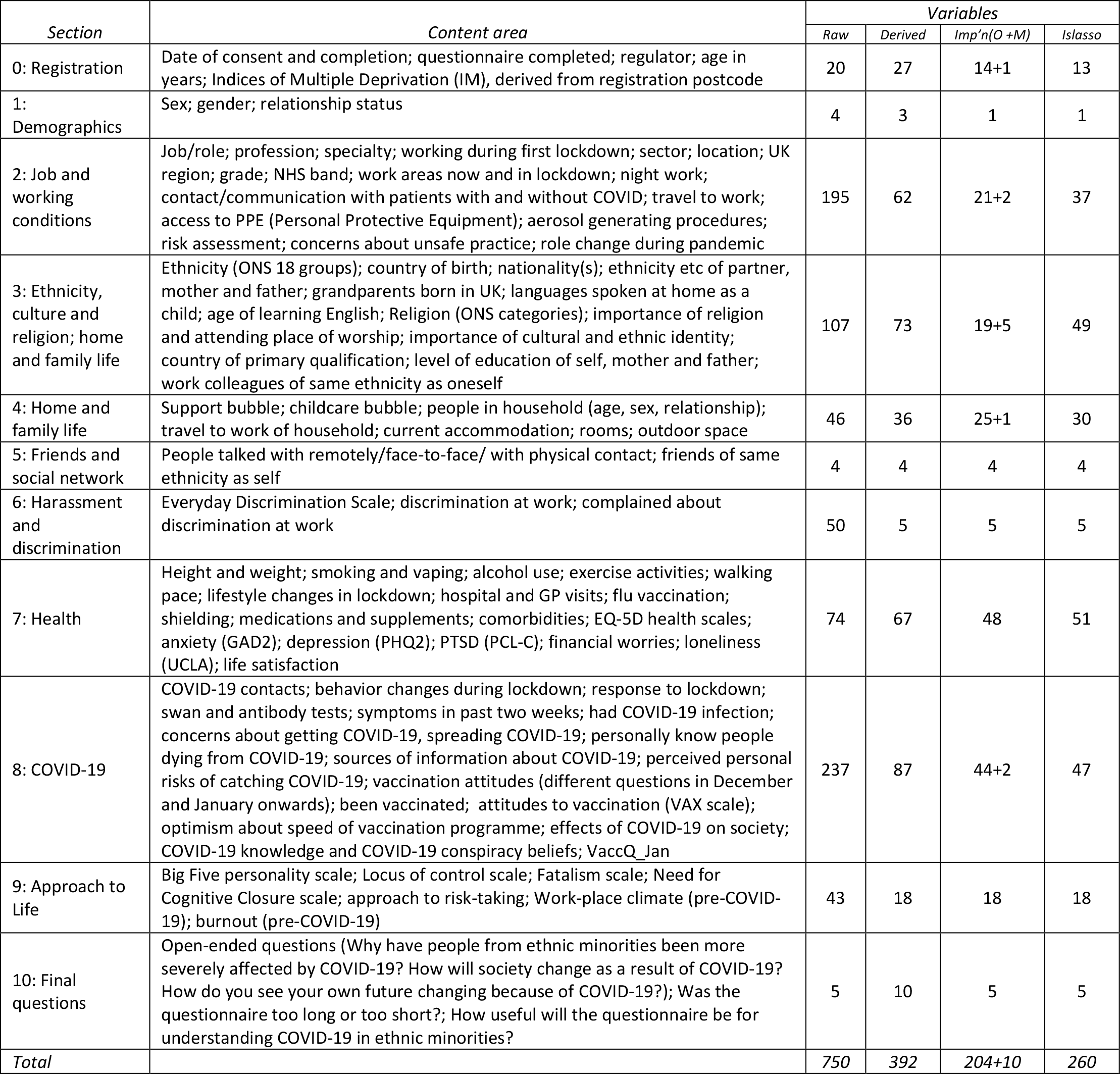
Summary of question and measures content in the eleven questionnaire sections. For further details see the Data Dictionary^m^. Raw items refer to individual variables in *RedCap*, and in many cases involve logic chains so that not all participants see all questions, or have many raw items due to the multiple checkbox format. Derived variables sometimes have the same information in different formats (e.g. age in years, age in decadal groups, etc). Imputation variables are mostly binary, continuous, ordinal or multinomial (shown as O+M = ordinal+multinomial), with collinear variables omitted. Variables entered into *islasso* include multinomials post-processed after imputation into binary variables, with the multinomial variables themselves subsequently omitted. See Supplementary Information for further details.

| Section | Content area | Variables |  |  |  |
| --- | --- | --- | --- | --- | --- |
|  |  | Raw | Derived | Imp'n(O +M) | Islasso |
| 0: Registration | Date of consent and completion; questionnaire completed; regulator; age in years; Indices of Multiple Deprivation (IM), derived from registration postcode | 20 | 27 | 14+1 | 13 |
| 1: Demographics | Sex; gender; relationship status | 4 | 3 | 1 | 1 |
| 2: Job and working conditions | Job/role; profession; specialty; working during first lockdown; sector; location; UK region; grade; NHS band; work areas now and in lockdown; night work; contact/communication with patients with and without COVID; travel to work; access to PPE (Personal Protective Equipment); aerosol generating procedures; risk assessment; concerns about unsafe practice; role change during pandemic | 195 | 62 | 21+2 | 37 |
| 3: Ethnicity, culture and religion; home and family life | Ethnicity (ONS 18 groups); country of birth; nationality(s); ethnicity etc of partner, mother and father; grandparents born in UK; languages spoken at home as a child; age of learning English; Religion (ONS categories); importance of religion and attending place of worship; importance of cultural and ethnic identity; country of primary qualification; level of education of self, mother and father; work colleagues of same ethnicity as oneself | 107 | 73 | 19+5 | 49 |
| 4: Home and family life | Support bubble; childcare bubble; people in household (age, sex, relationship); travel to work of household; current accommodation; rooms; outdoor space | 46 | 36 | 25+1 | 30 |
| 5: Friends and social network | People talked with remotely/face-to-face/ with physical contact; friends of same ethnicity as self | 4 | 4 | 4 | 4 |
| 6: Harassment and discrimination | Everyday Discrimination Scale; discrimination at work; complained about discrimination at work | 50 | 5 | 5 | 5 |
| 7: Health | Height and weight; smoking and vaping; alcohol use; exercise activities; walking pace; lifestyle changes in lockdown; hospital and GP visits; flu vaccination; shielding; medications and supplements; comorbidities; EQ-5D health scales; anxiety (GAD2); depression (PHQ2); PTSD (PCL-C); financial worries; loneliness (UCLA); life satisfaction | 74 | 67 | 48 | 51 |
| 8: COVID-19 | COVID-19 contacts; behavior changes during lockdown; response to lockdown; swan and antibody tests; symptoms in past two weeks; had COVID-19 infection; concerns about getting COVID-19, spreading COVID-19; personally know people dying from COVID-19; sources of information about COVID-19; perceived personal risks of catching COVID-19; vaccination attitudes (different questions in December and January onwards); been vaccinated; attitudes to vaccination (VAX scale); optimism about speed of vaccination programme; effects of COVID-19 on society; COVID-19 knowledge and COVID-19 conspiracy beliefs; VaccQ_Jan | 237 | 87 | 44+2 | 47 |
| 9: Approach to Life | Big Five personality scale; Locus of control scale; Fatalism scale; Need for Cognitive Closure scale; approach to risk-taking; Work-place climate (pre-COVID-19); burnout (pre-COVID-19) | 43 | 18 | 18 | 18 |
| 10: Final questions | Open-ended questions (Why have people from ethnic minorities been more severely affected by COVID-19? How will society change as a result of COVID-19? How do you see your own future changing because of COVID-19?); Was the questionnaire too long or too short?; How useful will the questionnaire be for understanding COVID-19 in ethnic minorities? | 5 | 10 | 5 | 5 |
| Total |  | 750 | 392 | 204+10 | 260 |

<sup>m</sup> https://uniofleicester-my.sharepoint.com/:x:/r/personal/mp426_leicester_ac_uk/_layouts/15/Doc.aspx?sourcedoc=%7B79688523-78DA-4968-8CA4-29DD01240921%7D&file=UK-REACH_questionnaire_data_dictionary_v1.0.xlsx&action=default&mobileredirect=true

The following key variables are briefly described here, with more extensive descriptions in the Supplementary Information^d^: ethnicity, religion, pro-vaccine attitudes, COVID-19 conspiracy beliefs, influenza vaccine uptake, optimism about the COVID-19 vaccine roll-out, perceived risks of COVID-19 to self, fatalism, locus of control, and need for cognitive closure. Further details are also found in the study protocol ^33^, and in the UK-REACH online data dictionary (https://www.uk-reach.org/data-dictionary).

#### Ethnicity: ONS18 and ONS5 category variables

Participants self-identified their ethnic group using the 18 Office for National Statistics categories used in the 2011 Census (‘ONS18’) ^35^. We also used the five main ethnic sub-categories of the Census (Black, Asian, Mixed, Other, White; ‘ONS5’). In our choice of terms we have followed the BMJ’s special edition on Racism in Medicine and used the term “ethnic minority” ^36 37^. Too inclusive ethnic groupings can mask important ethnic and cultural differences, but too fine a categorisation can lose statistical power.

#### Religion and religiosity

Religion was self-described using the UK Census 2011 question, with seven categories plus a freetext “Other” option. Participants were also asked on a four-point scale how important religion was to them in their everyday life.

#### Pro-vaccine attitudes

Four items of the Vaccination Attitudes Examination (VAX) Scale ^38^, one from each subscale, were summed, with high scores indicating pro-vaccination attitudes.

#### COVID-19 conspiracy beliefs

Sum of six scores on the COVID-19 conspiracy belief scale^7^, each rated on a 4-point scale with high scores indicating high COVID-19 conspiracy beliefs.

#### Optimism about vaccination programme

A single item asking how soon it was thought it would be possible to vaccinate most of the population against coronavirus, with greater optimism indicated by shorter times.

#### Influenza vaccination winter 2019/2020

A single Yes/No item asking whether the participant had received a flu vaccine in the winter of 2019-2020.

#### Perceived risk of COVID-19 to self

Mean response to two questions on participant’s rating of their personal chances of catching the coronavirus in the next six months and b) needing hospital treatment If they did catch coronavirus. High scores indicated higher perceived risk.

#### Fatalism

Sum of four scores on 7-point items from the Fatalism Scale^39^. Higher scores indicate higher fatalism (fate determining outcomes).

#### Locus of control

Sum of three items on 7-point scales for each of the three subscales of a version of Levenson’s Locus of Control (LoC) scale ^40^. Internal LoC indicates that one’s own actions are perceived to determine outcomes; chance LoC that random external events determine outcomes; and external LoC that powerful others determine outcomes.

#### Need for cognitive closure

Sum on two items on a 7-point scale from the short version of the Need for Cognitive Closure scale ^41^. Higher scores indicate higher need for cognitive closure and lower tolerance of ambiguity.

#### Personality

Sum of three items, each on 7-point scales, from each of the five sub-scales of the Big Five personality inventory, with traits of Neuroticism, Extraversion, Openness to Experience, Agreeableness and Conscientiousness, as used in *Understanding Society* ^42^.

### Statistical analysis

Statistical analyses were performed in *SPSS* and *R* ^43^. Further details of all analyses are provided in the Supplementary Information^e^.

#### Overall approach to the statistical analysis

Our key aim was to carry out a path analysis in which firstly predictors of vaccine hesitancy were identified from the large set of behavioural variables, the phenome. Then in turn predictors were found for those predictors, and then for predictors of the predictors, etc.. Each step required a variable selection, for which the least absolute shrinkage and selection operator (LASSO) is now a recognized approach ^44^. Missing data however had to be accounted for, and therefore the analyses took place separately on each of a set of 20 multiply imputed datasets. Combining analyses of separate imputed datasets usually combines estimates and standard errors using Rubin’s Rules ^45^. However, LASSO does not provide standard errors of estimates, particularly for variables subject to shrinkage, meaning that Rubin’s Rules cannot be applied. A solution is to use the *islasso* package in R ^46 47^which does provide standard errors, and then results can be combined in the usual way. A further challenge is that a statistical criterion for significance is required for the combined variables, and for that the Bayesian approach of Kass and Raftery^48 49^ was used, coupled with a Bonferroni correction. The separate steps are described briefly below and in detail in the Supplementary Statistical Information.

#### Missing data and multiple imputation

For the 12,431 participants who signed off and submitted their results, 4.6% of data points were missing (121578/2660234), with a median of 3.3% missing data points per participant. Pro-vaccine attitudes data (VAX) were structurally missing for participants who answered VQ1 from 4^th^ to 20^th^ December 2020. Missing data were multiply imputed for 20 imputation sets with predictive mean matching ^50^ using the package *mice*. Details of variables used in the imputation are provided in Table 1 and in the Supplementary Information ^*f*^. Estimates and standard errors were combined across imputation sets using Rubin’s Rules ^45^.

#### Univariate analyses

Correlations between hesitancy and predictor variables (partialling out hesitancy question type) were calculated using the *micombine*.*cor()* function in the *miceadds* package of R.

#### Selection of variables

The Induced Smooth (IS) lasso method in the *islasso* package ^46 47^ was used to select variables for inclusion in the path model. The hesitancy outcome measure was used as the dependent variable in relation to the set of predictor variables, each of which could either remain in the analysis or its regression coefficient could shrink towards zero. The exception was the dummy vaccination question variable, which was unpenalized and remained in the model. *islasso* requires that all variables are standardized to a mean of zero and standard deviation of one, so that all coefficients are directly comparable.

#### Path modelling

The predictors of hesitancy were modelled piecemeal, as a series of separate multiple regressions ^51–53^ using *islasso*, with previous predictors which had reached significance being set in turn as dependent variables. To avoid problems of reciprocal causation, dependent variables were restricted to those that were not obviously causally posterior and were not previously in the analysis ^54^. Demographic variables were regarded as exogenous and not modelled further.

#### Statistical significance levels

Using the Bayesian approach of Raftery and Kass ^48 49^, a Bayes Factor of 150 (“Very strong”) and N of 12,431 has an equivalent p value of 1.04 × 10^−5^. Bonferroni corrections for 259 tests for the primary analysis (and simple correlations) gave a threshold of p= 4.02 × 10^−8^, and 259×258/2 tests for the path models gave a threshold of p=3.11 × 10^−10^ ^g^. These significance levels lead to very similar results to those found by the different approach of the *glasso* program for Gaussian Graphical Modelling (GGM)^h^.

### Ethical approval

The study was approved by the Health Research Authority (Brighton and Sussex Research Ethics Committee; ethics reference: 20/HRA/4718). All participants gave written informed consent. The study was registered with ISRCTN on 27/11/2020 (https://doi.org/10.1186/ISRCTN11811602).

### Involvement and engagement

The UK-REACH study has regular input from a Professional Expert Panel of HCWs from a range of ethnic backgrounds, occupations, and genders, as well as with national and local organisations, described in the study protocols ^33 55^.

## Results

### Participants

Overall 12,431/15,592 (78%) of registered participants who completed the questionnaire between 4^th^ Dec 2020 and 8^th^ March 2021 were included in the analysis with missing data for 4.6%. Participants who registered but did not complete were missing 75.5% of data. Supplementary Table 1^i^, which shows the demographic characteristics of the cohort stratified by COVID-19 vaccine hesitancy, is an updated version of Table 2 in our previous, interim analysis ^5^, but with the somewhat larger final sample.

### Simple correlations with vaccine hesitancy

Using a slight modification of the approach of Ganna and Ingelsson^15^, correlations were calculated between each phenomic measure and vaccine hesitancy, after partialling out questionnaire variant. Age inevitably correlates with very many measures in life, and higher age is a significant risk factor for COVID-19 mortality and morbidity ^56^, while lower age an important predictor of vaccine hesitancy ^5 23^, making it sensible to plot correlations with hesitancy against age, as did Ganna and Ingelsson, which helps both in visibility and interpretability.

Figure 1 shows a scattergram of the correlation of each phenomic variable with age (horizontal) and with hesitancy (vertical). Significance levels are shown by size and shape of symbols, and colours indicate question type (see Table 1). Vaccine hesitancy correlated r=0.183 with age (p=9.79×10^−94^), after adjusting for hesitancy question type (unadjusted r=-0.171, p=1.55×10^−81^).

**Figure 1:**
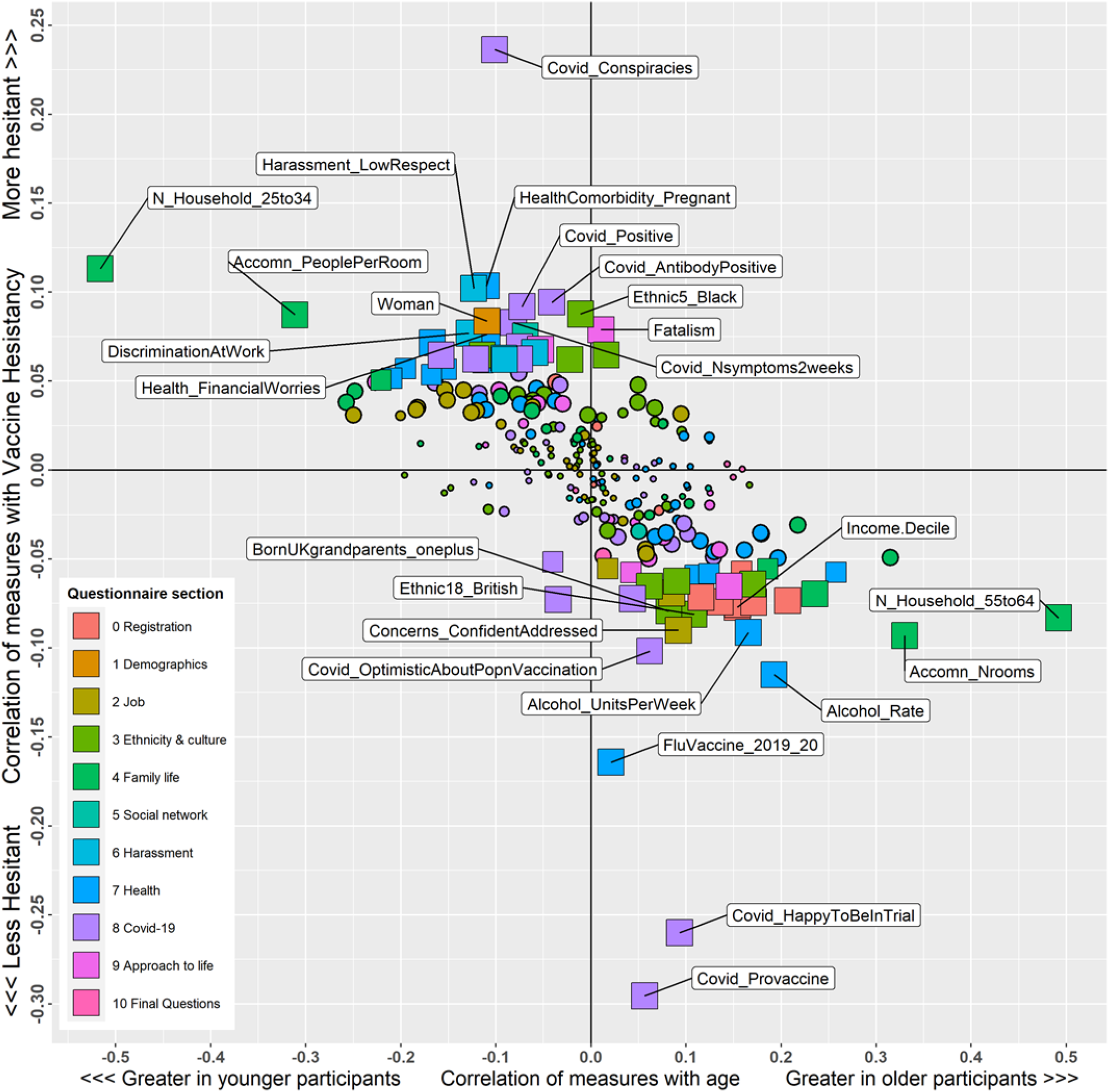
Phenome plot for 257 measures in the study. The *y* axis shows the simple correlation of each measure with vaccine hesitancy, after partialling out effects of *VaccQ_Jan*. The *x* axis shows simple correlations of each measure with age. Significance of correlations with vaccine hesitancy is shown by the size and shape of symbols. Small, medium and large circles have significance levels of >.05, <.05 and <.001, none of which reach the criterion of <4.04×10^−8^. Smaller and larger squares reach the criterion of p<4.04×10^−8^, with larger squares indicating <4.04×10^−11^. Labels point to the 25 measures with the highest correlations, and all have p<1.88 × 10^−16^. Colour of points indicates the section of the questionnaire from which they are derived (see key for details).

Of the remaining 257 measures, 68 (26.4%) were significantly correlated with hesitancy at the adjusted criterion of p=4.02×10^−8^. Of the 68 measures correlated with hesitancy only two had correlations greater than the correlation of hesitancy with age: lower pro-vaccine score (r= −0.296), and higher COVID-19 conspiracy beliefs score (r=0.236). Many of the measures associated with hesitancy in Figure 1 were also associated with age, the correlation between the correlation of hesitancy with age and the correlation of hesitancy with individual measures being r=-0.620. As can be seen from the strong downward slope of the points in Figure 1, measures associated with greater age were associated with less vaccine hesitancy. As such there is much potential for spurious correlations of measures with hesitancy due to correlations with age. The statistical challenge is to identify which of the many variables plotted in Figure 1 are indeed predictors of vaccine hesitancy.

### *islasso* analysis of predictors of vaccine hesitancy

Figure 2a, uses an extension of the plotting method used in Figure 4 of Cilluffo et al ^46^, to show the distribution of the simple correlations with hesitancy already seen in figure 1. Significance levels and the order of the correlation are shown on log10 scales for ease of visualisation. 68 correlations reach the significance level of <4.02 × 10^−8^. Figure 2b shows the *islasso* analysis, with only six of the 258 phenomic measures being significantly related to vaccine hesitancy at the criterion of p<4.02 × 10^−8^. Hesitancy was greater in those with lower pro-vaccination attitudes; who had not had a flu vaccination in the winter of 2019-20; who were pregnant; who scored higher on the COVID-19 conspiracy beliefs scale; who were younger; and who were less optimistic about the prospects for the roll-out of population vaccination. The dummy variable for vaccine question type, which was required to be in the model, did not reach the overall significance criterion but had a raw unpenalized significance of p=.00096. The difference between Figures 2a and 2b is striking, and shows how *islasso* has reduced the significant measures from 68 to 6.

**Figure 2:**
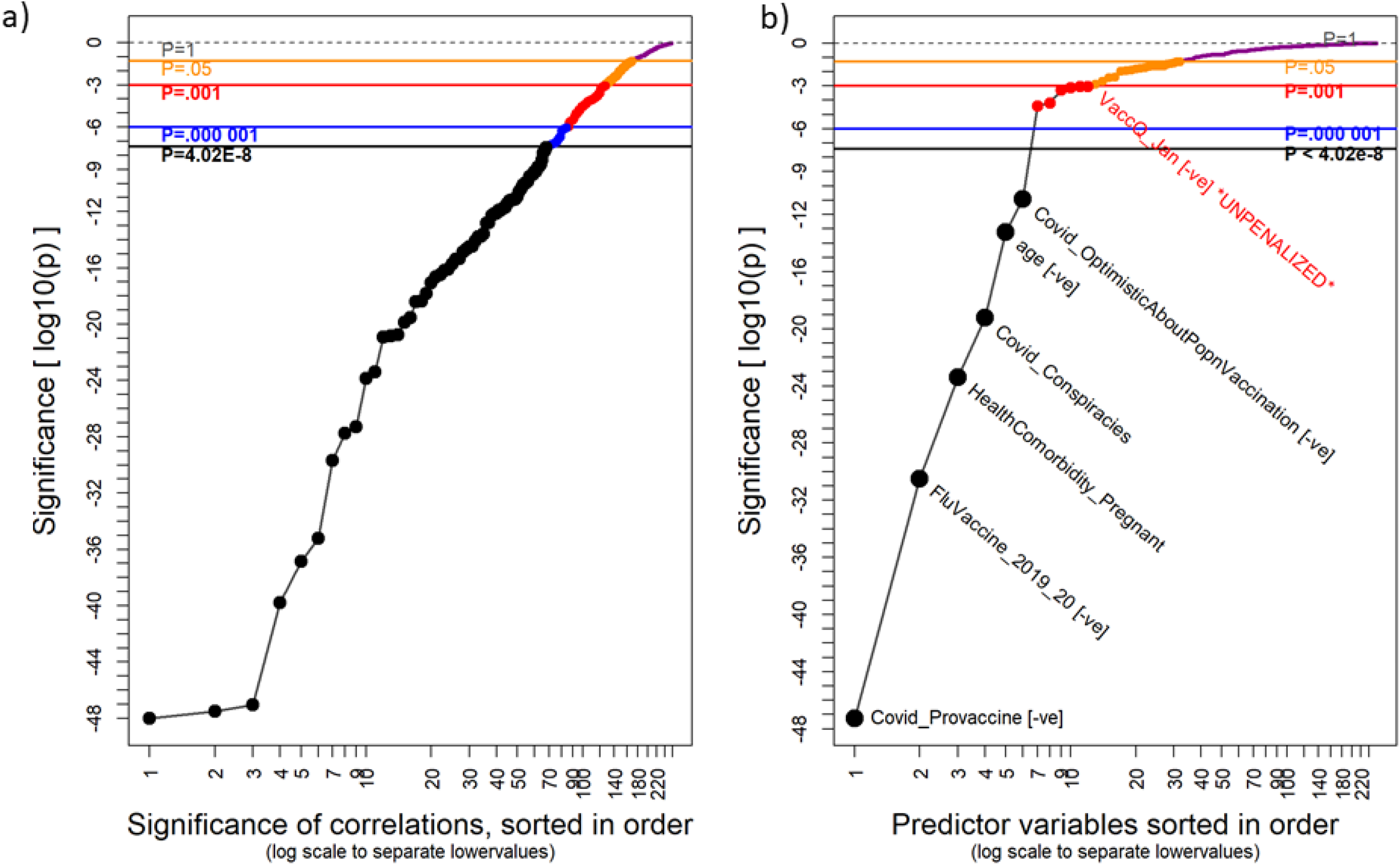
Significance level distributions for a) simple correlations of measures with hesitancy (after partialling out VaccQ_Jan); and b) results of the *islasso* analysis of hesitancy. Significance levels are plotted on the y axis, on a logarithmic scale. Note that three very significant correlations (p=2×10^−134^, 2×10^−120^,9×10^−74^) have been censored at 10^−48^. The *x* axis shows variables in ascending order of significance, on a logarithmic scale to make it easier to see the most significant measures. Black dots indicate significance of p<4.02×10^−8^, and names are provided in (b) only for the six measures which reach that criterion, and for *VoccQ_Jon* which is unpenalized (see text). Significance levels shown for p=l, .05, .001 and .000001 are for illustrative purposes only.

### Path model of the predictors of COVID-19 vaccine hesitancy

The final step in the analysis uses a path model to ask what variables predict each of the significant variables in Figure 2, and then what variables predict those variables, and so on. The final model is summarized in Figure 3. The inner circle, ①, shows vaccine hesitancy, and the second circle, ②, the six variables that predict vaccine hesitancy (as well as the version of the vaccine questionnaire). The third circle, ③, shows predictors of the variables in circle ② that meet the criterion of p<3.11 × 10^−10^ (see statistical significance levels, above); and so on for circles ④ to ⑥. Green arrows indicate variables that increase vaccine hesitancy, and red arrows those that reduce it. Standardized path coefficients are shown alongside arrows, and arrow width is proportional to the effect size.

**Figure 3:**
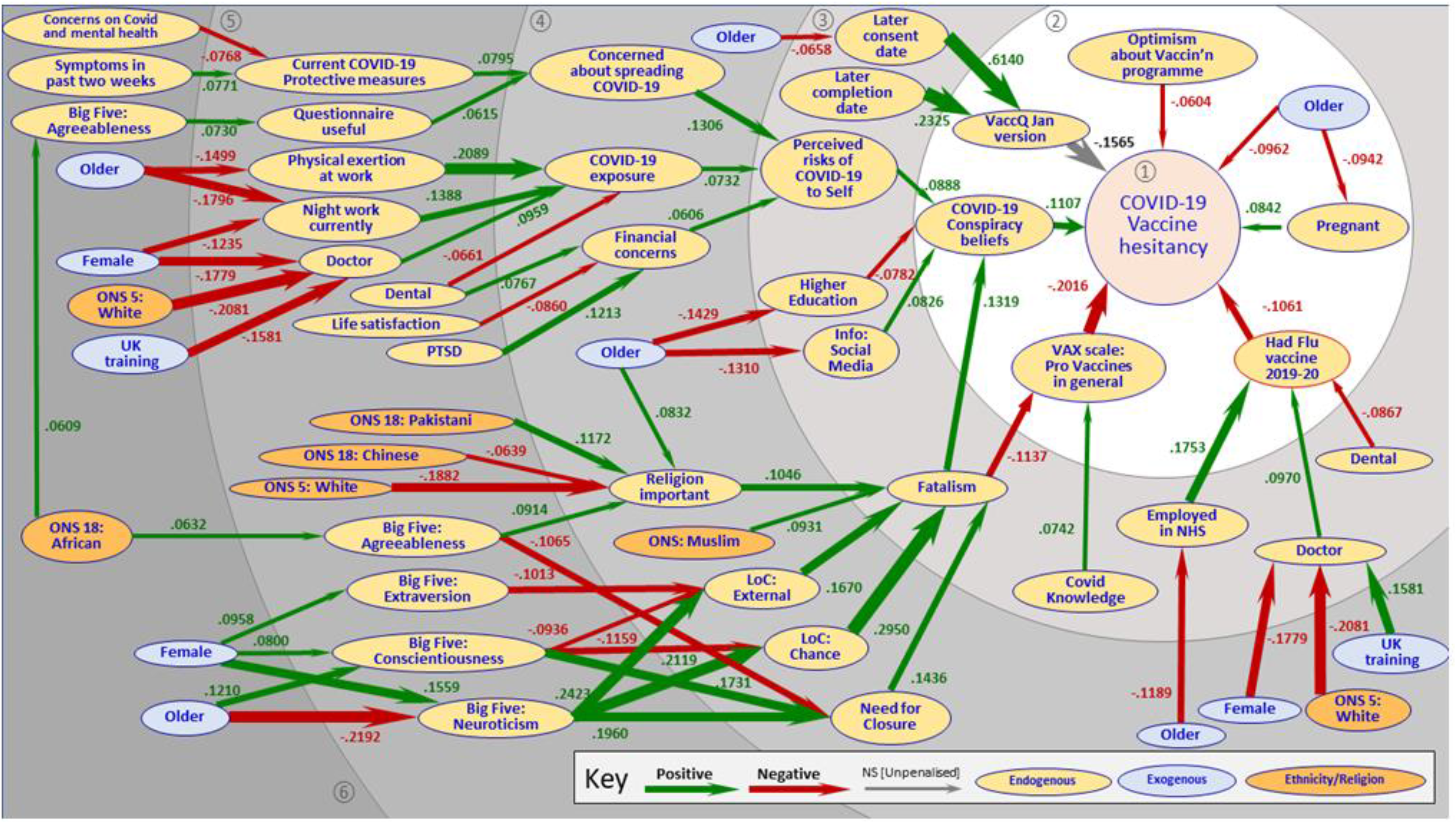
Path model showing direct and indirect predictors of vaccine hesitancy (including VaccQ_Jan) using *islasso* to assess significance of predictors of predictors, etc.. Path coefficients are shown as standardized beta coefficients from linear regressions and can directly be compared in magnitude. Green lines indicate positive regression coefficients, i.e. higher values of predictors indicate higher outcomes of dependent variables; all labels indicate the direction of coding of the measures. Red lines indicate negative regression coefficients. Circles indicate the successive analyses, with the primary analysis being of vaccine hesitancy itself. Measures in yellow are endogenous, and measures in blue or orange are exogenous, measures of ethnicity or religion being shown in orange.

The final path model in Figure 3 includes 46 direct and indirect predictors of hesitancy of which eight are demographic (including age, sex, and six measures of ethnicity and religion) and therefore exogenous.

Four of the six direct predictors of vaccine hesitancy have few other predictors. *Optimism about vaccination programmes* has no other predictors and *age* is exogenous. *Pregnancy* is predicted only by *age*. Receiving an *influenza vaccination* is predicted by occupational factors, themselves predicted by demographic factors. By contrast, *COVID-19 conspiracy beliefs* and *pro-vaccine attitudes* are underpinned by long chains of causation, with one chain primarily relating to perceived risks of COVID-19 and exposure to COVID-19, and the other chain mainly relating to psychological factors and religion. Ethnic differences are mostly observed on the latter chain. While *pro-vaccine attitudes* are largely predicted by factors within the psychological and religion-related chain, *COVID-19 conspiracy beliefs* are predicted by both the perceived risk and exposure chain and the psychological and religion chain via *fatalism. Fatalism* itself is influenced by religion being important in one’s daily life, particularly for Muslim participants, as well as by psychological factors that generally drive attitudes and behaviours, particularly chance and external locus of control, the Need for Cognitive Closure (aversion to ambiguity), high Neuroticism (anxiety-proneness) and low Extraversion (low sociability).

The *islasso* path model found no direct influences of ethnicity or religion on hesitancy at the adjusted significant criterion, and effects were instead mediated via various other factors in complex ways. Muslim participants are on average higher on fatalism. Fatalism is predicted by *religiosity*, which is higher in the Pakistani group and lower in the White and Chinese groups. The African group have higher agreeableness scores which increase hesitancy indirectly via greater religiosity, but also reduce hesitancy indirectly via lower need for closure. Finally, White ethnicity is related to greater hesitancy indirectly because White participants are less likely overall to be doctors, and doctors are more likely to have had influenza vaccination.

In terms of other demographic predictors, age is the only factor to directly predict hesitancy, with older HCWs being less hesitant, but the model also shows complex indirect effects of age mediated by working patterns, education and social media information use. With regards to sex, all of those in our survey who were pregnant self-identified as female, and pregnancy was a strong direct predictor of hesitancy. Other influences of sex were more complex being mediated in both positive and negative directions via personality, working patterns and working roles.

### Measures not included in the path model

Although in Figures 1 and 3 we have described many measures that seem to influence vaccine hesitancy, it is also clear that many of our 259 measures do *not* influence hesitancy. Some of the ‘top 25’ in Figure 1 do not appear in Figure 3, and do not have any obvious relation to hesitancy. As an example, rate of using alcohol and alcohol units per week are associated with lower rates of hesitancy, but in the Gaussian Graphical Model in the Supplementary Information^j^ have positive partial correlations with older age, being male, being White, and negative partial correlations with being Muslim, and religion being important in everyday life^k^, all of which are related to hesitancy in Figure 3. Likewise, the interim analysis in our previous paper ^5^ with a more restricted set of measures had suggested that confidence in concerns being addressed at work, and feeling secure at raising concerns might be predictors of lower hesitancy. Neither measure though makes it through to Figure 3. The only variable from Figure 3 showing a partial correlation with confidence is Agreeableness, higher scores of which predict hesitancy via religion being more important, lower need for closure, higher fatalism, higher conspiracy beliefs and lower pro-vaccine beliefs in general.

## Discussion

Vaccine hesitancy has been much studied, not only during the COVID-19 pandemic but for many years previously ^31 57 58^, particularly in relation to influenza vaccination and childhood vaccinations which have major public health implications. HCWs are not vaccine hesitant, in general ^59^ or to COVID-19 vaccination in particular ^4^. Nevertheless, a quarter or so of HCWs in our study *are* vaccine hesitant. While vaccine hesitancy in general is well-recognized, the underlying behavioural processes are not so well described. Although many studies find associations of hesitancy with individual psychological or social measures, most of those studies are relatively small with small numbers of measures. A scoping review of 95 studies (median N ∼ 1400) found 57 factors reported to be related to hesitancy (25 contextual, 22 individual or group, 10 vaccine-related determinants)^60^.

The present study has several major strengths over previous research for studying hesitancy. UK-REACH with 12,431 participants is in the top 1% by size of studies examining hesitancy^60^, and is one of the largest studies of HCWs. UK-REACH focusses on UK HCWs who have been deeply enmeshed within the COVID-19 pandemic, and it is broad, with several hundred measures including social, cognitive and behavioural scales, information on culture, religion and ethnicity, and detailed assessments of work experiences. This observational study was undertaken at the peak of the second UK wave of the COVID-19 pandemic, at the start of the UK vaccine roll-out.

Interpreting the large amount of data, and how it relates to vaccine hesitancy, benefited from the phenomic approach, which assessed the relationships of the large set of phenotypic measures to an outcome to which they are plausibly related, analogously to the way a GWAS assesses large numbers of genotypic measures which potentially are related to a particular outcome, without having to make arbitrary or *a priori* choices for inclusion. Our analyses of vaccine hesitancy identified direct and indirect causal pathways from a wide range of variables describing the working lives, home lives, behaviours, attitudes and personalities of UK HCWs from diverse ethnic groups. While our study measures self-reported vaccine-related behaviours and intentions, and not vaccine uptake directly, there is increasing evidence that survey measures of COVID-19 vaccine hesitancy do reflect actual vaccine uptake across countries ^61^. Our survey was voluntary and administered during the difficult NHS working conditions in the UK second wave, and as such could have been affected by response bias; however, the cohort respondents’ characteristics were broadly similar to the wider NHS workforce^l^. The cohort did however include only a small number of ancillary staff, reflecting the methods of recruitment, which limits generalisability to that group.

The validity of the phenomic approach is shown by its identification of pregnancy, younger age, and lack of previous influenza vaccination as major demographic predictors of hesitancy, which we had identified previously^5^, as well as in identifying low pro-vaccine attitudes and COVID-19 conspiracies as important hesitancy predictors, which concurs with other studies of COVID-19 vaccine hesitancy ^3 62–65^.

The phenomic approach also allowed the construction of a path model (Figure 3) showing 46 direct and indirect influences on COVID-19 vaccine hesitancy, whereas previous studies had mainly looked entirely at direct influences or simple correlations. It is also of interest that as well as the 46 influences, there were also 212 measures which within the path model showed *no* evidence of direct or indirect effects upon hesitancy, and they are also of importance and theoretical interest. We also acknowledge that there are myriad other cognitive measures that could have been included, although that would not have been practically feasible, and will be of interest for future studies.

We found three clusters of measures of particular interest in Figure 3, although all of the individual paths have interpretations of potential interest. Many of the routes pass through COVID-19 conspiracy beliefs and the VAX scale of pro-vaccine attitudes in general. In particular we note:

i. *Conspiracy theory beliefs*. The proliferation of false information about COVID-19 has been called an “infodemic” by the WHO ^66^. There is growing evidence that increased hesitancy, as well as reduced intentions to comply with public health behaviours such as mask-wearing, can be related to false information and conspiracy theories, which decrease trust in the safety and efficacy of COVID-19 vaccines, reduce perceptions of the risks posed by COVID-19, and increase vaccine hesitancy ^67–69 7 70^. Defining conspiracy theories (CTs) is not easy ^71^ but broadly they are, “narratives about events or situations, that allege there are secret plans to carry out sinister deeds” ^72^. Research into CTs has grown in recent decades, particularly in understanding CTs (“conspiracy theory theory” ^73^). CTs have existed since ancient times ^74^, with “ethnographies of suspicion” appearing to be universal across time and cultures ^75^, and four key principles of being consequential, universal, emotional and social ^76^. Some conspiracies are, of course, real and then represent existential threat, and CTs may well have been adaptive in hunter-gatherers subjected to intergroup conflict and aggression ^77^. CTs about medical interventions have been prevalent at least since Jenner introduced vaccination ^72^, and the influenza A-H1N1 pandemic of 2009 had many CTs ^78^. A wide-range of CTs in relation to the origins of COVID-19 and to COVID-19 vaccination have been described ^79^, and the present study uses the CTs described by Duffy and colleagues ^7 80^. CTs should be distinguished from Conspiracy Theory Beliefs (CTBs) ^81^ in which CTs appear plausible to quite large proportions of the population ^82^, perhaps in part because of greater spread though social media ^83^. Different CTBs tend to be strongly associated with one another, individuals often having multiple CTBs ^81^, even if the formal content of the underlying CTs themselves is unrelated. Anti-vaccination attitudes co-occur with unrelated CTBs, suggesting that, “anti-vaccination beliefs are best explained as an extension of a common psychological predisposition for conspiracy beliefs”, and are a form of CTB ^84^. Reviews suggest that many psychological measures are potentially related to a tendency to believe in CTBs ^85–87^, although studies are often small and consider only a few measures. As well as being influenced by fatalism, described above, CTBs have several other sets of influences:
  a. *Education and information*. CTBs in Figure 3 are less frequent in study participants who have higher education, and more frequent in those who use social media more. Studies have suggested that CTBs are associated with lower critical thinking ability ^88^, and lower analytical ability ^89^, which partly mediates the association of CTBs with less education ^90^. Higher CTBs have also been associated with greater checking of social media ^80^.
  b. *Perceived risks to self*. A large cluster of measures in Figure 3 relate to CTBs via perceived risks of COVID-19 to the self, including concerns about spreading COVID-19 (related to current protective measures, recent symptoms, and worries that COVID-19 will affect the mental health of the population), greater COVID-19 exposure (related to more physical exertion at work, more night work, and being a doctor), and financial concerns (related to lower life satisfaction, more symptoms of PTSD, and working in dentistry), with most of those measures being plausible concerns about COVID-19 risk. Less clear is the mechanism for links to CTBs, but the literature suggests CTBs are often epistemic, reflecting a need for clarity and certainty, and existential, relating to a need to feel safe ^24 91^.. In contrast to the WHO’s “Three Cs” model of hesitancy ^22 30–32^, participants who perceived themselves to be at higher risk of COVID-19 were more hesitant about being vaccinated, with the relationship being indirect via increased COVID-19 conspiracy beliefs.
ii. *Fatalism*. Fatalism is a set of beliefs centred around predetermination, luck and pessimism ^39^. While fatalism generally permeates many aspects of thought, within health psychology it is seen as an answer to, “why do patients not follow their doctor’s advice [… particularly] for a healthy lifestyle, screening for disease or indeed treatment itself?” ^92^. We found fatalism to be associated both with negative attitudes to vaccines in general, and with higher COVID-19 conspiracy theory beliefs, both then resulting in greater COVID-19 vaccine hesitancy. Fatalism itself is influenced by two large clusters of measures, which broadly relate to personality and religion.
  a. *Personality*. The path diagram shows strong relationships of personality and fatalism, high fatalism scores being associated with higher chance locus of control and external locus of control, and also a greater need for cognitive closure. In Figure 3, locus of control and need for closure are all related to Big Five personality measures, particularly Neuroticism, with its association with anxiety proneness. Fatalism is also associated indirectly with lower extraversion, but the relationship to agreeableness and conscientiousness is complicated. The literature suggests that fatalism and need for cognitive closure have both been related to CTBs ^81 93 94^. Locus of control is also related to vaccination intention, as well as religiosity which it partly mediates ^95^.
  b. *Religion*. Fatalism was higher in those saying that religion is important in their everyday life, and is also higher in Muslims. Those of Pakistani heritage say that religion is more important, but religion was less important for White and Chinese participants. The literature suggests that it is “a commonplace” that fatalism is higher in Muslims ^96^, with it “embedded in Middle Eastern societies”^97^, but cross-cultural studies suggest a more Durkheimian view that more regulated societies are more fatalistic ^98^, with few differences between faiths themselves. It is also suggested that fatalism is greater in minority religions within a society, and relates to structural inequalities in power, wealth, privilege and health ^92 99^. Although treated as a trait, there is evidence that fatalistic attitudes might be changeable, which might include by media exposure ^100^.
iii. *Ethnicity and demography*. Ethnicity and other demographics, particularly age and sex, are of interest to UK-REACH.
  a. *Ethnicity and religion*. Being Muslim was the only religious group related indirectly to hesitancy, via fatalism. Fatalism was also influenced by religion being important in everyday life, which in turn was higher in those of Pakistani ethnicity, but lower in Chinese and White HCWs, resulting in lower hesitancy. African ethnicity influenced Big 5 Agreeableness, which also influenced religion being important in everyday life, and a greater need for closure.
  b. *Age*. Younger age is undoubtedly a strong predictor of vaccine hesitancy ^5 23^. Age is the only demographic measure in Figure 3 with a direct influence on hesitancy, and since it is exogenous that association cannot be taken apart further. Presumably it is not, however, being mediated by any of the other 257 variables included within the analysis. Age does however have other indirect effects, as older participants were less likely to work in the NHS, had different working patterns, were less likely to have higher education, used social media less, were more religious, more conscientious, had lower neuroticism scores, and responded to the questionnaire sooner.
  c. *Sex*. In previous studies women have been more hesitant about COVID-19 vaccination ^4 101^, and the highly significant simple effect can be seen in our data in Figure 1. Although female sex was not a direct predictor of hesitancy in our data, sex did have indirect influences, via personality measures, different working patterns, and areas of work.

### Interventions and implications

Even before COVID-19, vaccine hesitancy was described by the WHO has one of the ten leading threats to global health, with complacency, inconvenience, and lack of confidence as primary drivers ^102^. A special issue of *Vaccine* in 2015 was devoted to vaccine hesitancy^103^, and included a review identifying 166 evaluations of interventions (and 983 papers describing but not evaluating interventions), and although some studies did show a positive effect, heterogeneity, “limited [the] ability to draw many general conclusions about the effectiveness of different strategies”^57^. In the same issue, an analysis of fifteen reviews and meta-analyses concluded there was “no strong evidence to recommend any specific intervention”, that, “educational tools … had little or no impact”, and that, “some communication interventions could even reinforce vaccine hesitancy” ^104^. More broadly was emphasized the influence of social norms and social networks, as well as interactions with healthcare providers^104^, following work on the ‘big picture’, which emphasized that, “addressing vaccine hesitancy involves developing a deep understanding of the psychological and social dimensions of vaccine acceptance … [with] interventions operating at the individual, family and community level”^105^. Models of cognitive obstacles to pro-vaccination beliefs^106^ are still rare, but suggest that “psychological mechanisms can conspire to render pro-vaccination beliefs counter-intuitive” through mechanisms such as omission bias, intuitive responses to disgust, and low perceived saliency of disease^106^.

Effective interventions to reduce vaccine hesitancy have been seen as ever more important in the era of COVID-19, with the tensions shown in two recent opinion pieces in *Nature*, one arguing for, “direct, even confrontational, approaches … [to] anti-vaccine groups”^107^, while a response to that paper argued for, “constructive ways … centred around respect, openness and empathy”^108^. Researchers on cognitive obstacles have warned that although,

“forceful incentives for vaccination … might make sense in the short term, they are liable to create more distrust towards the very institutions that need to be trusted”^106^. The role of trust has been emphasised by Maya Goldenberg in her book *Vaccine Hesitancy*^109^, with the need to harmonize values with fact-based decision-making^110^, and not to make “facile calls to trust science”^110^. Interestingly, Goldenberg does not consider hesitancy in HCWs, although they present a theoretical problem, since HCWs “are the public face of vaccine advocacy”^109^(p.54), potentially putting them on both sides of the ‘them’ and ‘us’ divide. For Goldenberg there is not so much a war on science as “a crisis of trust”^110^, and trust does seem to be an issue, as identified in the UK-REACH qualitative study of HCW hesitancy^8^. Trust and mis-trust are rooted in ‘Epistemic vigilance’^111^, which involves the monitoring of communications to avoid being misled by others (which is always a risk in society). There are elaborate mental and social mechanisms for evaluating communications, and thereby associating them with trust^106^, which may be at odds with a purely scientific or evidence-based approach. Greater trust in employers and the NHS were associated with subsequent vaccination in our recent follow-up study of HCWs hesitant on our earlier questionnaire.^112^

Our study fits well into such approaches. Of the two legs of the ‘big picture’ ^104 105^, interactions with healthcare providers seem of marginal relevance to hesitancy in a population of HCWs, making the influence of social norms and social networks particularly salient. Conspiracy beliefs and pro-vaccine attitudes, which are both influenced by fatalism, which is influenced by religion, locus of control and personality, with clear ethnic and religious differences, are all clearly “imbued [with] social, cultural, and historical context”^110^ as well as associations with trust.

The UK-REACH study was not set up to investigate interventions for hesitancy, but our Figure 3 does suggest several pinch-points at which interventions to alter vaccine hesitancy might be relatively more effective. Addressing the influence of fatalism, which may well take different forms in different groups, might allow tailoring of interventions to the needs of different communities, with values, attitudes and beliefs at the centre of the process. However, given the complexity of Figure 3, it should be remembered that simple solutions to complex problems may neither be easy nor effective.

## Summary

A large sample size and a very broad range of measures allows UK-REACH a much more detailed picture of vaccine hesitancy than in most other studies, which are smaller in size and scope. Our study adds to the rapidly growing body of knowledge on vaccine hesitancy in putting together a broad picture of how all these factors relate to influence hesitancy in HCWs. We do not doubt that more complex structural equation models could be fit to the vast mass of data, and we look forward to other researchers exploring further within these data.

Although our study entirely concerns HCWs in a single country, we believe that much of it will likely generalize to a wider population, at least in high-income countries^113 114^, although we note that at a societal (macro level), vaccine confidence does relate to trust in science^16^. The NHS is the largest employer in the UK, and our sample is broadly representative of NHS employees, who have very many skills, backgrounds, and job types. The advantage of studying HCWs is that it was also possible to measure working conditions in a meaningful way across the well-defined and COVID-19 relevant working environment of hospitals and clinics.

While our phenomic analysis cannot claim to be comprehensive (just as few genomic studies can consider all base pairs in the genome, and any description of the ‘environmentome’ ^115^ for HCWs must be far into the future), the path modelling approach has enabled us to infer potential causal pathways. These can be tested with linked longitudinal data from further waves of the UK-REACH cohort study which are under way, and with more interventional research designs.

The complex nexus of causes and effects behind the apparently simple behaviour of being COVID-19 vaccine hesitant is clear from Figure 3. The most important predictors of COVID-19 vaccine hesitancy are psychological measures of vaccine-related attitudes and beliefs, as well as previous vaccine-related behaviour (influenza vaccine uptake). That is hardly surprising, as hesitancy is a behaviour, and behaviours tend to be explained best by other behaviours, attitudes and cognitions. Other than age, most demographic effects are mediated via other factors. Hesitancy may correlate with demographic and other measures, but the effects are not direct, and instead are mediated through a range of behaviours and cognitions in order to impact upon COVID-19 vaccine hesitancy. The implications for interventions to reduce hesitancy are discussed.

## Supporting information

PDF for looking at XLS file of variables

## Data Availability

To access data or samples produced by the UK-REACH study, the working group representative must first submit a request to the Core Management Group by contacting the UK-REACH Research Manager in the first instance. For ancillary studies outside of the core deliverables, the Steering Committee will make final decisions once they have been approved by the Core Management Group. Decisions on granting the access to data/materials will be made within eight weeks. Third party requests from outside the Project will require explicit approval of the Steering Committee once approved by the Core Management Group. Should there be a significant numbers of requests to access data and/or samples then a separate Data Access Committee will be convened to appraise requests in the first instance.

## Contributors

MP conceived of the idea and led the application for funding with input from MDT, KK, ICM, KW, RF, LBN, SC, KRA, LJG, ALG and CJ. The survey was designed by KW, MP, ICM, CMel, CJ, ALG, LBN, RF and CAM. Online consent and survey tools were developed by LB. ICM wrote the first draft of the manuscript with subsequent input from MP, KW and all co-authors. All authors approved the submitted manuscript.

## Acknowledgements

We would like to thank all the healthcare workers who took part in this study when the NHS was under immense pressure.

We wish to acknowledge the Professional Expert Panel group (Amir Burney, Association of Pakistani Physicians of Northern Europe; Tiffanie Harrison; London North West University Healthcare NHS Trust; Ahmed Hashim, Sudanese Doctors Association; Sandra Kazembe, University Hospitals Leicester NHS Trust; Susie M. Lagrata (Co-chair), Filipino Nurses Association, UK & University College London Hospitals NHS Foundation Trust; Satheesh Mathew, British Association of Physicians of Indian Origin; Juliette Mutuyimana, Kingston Hospitals NHS Trust; Padmasayee Papineni (Co-chair), London North West University Healthcare NHS Trust; Tatiana Monteiro, University Hospitals Leicester NHS Trust) and UK-REACH Stakeholder Group (see the cohort study protocol^33^ for details), the Study Steering Committee, SERCO, and the following people for their support in setting up the study from the regulatory bodies: Kerrin Clapton and Andrew Ledgard (General Medical Council), Caroline Kenny (Nursing and Midwifery Council), David Teeman and Lisa Bainbridge (General Dental Council), My Phan and Jenny Clapham (General Pharmaceutical Council), Angharad Jones (General Optical Council), Katherine Timms and Charlotte Rogers (The Health and Care Professions Council) and Mark Neale (Pharmaceutical Society of Northern Ireland).

We would also like to acknowledge the following trusts and sites who recruited participants to the study: Nottinghamshire Healthcare NHS Foundation Trust, University Hospitals Leicester, Lancashire Teaching Hospitals NHS Foundation Trust, Northumbria Healthcare, Berkshire Healthcare, Derbyshire Healthcare NHS Foundation Trust, South Tees NHS Foundation Trust, Birmingham and Solihull NHS Foundation Trust, Affinity Care, Royal Brompton and Harefield, Sheffield Teaching Hospitals, St George’s Hospital, Yeovil District Hospital, Lewisham and Greenwich NHS Trust, Black Country Community Healthcare NHS Foundation Trust, Sussex Community NHS Foundation Trust, South Central Ambulance Service, University Hospitals Coventry and Warwickshire, University Hospitals Southampton NHS Foundation Trust, London Ambulance Trust, Royal Free, Birmingham Community Healthcare NHS Foundation Trust, Central London Community Healthcare, Chesterfield Royal Hospital, Bridgewater Community Healthcare, Northern Borders, County Durham and Darlington Foundation Trust, Walsall Healthcare NHS Trust.

## Data sharing

### Transparency statement

The lead authors affirm that this manuscript is an honest, accurate, and transparent account of the study being reported; that no important aspects of the study have been omitted; and that any discrepancies from the study as planned (and, if relevant, registered) have been explained.

## Funding

UK-REACH is supported by a grant from the MRC-UK Research and Innovation (MR/V027549/1) and the Department of Health and Social Care through the National Institute for Health Research (NIHR) rapid response panel to tackle COVID-19.

Core funding was also provided by NIHR Biomedical Research Centres.

KW is funded through an NIHR Career Development Fellowship (CDF-2017-10-008).

CAM is an NIHR Academic Clinical Fellow (ACF-2018-11-004).

LBN is supported by an Academy of Medical Sciences Springboard Award (SBF005\1047).

ALG was funded by internal fellowships at the University of Leicester from the Wellcome Trust Institutional Strategic Support Fund (204801/Z/16/Z) and the BHF Accelerator Award (AA/18/3/34220).

MDT holds a Wellcome Trust Investigator Award (WT 202849/Z/16/Z) and an NIHR Senior Investigator Award.

KK and LJG are supported by the National Institute for Health Research (NIHR) Applied Research Collaboration East Midlands (ARC EM).

KK and MP are supported by the NIHR Leicester Biomedical Research Centre (BRC). MP is supported by a NIHR Development and Skills Enhancement Award.

This work is carried out with the support of BREATHE - The Health Data Research Hub for Respiratory Health [MC_PC_19004] in partnership with SAIL Databank. BREATHE is funded through the UK Research and Innovation Industrial Strategy Challenge Fund and delivered through Health Data Research UK.

## Disclaimers

The views expressed in the publication are those of the author(s) and not necessarily those of the National Health Service (NHS), the NIHR or the Department of Health and Social Care. This research was funded in whole, or in part, by the Wellcome Trust [WT204801/Z/16/Z and WT 202849/Z/16/Z]. For the purpose of open access, the author has applied a CC BY public copyright licence to any Author Accepted Manuscript version arising from this submission.

## Footnotes

a For simplicity we use COVID-19 to describe the disease whereas SARS-Cov-2 is the causative agent. Vaccines act specifically on SARS-Cov-2, but the public health vaccination programme is against the disease COVID-19. Vaccine hesitancy is a behaviour and can be regarded as against COVID-19 vaccination (and hence COVID-19) and not against vaccines for SARS-Cov-2 *per se*.

b Supplementary Information: “Assessing the extent of response bias”

c Supplementary Information: “Descriptions of particular variables / Vaccine hesitancy”

d Supplementary Information: “Descriptive summaries of particular variables”

e Supplementary Information: “Overview of variables in the analysis and in the multiple imputation”; “Summary of the multiple imputation”; “Selection of variables”; “Setting of a significance level”; “Analysis of predictors of predictors”.

f See Supplementary Information, “Overview of variables in the analysis…”, and “Summary of the multiple imputation.

g See Supplementary Information, “Setting of a significance level”

h See Supplementary Information, “Gaussian graphical models”

i Supplementary Information: “Descriptive measures by vaccine hesitancy”.

j See Supplementary Information, “Gaussian graphical models”

k See Supplementary Information, “Gaussian graphical models”

l See Supplementary Information, “Assessing the extent of response bias”

m https://uniofleicester-my.sharepoint.com/:x:/r/personal/mp426_leicester_ac_uk/_layouts/15/Doc.aspx?sourcedoc=%7B79688523-78DA-4968-8CA4-29DD01240921%7D&file=UK-REACH_questionnaire_data_dictionary_v1.0.xlsx&action=default&mobileredirect=true

