## Supplementary Material for "Vaccine hesitancy for COVID-19 explored in a phenomic study of 259 socio-cognitive-behavioural measures in the UK-REACH study of 12,431 UK healthcare workers"

### Overview of variables in the analysis and in the multiple imputation.

Overall there are many raw and derived variables in the analysis, and they are summarised in Table 1 of the present main paper, which is similar to that in the Supplementary Material[a] for the interim report[1].

#### Variables.

Overall the questionnaire had 797 raw items to which participants might respond, although not all were available to all participants, logic chains making them contingent on earlier responses. Using Vaccine question VQ2 as an example (see Supplementary Figure 1, below), the main item has six possible responses (including PNTA, 'prefer not to answer'). Three responses have subsidiary items each with three possible responses. VQ2 therefore has four raw items. For the present analysis there is however one derived variable with two responses, shown in Supplementary Figure 1) as hesitant (orange) and non-hesitant (green).

Some raw items such as the ONS questions on ethnicity of self, mother, father and partner, are multinomial, each being one item with 18 possible responses (+PNTA). Derived variables consist of 18 binary variables, one for each of the 18 responses, supplemented by five binary variables for the ONS5 categories (White, Asian, Black, Mixed, Other). For ethnicity, and many other variables, derived variables are coded as 1 for the focal measure (e.g. Pakistani) and 0 for all other responses; such dummy variables tend to be negatively correlated (and that will be seen later in the *glasso* analysis). Normally, with *N* categories contrasts would be coded with *N-1* dummy variables, so that there is no collinearity between contrasts. The use of *islasso* however allows the inclusion of a set of variables which is jointly collinear (i.e. one eigenvalue is zero), and the method of penalization then drops the least significant until only those of significance remain. That has the advantage of not having to decide *a priori* which category will be the reference category, and also allows several dummy variables within a variable to be significant.

Multiple imputation places constraints on the items and not all items are included in the imputation, particularly due to multicollinearity (see below). For items such as ethnicity, the multinomial 18 item measure is used for the imputation itself, but then the derived variables are calculated in the imputed datasets, and after that the raw multinomial variables are dropped from the analysis.

Overall the imputation used 214 variables, of which 11 were multinomial, and after calculating derived variables from the ordinal and multinomial variables, and dropping the original ordinal and multinomial variables, there were 260 measures which were entered into the *islasso* analysis, one of which was vaccine hesitancy, and the other 259 were predictor variables.

*Excel file VariableSummaryForSupplementaryInformation.xlsx .* A summary of all of the variables used in the analyses is available in the Excel file *VariableSummaryForSupplementaryInformation.xlsx*. The Excel file can be sorted as required, and lists all raw items and derived variables, labelled by their *SPSS* variable name and the label from the *Data Dictionary*. Row 2 of the file summarises the counts for the different types of variable. The contents of the Excel file are summarized below in Supplementary Table 1.

[a] See Supplementary Table 2, Variables included in the imputation model, from the interim report.

*Supplementary Table 1: Variables included in the Excel file VariableSummaryForSupplementaryInformation.xlsx*

| *Column* | *Description* |
|---|---|
| **Section** | The code for the section from 0 (Registration) to 10 (Final Questions). |
| **Raw** | A flag of 1 indicates if a variable is raw (i.e. imported as such from the *RedCap* and Registration files) |
| **Derived** | A flag of 1 indicates that the variable has been derived by computation from other variables. |
| **Marker** | A flag of 1 indicates a dummy variable that is a marker to help in navigating around long SPSS or R files with many variables. They can be ignored for other purposes, and are not included in the summary table. |
| **SPSS variable** | The variable name in the main *SPSS* file, which is also imported into the *R* files. |
| **Impn** | A flag of 1 indicates that a variable has been included as a binary or continuous variable in the imputation, and a flag of 4 that it has been included as a multinomial variable. Note that predictive mean matching treats binary, ordinal and continuous variables in an equivalent way. |
| **Islasso** | A flag of 1 indicates that the variable has been included in the *islasso* analyses of vaccine hesitancy. |
| **Figure 3** | A flag of 1 indicates that the variable is included within Figure 3 in the main paper. |
| **Figure 3 label** | The label used in Figure 3 in the main paper. |
| **Path level** | The level of the variables shown in the circles of the main paper Figure 3. Level ① is Vaccine Hesitancy, and levels ② to ⑥ are the successive circles in figure 3. Level 7 indicates variables that were not included within the main Figure 3. |
| **Code** | The code numbers used for the variables in Supplementary Figures 2 and 3. |

### Summary of the multiple imputation.

15,592 HCWs registered for the survey, of whom 12,431 completed and signed off the questionnaire, and analyses are restricted to them. Of the 260 derived variables, 214 were used for multiple imputation (see Table 1 in the main text) with the remainder excluded because of errors reported by *mice()* due to collinearity. The 3,161 registrants not signing off the questionnaire had answered only a small proportion of questions, 75.5% of data points being missing (505766/670248). Of the 12,431 participants who signed off and submitted their results, 4.57% of data points were missing (121578/2660234), with a median of 3.3% missing data points per participant.

Missing data were imputed using the *mice()* algorithm in ,*R* generating 20 imputation sets, with 5 iterations for each imputation [2]. Imputation used predictive mean matching, which is robust and avoids problems with out of range and implausible imputed values [3]. Estimates and standard errors were combined across imputation sets using Rubin's Rules [4]. Calculating significance within multiply imputed datasets can be problematic for variable selection methods such as stepwise regression or conventional LASSO (least absolute shrinkage and selection operator), it often being necessary to resort to approaches such as 'vote-counting' [3]. However, *islasso* provides estimates and standard errors which can be combined using Rubin's Rules [4]. Correlations in the imputed datasets for the behavioural phenomics analysis of Figure 1 in the main text were calculated using the *micombine.cor()* function in the *miceadds* package in *R*, which also calculates partial correlations.

### Descriptions of particular variables

#### Vaccine hesitancy

The primary outcome measure is Vaccine Hesitancy, which is described in Supplementary Figure 1 of the previous, interim, report [1], which is also repeated here for the reader's convenience as the present Supplementary Figure 1.

***Supplementary Figure 1***: Caption: Diagram demonstrating how the primary outcome measure of Vaccine Hesitancy was derived from the two vaccine questions (VQ1 administered from 4th to 20th December 2020, and VQ2 administered from 21st December 2020 onwards). PNTA=prefer not to answer. Orange boxes indicate - responses coded as hesitant, green boxes indicate responses coded as not hesitant in the primary outcome measure of vaccine hesitancy.

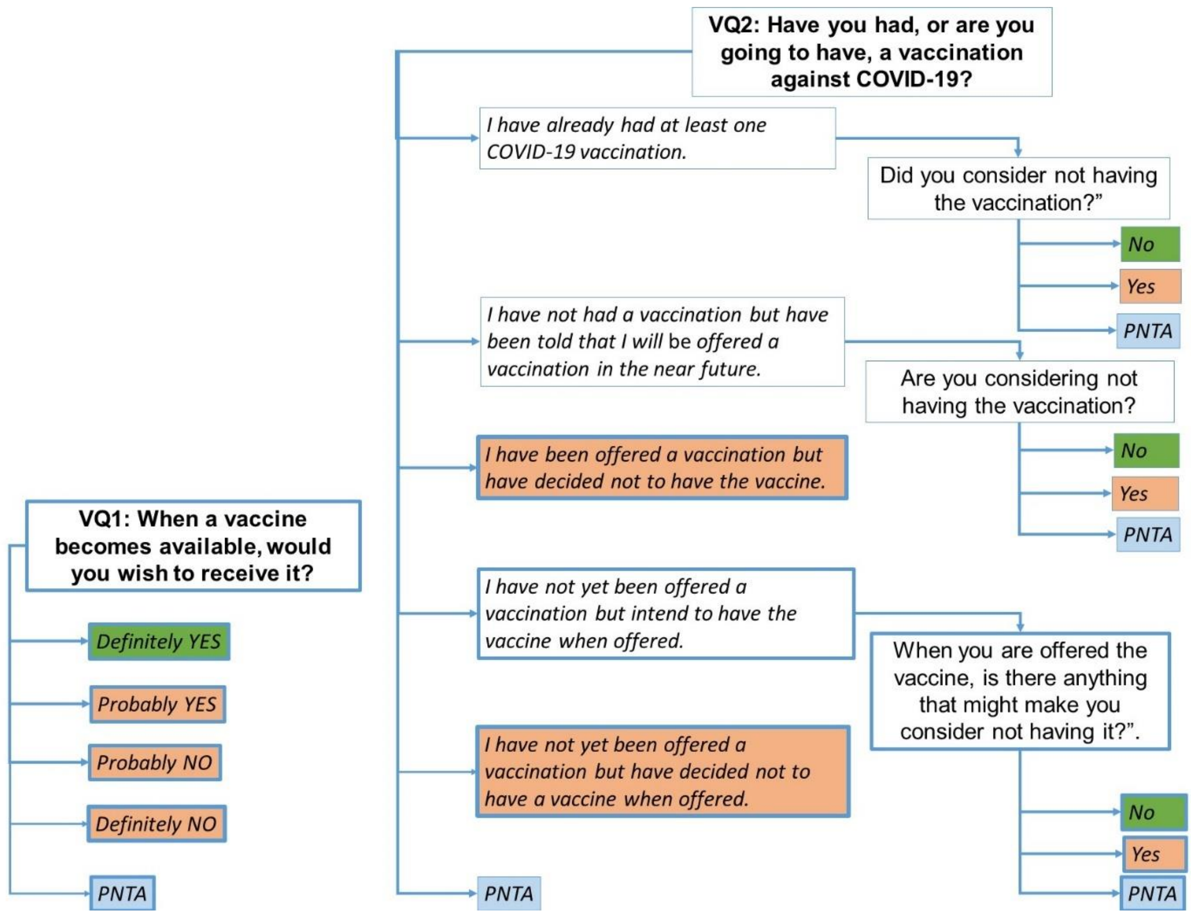

#### Ethnicity: ONS18 and ONS5 category variables

Participants reported the ethnic group with which they most identified using the 18 categories of the 2011 UK Census [5]. We also collapsed the 18 categories (ONS18) into five main ethnic categories also defined within the Census (ONS5: Black, Asian, Mixed, Other, White). There is considerable debate and controversy about the words used to describe such a broad and heterogeneous grouping, with terms such as "people of colour", "Black Asian and Minority Ethnic" or "BAME" used. In our choice of terms we have followed the BMJ who in their special edition on Racism in Medicine [6], used the term "ethnic minority" as one that is most likely to be understood by our study population, but we also include BAME for clarification. We fully acknowledge that broad ethnic groupings can mask important ethnic differences, and where possible we use the more refined ONS5 and ONS18 categories, and Supplementary Table 6 provided later shows the advantage of that approach.

### Religion and religiosity

Participants reported their religion using the UK Census 2011 question, which includes the following seven categories plus a free-text "Other" option: "*No religion", "Christian (including Church of England, Catholic, Protestant and all other Christian denominations)", "Buddhist", "Hindu", "Jewish", "Muslim", "Sikh", "Any other religion (please specify)".* Participants were also asked, "*How important is religion to you in your everyday life?*", and, "*How important was religion in your upbringing?*", each being rated on a 4-point scale from 1 "Not at all important" to 4 "Extremely important", plus PNTA.

### Pro-vaccine attitudes

Sum of scores on four items of the Vaccination Attitudes Examination (VAX) Scale [7], each measuring one of the four subscales on a 5-point Likert-type scale from 1 ("strongly disagree") to 5 ("strongly agree"): trust/mistrust of vaccine benefit ("*I can rely on vaccines to stop serious infectious diseases*"), worries over unforeseen future effects ("*Although most vaccines appear to be safe, there may be problems that we have not yet discovered*"), concerns about commercial profiteering ("*Authorities promote vaccination for financial gain, not for people's health"*) and preference for natural immunity ("*Being exposed to diseases naturally is safer for the immune system than being exposed through vaccination"*). High scores indicate pro-vaccination attitudes. The VAX Scale was included in the questionnaire from 21st December 2020 onwards.

### COVID-19 conspiracy beliefs

Sum of six scores on the COVID-19 conspiracy belief scale [8], each rated on a 4-point scale from 1 ("definitely true") to 4 ("definitely false") and then reverse-coded so that high scores indicate high COVID-19 conspiracy beliefs: *"Coronavirus was created in a laboratory", "Most people in the UK have already had coronavirus without realising it"*, "*The current pandemic is part of a global effort to force everyone to be vaccinated whether they want to or not*", "*The number of people reported as dying from coronavirus is being deliberately reduced or hidden by the authorities*", "*The symptoms that most people blame on coronavirus appear to be linked to 5G network radiation*", "*There is no hard evidence that coronavirus really exists*".

### Optimism about vaccination programme

One item "*When, if at all, do you think it will be possible to vaccinate most of the population against coronavirus?"* with 8 options, which were reverse-coded so higher scores (i.e. shorter intervals) indicate more optimism: "*1 month from now" "2 months from now" "3 months from now" "6 months from now" "12 months from now" "18 months from now" "2 years from now" "More*".

### Influenza vaccination winter 2019/2020

A binary question "*Did you have a flu vaccine last winter (2019-2020)?"* coded as no=0 and yes=1.

### Perceived risk of COVID-19 to self

Mean of responses to two questions, each rated on a scale from 0 (no possibility) to 100 (definitely will): "*What do you think is your personal chance of catching the coronavirus in the next six months?"* and "*If you do catch coronavirus, what do you think are your chances of needing hospital treatment?".* High scores indicate high perceived risk.

### Fatalism

Sum of five scores, each measured on a 7-point scale from 1 ("strongly disagree") to 7 (strongly agree"). Four items from the Fatalism Scale of Shen *et al* [9] "*If someone is meant to have a serious disease, they will get that disease*", "*My health is determined by fate", "My health is determined by something greater than myself*", "*I will stay healthy if I am lucky",* and a slightly adapted item 13 from the questions on genetic fatalism of Parrott et al [10], "*Genes are more important than one's own* *behaviour in determining one's health*". High scores indicate high fatalism (fate determining outcomes).

#### Locus of control.

Sum of scores on three subscales (Internal, Chance, External/Powerful others) of a short version of Levenson's Locus of Control scale [11]. Each subscale contained three items measured on a 7-point scale from 1 ("strongly disagree") to 7 (strongly agree"). Internal: "*My life is determined by my own actions*", "*I am usually able to protect my personal interests*", "*I can pretty much determine what will happen in my life*". Chance: "*To a great extent, my life is controlled by accidental happenings*" "*Often there is no chance of protecting my personal interest from bad luck happenings*" "*When I get what I want, it's usually because I'm lucky*". External/Powerful others: "*People like myself have very little chance of protecting our personal interests where they conflict with those of strong pressure groups*", "*My life is chiefly controlled by powerful others*" , "*I feel like what happens in my life is mostly determined by powerful people*". High scores on each subscale indicate high internal LoC (belief that one's actions determine outcomes), high chance LoC (random external events determine outcomes), and high external/powerful others (belief that powerful others determine outcomes).

#### Need for cognitive closure

Sum on two items from the short version of the Need for Cognitive Closure scale [12] each measured on a 7-point scale from 1 ("strongly disagree") to 7 (strongly agree"): "*I would quickly become impatient and irritated if I could not find a solution to a problem immediately*", "*I like having a clear and structured mode of life*". High scores indicate high need for cognitive closure and low tolerance of ambiguity.

#### Personality

Sum of scores on 15 items from Understanding Society [13] measuring the Big Five personality traits of Neuroticism, Extraversion, Openness to Experience, Agreeableness and Conscientiousness. The question stem asked "*I see myself as someone who….*" and then each item contained a statement (e.g. "…is talkative"), which participants rated on a 7-point scale from 1 "nothing like me" to 7 "exactly like me". Each trait was measured by 3 items: Neuroticism ["*worries a lot*", "…*gets nervous easily*", "*Is relaxed, handles stress well*" (reverse-coded)], Conscientiousness [*"…does a thorough job*", "…*does things efficiently*" "*tends to be lazy*" (reverse-coded)], Extraversion ["…*is talkative*", *"…is outgoing, sociable*", "…*is reserved*" (reverse-coded)], Openness ["…*is original, comes up with new ideas*", *"…values artistic, aesthetic experiences*", "…*has an active imagination*"], Agreeableness ["…*is sometimes rude to others*" (reverse-coded), *"…has a forgiving nature","…is considerate and kind to almost everyone*"].

### Selection of variables

This section expands on statistical and methodological issues which are mentioned only briefly in the main paper.

The analysis principally is interested in factors affecting COVID-19 vaccine hesitancy, but hesitancy might be related to some or many of the 259 (i.e. 260-1) variables, with many variables likely to be correlated. Particularly as all measures were originally included in the questionnaire as being plausibly related to COVID-19 and its effects in HCWs. Formal modelling, as with path analysis, requires a clear causal [14] ordering of variables, but this study resembles many others for which, "it is increasingly common to deal with many candidate predictors, often with modest *a priori* information about their potential relevance" [15 16].

Variable selection methods, such as exploratory stepwise selection, which remove predictors not attaining particular significance levels, are problematic in conventional regression, particularly as the number of predictors increases [17]. LASSO methods are popular, but have the limitation of not providing satisfactory standard errors for fitted parameters [18] which is problematic when dealing with multiply imputed datasets.

A requirement of *islasso* is that all independent variables are standardized (i.e. have a mean of zero and a standard deviation of one), and it is also convenient for comparison purposes to have all measures, including dependent variables, standardized (and although COVID-19 vaccine hesitancy is binary, not all variables acting as potential dependent variables are binary). The *islasso* analyses therefore treat all dependent variables as gaussian, since there is no straightforward way of converting logistic regressions to conventional beta coefficients [19]. Although it is sometimes suggested that only logistic regressions should be used for binary variables, for binary variables with proportions from 0.1 to 0.9, such as COVID-19 vaccine hesitancy, standard linear regression gives very similar results to logistic regression [20-23]. Standardised (beta) linear regression coefficients are therefore used for comparison across all variables in regression analyses, and similarly Pearson correlations are used for comparison of associations between variables. Beta coefficients and Pearson correlations are conventional measures of effect size, conversion to Cohen's *d* or similar being straightforward [24 25], and odds ratios can be transformed in a similar way [26].

### Setting of a significance level

The conventional significance level of alpha=0.05 is problematic in several ways, particularly with very large sample sizes and multiple significance testing. Even with a small sample and a single test, for a result with important practical implications it is prudent to set alpha=.001. Sample size can be taken into account using the approach of Raftery [27 28], who for a typical small study with N=30, found that an alpha value of 0.001 was equivalent to a Bayes Factor (*BF*) of about 150, which was called 'Very Strong'. For an *N* of 12,431, as in the present study, an appropriate equivalent significance level for a *BF* of 150 would be $1.042 \times 10^{-5}$ (calculated as $t=sqrt(log_e(N)+2*log_e(BF)$, where $t$ is equivalent to a two-tailed $z$-statistic given the large *N*). For 259 separate predictor measures (i.e 260-1), each with its own significance test, a Bonferroni corrected significance level be $1.042 \times 10^{-5}/259 = 4.02 \times 10^{-8}$, which is the primary significance level used for analyses in the present study.

### Analysis of predictors of predictors.

If, in a regression, A, B and C are significant predictors of Y, then other variables such as D, E and F, despite showing simple relations to Y, may not be significant after A, B, and C are taken into account. D, E and F may however be indirect causes of Y, being mediated through A, B or C. Using *islasso* such situations can be modelled as a series of conventional path models [29-31], with previous predictors being set in turn as dependent variables and analysed using *islasso*. Dependent variables in those analyses were restricted to those which were not previously in the analysis to avoid problems of reciprocal causation [32]. For the path modelling, a separate Bonferroni corrected significance level is required. A full, non-recursive, path model can in principle be considered in LISREL terms as the lower triangle of the Beta matrix[33], with a saturated model having 259x258/2 =33,411 parameters, any of which in principle could be significant. A conservative significance level using a Bonferroni correction for variables in the path model is therefore $1.042 \times 10^{-5}/33411 = 3.11 \times 10^{-10}$, which is the level used in the path modelling. Although most variables were endogenous, a few, mostly demographic, were regarded as exogenous (see main Figure 3), and were not modelled further.

In principle the data could be analysed by a conventional structural equation model analysis such as that provided by LISREL [33] or *lavaan* [34]. The practical problems of so come from the large number of variables and the even larger number of potential paths between them, and the need for combining results across the various imputation sets. The reality, given the need to explore the present data with speed, was that a traditional approach to path analysis with successive regression analyses, was more straightforward.

At present the innovative *islasso* approach of Cilluffo and colleagues [35 36] applies only to a situation in which there is a single dependent variable, and therefore cannot be used for path analysis or structural equation modelling.

### Gaussian graphical models

In recent years there has been a growing interest in Gaussian Graphical Models (GGMs), which assess the partial correlation matrix of relationships within a large set of variables [37]. Partial correlations have the advantage that their estimated association takes into account all other measures in the analysis [38]. Partial correlations can readily be calculated from the inverse of a correlation or covariance matrix. GGMs can also be estimated by a variant of LASSO, the graphical LASSO, in which network components are either retained or else end up as zero [39], using the package *glasso* which is available in *R.* In practical terms, the *R* package *qgraph* is particularly useful, not only for including *glasso,* but also for fitting a range of graphical models for visualization of data [40].

Given the popularity of GGMs it may be asked why the present study has used a different approach, based on *islasso* and path modelling. GGMs have two problems, which are not entirely solved. Firstly, *glasso* models, as with LASSO models in general, do not allow for the calculation of standard errors of estimates, which makes it difficult to use them in the context of multiple imputation, although a partial but computationally intensive solution is available using bootstrap approaches [14]. Secondly, the networks estimated through *glasso* are undirected networks, so that although an association of A and B can be shown to non-zero, it is not straightforward to make a case for a directed network, so that A causes B rather than vice-versa. Path analysis solves that problem by allowing a priori, theoretical knowledge about the likely causal direction of effects to be modelled uses directed edges which are equivalent to the beta matrix of the LISREL model [31 33].

Despite the problems of fitting only an undirected network it is still of interest to fit a *glasso* model to the present data, not least to confirm that the broad principles found in the main paper, and particularly in its Figure 3, are reasonable descriptions of the data and still appear when fitted using *glasso*. Were the two approaches to be wildly discrepant then that would throw doubt on the approach of the main paper.

The *glasso* approach cannot be used on multiply imputed datasets in the usual way. Since the present analysis is only exploratory, a correlation matrix for the data was calculated by firstly stacking the 20 multiple imputations into a single data set with 12431 x 20 = 246,420 'cases', and then calculating the correlations between the entire set of 260 measures[b]. The correlation matrix from the stacked data was then input into the using the *EBICglasso* function in the *qgraph* package, with N set as 12,431 (i.e. the true number of cases not the spuriously large apparent number of 'cases' in the stacked data). Default parameters were used, with gamma=0.5. The network was then plotted using the *qgraph* function with the 'spring' option which implements the Fruchterman-Reingold algorithm [41]. Supplementary Figure 2, below, shows the fitted network with the 260 nodes, with positive (green) and negative (red) edges linking the nodes which are included in the model. The supplementary file *VariableSummaryForSupplementaryInformation.xlsx* shows the numerical codes used in Supplementary Figure 2 the figure. Boxes are coloured to help see the relationship to Figure 3 in the main paper, with the main dependent variable, Vaccine Hesitancy, node 250, being shown in black. Nodes included in the 2nd, 3rd, 4th, 5th and 6th circles of the main Figure 3 are shown in red, orange, yellow, green and blue, and nodes not included in main Figure 3 are shown as white boxes

[b] This approach was adopted because analyses of a single imputation dataset gave occasional errors due to zero variance, presumably because of small proportions occasionally giving zero cases and hence zero variance for some measures. There were no such problems with the stacked dataset.

Supplementary Figure 2 is clearly complex. There are 849 edges connecting the 260 nodes, and visually complex, that network is relatively sparse, only 2.52% of the possible 260x259/2 = 33,670 edges being present. Only 6 of the 260 nodes are unconnected to other nodes, and apart from one isolated set of 3 nodes connected only to each other (122, 123, 124) all of the remaining 251 nodes are interconnected, albeit in some cases quite distantly. The was not an unexpected result given the nature of the cognitive-socio-behavioural measures in the phenomic dataset.

On a qualitative basis it can be seen that vaccine hesitancy, node 250 in black, is relatively close to most of the red, orange and yellow nodes which are circles 2, 3 and 4 of the main figure 3.

More specifically, Vaccine Hesitancy, has direct connections to six other nodes, two positive, to Covid-19 Conspiracy Beliefs, and being Pregnant, and four negative, being Older, being Pro Vaccines in general, having had a Flu vaccine, and having received the January version of the vaccination question. All six of these are in main Figure 3, with only Optimism about the Vaccination programme from main Figure 3 being missing from the *glasso* model.  The path model and *glasso* are therefore giving very similar results.

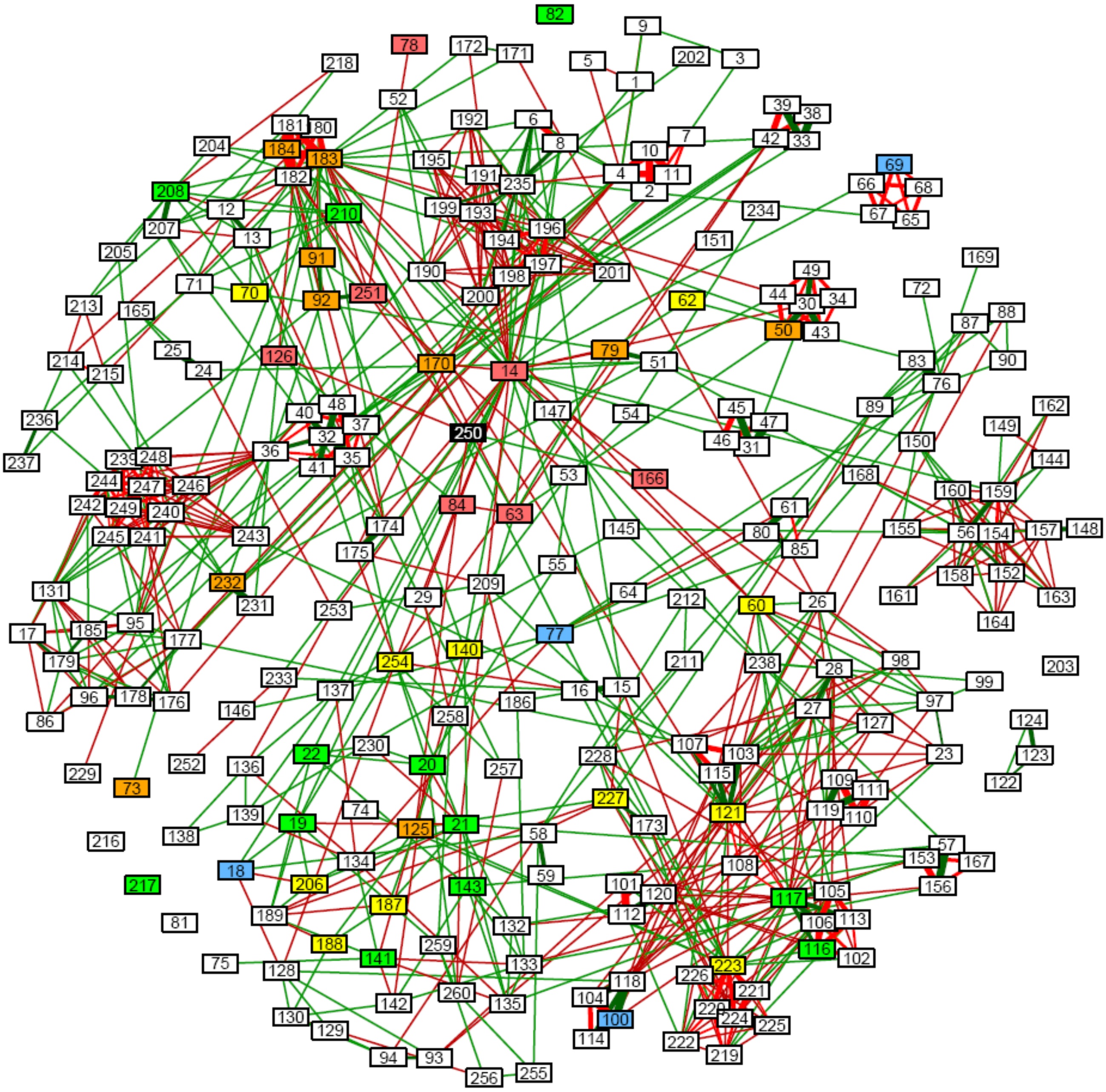

*Supplementary Figure 2: glasso model for all 260 variables. See text for details.*

As a second comparison, the seven edges connected to Fatalism (node 125 in orange, located south by west of Vaccine Hesitancy in black) were explored, six of which were positive (Covid-19 conspiracy beliefs, Chance Locus of Control and External Locus of Control, Need for Closure, being Muslim, and Religion being Important in Everyday Life, with the only negative edge being to Pro Vaccines in general. All seven of the edges to Fatalism are therefore identical to those in main Figure 3.

Without exploring further it seems clear that the path analysis approach, and the *glasso* method are giving very similar models, with the path analysis of main Figure 3 having the advantages a) of having taken the multiple imputations into account, and b) the causal direction being more secure as which variables are dependent is stated directly in the analyses. The comparison with the *glasso* model also strongly suggests that the significance levels adopted in the path analyses are broadly correct, giving similar numbers of paths/edges in the two analyses.

Supplementary Figure 2 is complex, and therefore Supplementary Figure 3 shows a simpler version, the only variables included being those which were also present in the main Figure 3.

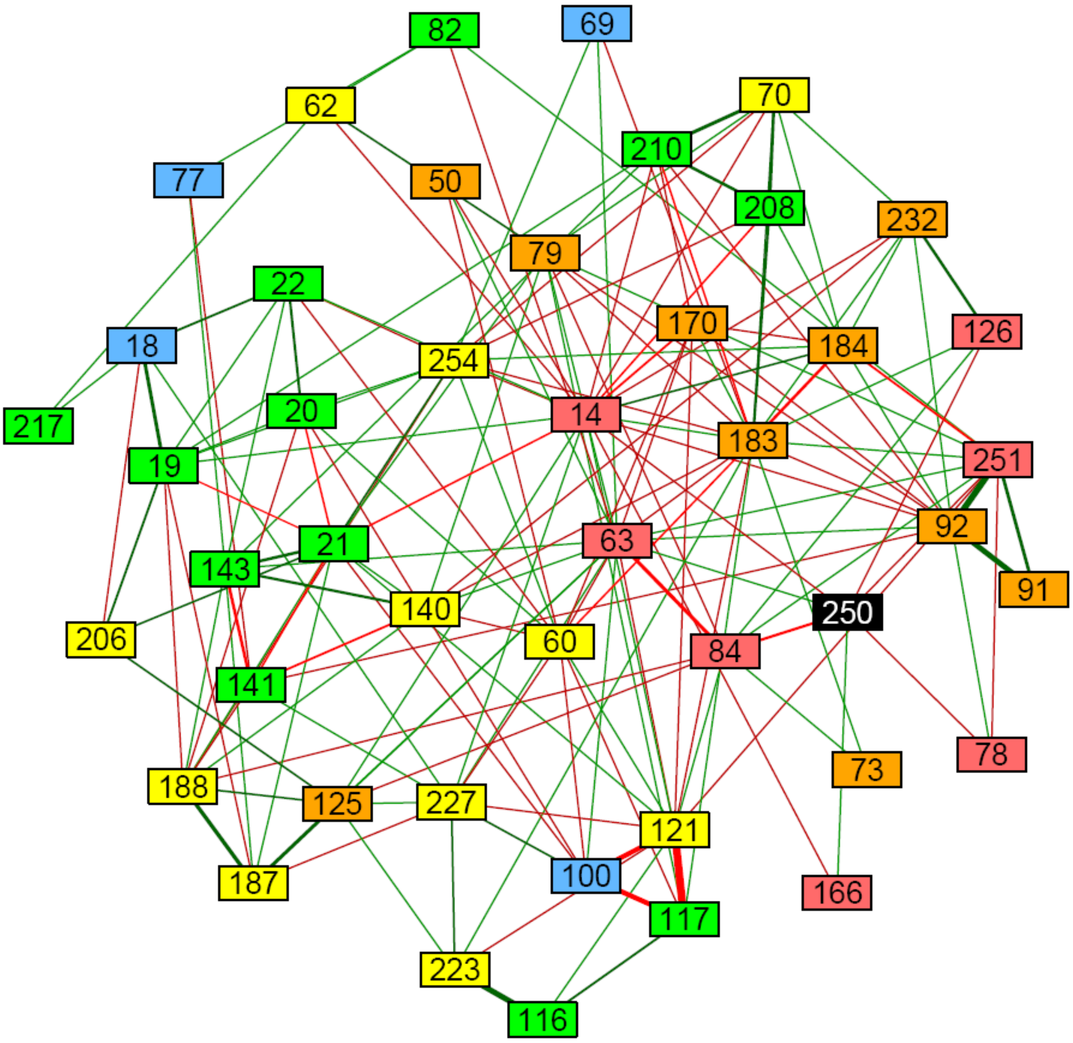

*Supplementary Figure 3: glasso model fitted to the variables included in Main Figure 3.*

The most important feature of Supplementary Figure 3 is that the red and orange nodes, from circles 2 and 3 of main figure 3, are visually closer to Vaccine Hesitancy, in black, than are the nodes from the outer circles in yellow, green and blue, reflecting the broad structure of Main Figure 3.

Without going into more details, it seems clear that the path analysis method in the main paper is giving broadly equivalent results to the very different approach using *glasso* to fit Gaussian Graphical Models, which is reassuring. The main Figure 3 can therefore be trusted as a representation of the complex influences on vaccine hesitancy, and on the causes of the causes of vaccine hesitancy.

*Supplementary Table 2: Demographic measures in relation to Vaccine Hesitancy*

| | SARS-Cov2-2-vaccine | | |
|---|---|---|---|
| **Variable** | **Not Hesitant 9325 (74.8%)** | **Hesistant 2915 (23.4%)** | **Missing 220 (1.7%)** |
| **Age (Median; IQR)** | 46 (36-55) | 39 (31-50) | 42 (33-53) |
| **Sex (n, %)** | | | |
| Male | 2435 (81.8%) | 500 (16.8%) | 40 (1.4%) |
| Female | 6875 (72.7%) | 2405 (25.4%) | 175 (1.8%) |
| **Ethnicity (n, %)** | | | |
| Asian - Pakistani | 230 (66.9%) | 105 (30.8%) | 10 (#) |
| Asian - Bangladeshi | 60 (78.7%) | 15 (#) | 0 (#) |
| Asian - Chinese | 170 (64.2%) | 90 (34.0%) | 5 (#) |
| Asian - Other Asian | 295 (73.9%) | 95 (24.4%) | 5 (#) |
| Black - African | 225 (59.3%) | 130 (34.9%) | 20 (#) |
| Black - Caribbean | 50 (43.6%) | 60 (50.4%) | 5 (#) |
| Black - Other Black | 15 (#) | 10 (#) | 5 (#) |
| Mixed - White and Black Caribbean | 55 (61.4%) | 30 (35.2%) | 5 (#) |
| Mixed - White and Black African | 50 (72.9%) | 20 (#) | 0 (#) |
| Mixed - White and Asian | 145 (75.4%) | 45 (23.6%) | 0 (#) |
| Mixed - Other | 115 (72.2%) | 40 (25.3%) | 5 (#) |
| White - British | 5770 (77.7%) | 1565 (21.1%) | 95 (1.3%) |
| White - Irish | 170 (75.4%) | 55 (23.2%) | 5 (#) |
| White - Other | 640 (68.4%) | 270 (29.0%) | 25 (2.6%) |
| Other - Arab | 85 (68.0%) | 35 (28.1%) | 5 (#) |
| Other - Any other | 95 (71.2%) | 35 (28.0%) | 0 (#) |
| **Religion (n, %)** | | | |
| None | 3200 (75.8%) | 970 (23.0%) | 50 (1.2%) |
| Buddhist | 95 (69.6%) | 35 (27.4%) | 5 (#) |
| Hindu | 585 (79.7%) | 140 (19.2%) | 10 (#) |
| Jewish | 95 (85.0%) | 15 (#) | 0 (#) |
| Muslim | 500 (68.9%) | 210 (28.6%) | 20 (#) |
| Sikh | 100 (73.0%) | 35 (24.8%) | 5 (#) |
| Other | 105 (63.7%) | 60 (35.1%) | 0 (#) |
| **Country of birth (n, %)** | | | |
| Not born in UK | 2405 (72.7%) | 820 (24.8%) | 80 (2.5%) |
| Born in UK | 6900 (75.6%) | 2085 (22.9%) | 135 (1.5%) |
| **Job role (n, %)** | | | |
| Doctors and Medical Support | 2270 (80.8%) | 515 (18.4%) | 25 (0.8%) |
| Nurses, NAs, Midwives | 1770 (69.4%) | 720 (28.2%) | 60 (2.4%) |
| Allied Health Professionals | 3875 (74.3%) | 1250 (24.0%) | 90 (1.8%) |
| Dental | 585 (77.3%) | 155 (20.7%) | 15 (#) |
| Administrative/Estates/Other | 485 (74.6%) | 155 (23.5%) | 10 (#) |
| **Job location (n, %)** | | | |
| Not in hospital | 4125 (77.0%) | 1140 (21.3%) | 90 (1.7%) |
| Hospital | 4955 (73.1%) | 1705 (25.2%) | 115 (1.7%) |
| **IMD quintile (n, %)** | | | |
| 1 (most deprived) | 735 (67.4%) | 325 (29.9%) | 30 (2.7%) |
| 2 | 1280 (70.1%) | 510 (28.0%) | 35 (2.0%) |
| 3 | 1700 (74.7%) | 525 (23.1%) | 50 (2.1%) |
| 4 | 2050 (76.6%) | 570 (21.4%) | 55 (2.0%) |
| 5 (Least deprived) | 2485 (78.5%) | 650 (20.6%) | 30 (1.0%) |

Note: All integer counts rounded to the nearest 5; # indicates percentages suppressed when N< 22.5.

### Demographic measures by vaccine hesitancy

Supplementary Table 2 is equivalent to Table 2 in the previous, interim, analysis [1] but has been updated to include the final sample used in the current study. Note that all sample sizes have been adjusted following the method described by HESA for statistical disclosure limitation[c] by a) rounding all Ns to the nearest 5, and b) suppressing all percentages where N is less than 22.5 (indicated by #).

### Assessing the extent of response bias

Our previous, interim, report [1] provided information on the extent of response bias in UK-REACH and for convenience the section on bias is reprinted below. The section is however updated by including a major multivariate analysis for respondents who took part via the GMC website, along with other minor modifications and updatings.

The UK-REACH (UK Research Study Into Ethnicity and COVID-19 Outcomes) questionnaire studying Covid-19 in UK Health Care Workers (HCWs) with a particular interest in HCWs from ethnic minorities, was distributed from 4/12/2020 to 2/3/2021 by Healthcare Regulatory bodies and some NHS Trusts, with reminders sent on several occasions. Invitees were sent a regulator-specific response address, but responses could also be made directly to the UK-REACH website. It should be emphasized that none of the questionnaire data were available to regulators, trusts or employers.

The total number of HCWs contacted overall is not easy to estimate. About 1.07m individuals were contacted by the seven main regulatory bodies (GPhC, GMC, PSNI, GOC, NMC, HCPC and GDC), with much smaller numbers contacted by 29 employers, unions and trusts. Emails from regulators are not always opened, and 'open rates' provided by four regulators were 54%, 48%, 47% and 40%, which are typical of mass mailings even from professional bodies.

Response rates for contacts with UK-REACH included 25,784 individuals who 'registered', 17,902 who 'consented', 15,592 who began answering the survey, and 12,431 who signed off the questionnaire as completed. The latter group answered the majority of questions (4.6% missing data), whereas for the group who started but did not complete the questionnaire, (15,592-12,431= 3,161), 76% of data were missing.

Overall response rates are low, but sadly are typical of most studies relying on bulk emailing, particularly as here where there is no personal contact of the research team with those contacted, because of data protection and ethics concerns on the part of the regulators and employers. For most of the present analyses it is only the 12,431 HCWs who completed and signed off the questionnaire who could be analysed, making the overall response rate of the order of 12,431/1.2m ≈ 1%. Nevertheless the number of respondents with usable data, 12,431, is substantial, which is reassuring. Estimated response rates, based on the 17,902 who consented, differed between regulators, being highest for the GMC (3.44%), followed by the GOC (1.72%), HCPC (1.65%), GPhC (1.11%), GDC (0.67%) and NMC (0.48%)[d].

A key question is the extent to which the 12,421 respondents who completed are a representative subset of HCWs in general, or perhaps are biased to various sub-groups. Unfortunately, most regulators and trusts were unable to provide detailed statistics on the demographics of those to whom they had sent invitations. An additional problem with all regulators is that it is not possible to link data from regulators at the individual level with data returned by individuals in the UK-REACH questionnaire, making it hard overall to estimate response biases. It was however possible to make more detailed comparison for doctors who were contacted via the GMC, as will be described later.

[c] https://www.hesa.ac.uk/about/regulation/data-protection/rounding-and-suppression-anonymise-statistics
[d] Note that although numerators are accurate, denominators are not always precise. PSNI is not reported as the denominator was below 10,000.

### Comparison with NHS employees.

Finding a group with whom to compare the UK-REACH respondents is not straightforward. HCWs are employed in the NHS, but there are many other workers in the NHS, and likewise there are many HCWs who work outside of the NHS. In the absence of a formal sampling frame, the best that seems possible is to compare the sample with NHS employees in general. Supplementary Table 3, as a broad comparator, shows the gender, age and ethnicity of NHS employees and of UK-REACH respondents.

*Supplementary Table 3: Gender, age and ethnicity of respondents and NHS staff.*

| | | NHS (2018/19) | UK-REACH n=12,431 |
|---|---|---|---|
| **Gender**[e] | | | |
| | Female | 77% | 76.0% |
| | Male | 23% | 24.0% |
| **Age**[f] | | | |
| | <25 | 6% | 2.9% |
| | 25-34 | 23% | 22.8% |
| | 35-44 | 24% | 23.8% |
| | 45-54 | 29% | 26.7% |
| | 55-64 | 17% | 20.2% |
| | 65+ | 2% | 3.7% |
| **Ethnicity**[g] | | | |
| | Asian | 10% | 18.9% |
| | Black | 6% | 4.3% |
| | Mixed | 2% | 4.2% |
| | White | 76% | 69.0% |
| | Other | 2% | 2.1% |
| | NK | 5% | 1.7% |

The UK-REACH respondents have a similar gender mix as NHS employees, and are broadly similar in age, with slightly more older respondents, which might be expected of those on professional registers. The ethnicity mix is not the same, with fewer Whites, and more Asian and Mixed participants. However the survey included ethnicity in its name, some of the requests to take part specifically addressed ethnic minority HCWs, and the invitations to some groups of respondents, the GMC in particular, were over-sampled for ethnicity. The lower rate of responses from Whites is therefore a mark of success for the sampling strategy of UK-REACH.

### Response rates in doctors.

Estimates of responses biases are possible to some extent using data provided by the General Medical Council (GMC). The GMC sent out 100,000 invitations to take part in the UK-REACH study, with ethnic minorities intentionally oversampled, receiving about two thirds of invitations. Although data could not be linked at the individual level, the GMC provided us with an anonymized data file containing basic demographic and professional data for the 100,000 doctors to whom invitations were sent. The data were deposited in the HIC Safe Haven at the University of Dundee, and were only accessible by one of us (ICM).

Of the 12,431 respondents who signed off the questionnaire, 3,865 were known to be doctors, but of these only 2,945 had replied through a specific link for GMC respondents. The remaining 920 doctors had replied through a generic link on the UK-REACH website, and may have included doctors other than the 100,000 directly invited by the GMC. The remaining analyses are therefore restricted

[e] https://www.nhsemployers.org/2019/05/gender-in-the-nhs-infographic

[f] https://www.nhsemployers.org/case-studies-and-resources/2019/05/age-in-the-nhs-infographic

[g] https://www.nhsemployers.org/case-studies-and-resources/2019/05/age-in-the-nhs-infographic

Note that the infographic separates Asian and Chinese where they are grouped together in ONS5, and have been grouped together here as well to help comparison.

to the 2,945 doctors who definitely had received invitations via the GMC. Raw data were processed in SPSS, and logistic regressions carried out using the *glm*() function in *R.*

Supplementary Table 4 summarises data on five demographic measures.

> *Sex***.** Coded by the GMC as Man or Woman, corresponding to Male or Female. A similar classification was used in UK-REACH.
>
> *UK graduate*. Based on the place of Primary Medical Qualification (PMQ) on the GMC's LRMP (List of Registered Medical Practitioners), with a similar question asked in UK-REACH
>
> *GP Register*. The GMC provided information on whether doctors were on the GP Register or on the Specialist Register. In UK-REACH information was based on any of three questions where respondents replied that they were a GP or in GP training[h]. The UK-REACH study by accident omitted to ask whether a doctor was on the GMC's Specialist Register.
>
> *Year of Birth*. The GMC provided data on year of birth in decades which we grouped as <= 1959, 1960-69, 1970-79, 1980-89 and 1990+. Age in years in UK-REACH was provided at the day of consent, and was regarded as being at 1/1/2021 for calculating year of birth.
>
> *Ethnicity*. UK-REACH used the question and the standard 18 categories used by the UK's Office for National Statistics (ONS) for the 2011 Census. As well as the ONS18 categories, we also used the five categories used by ONS (ONS5), and a binary measure of minority ethnicity (white vs BAME (Black and Minority Ethnic)). The GMC has used somewhat different measures at different times and they adjusted their categories to match ONS18.

GMC data were complete for Sex, UK graduate, and GP Register, but there were some missing data for Year of Birth and Ethnicity. UK-REACH data were missing for some of the measures, and all analyses are reported only for those who provided answers, and since proportions of missing data are low, statistical comparisons used listwise deletion. Standard reporting restrictions (see earlier) were applied with all whole numbers rounded to the nearest five, and percentages and numbers omitted where N<22.5.

#### Univariate analyses of response biases in doctors.

Univariate analyses are shown in Supplementary Table 4 with odds ratio, confidence intervals and significance levels. Questionnaire respondents were more likely to be female (1.30x), less likely to be non-UK graduates (0.737x), and more likely to be on the GP Register (1.302x). Compared with doctors born in 1980-89, respondents were much more likely to be born before 1960 (2.364x), from 1960-69 (1.573x), and 1970-79 (1.187x), as well as slightly more likely to be born after 1989 (1.149x).

Ethnicity related to response rates. Overall, respondents were relatively less likely to be BAME (0.800x), although of course because of over-sampling in absolute terms there were more BAME than White doctors in the UK-REACH study (65.0% vs 35.0%). On the ONS5 grouping, relative to White respondents there were fewer Asian (.789x), Black (.726x) and Other (0.653x) respondents, but there were significantly more respondents of mixed ethnicity (1.326x).

On the ONS18 categories, data are not reported for four groups because of small numbers of UK-REACH respondents. Response rates in all groups are compared with the reference group of White British respondents. Amongst Asian respondents, Indian (.832x), Pakistani (.556x), Bangladeshi (.885x) and Other groups were less likely than Whites to respond, but Chinese showed no significant difference in response rate (1.085x). In the Black group, Caribbean and Other groups showed too few respondents to be reported, but Black African (0.679x) were less likely to respond than White British. Amongst the Mixed groups, White/Caribbean and White/African had too few respondents to be reported. However a higher response rate was found in the White/Asian (1.821x) group, with no significant difference for Other Mixed groups (0.902x). Amongst White groups, there were too few respondents to report results for Irish and Gypsy/Traveller groups. There was no significant difference from White British for the White Other group (.933x). Amongst the two 'Other' groups,

[h] Note that the GMC's GP Register only includes doctors who are registered as GPs and not those in training as GPs, whereas UK-REACH included GPs in training, and did not ask specifically about being on the GP Register.

response rates were not significantly different for the Arab group (.922x), but were substantially lower in the Other sub-group of Other (0.373x).

*Supplementary Table 4: Univariate analyses of demographics of GMC respondents*

| | | | GMC sampling frame N=100,000 | | | UK-REACH GMC respondents N=2,945 | | Univariate logistic analyses of response rate | | |
|---|---|---|---|---|---|---|---|---|---|---|
| | | | N | % | % ExcNK | N | % | Odds Ratio | 95% CI | p |
| Sex | | Male | 53585 | 53.6% | 53.6% | 1305 | 47.4% | 1 # | - | - |
| | | Female | 46415 | 46.4% | 46.4% | 1450 | 52.6% | 1.303 | (1.207 - 1.405) | 8.5E-12 |
| UK graduate | | Yes | 53875 | 53.9% | 53.9% | 850 | 61.0% | 1 # | - | - |
| | | No | 46125 | 46.10% | 46.10% | 1330 | 39.0% | 0.737 | (.675-.804) | 6.2E-12 |
| On GP Register | | No | 78400 | 78.40% | 78.40% | 2200 | 74.7% | 1 # | - | - |
| | | Yes | 21600 | 21.60% | 21.60% | 745 | 25.3% | 1.302 | (1.207 - 1.405) | 8.5E-12 |
| YearOfBirth | | <=1959 | 5490 | 5.5% | 5.6% | 295 | 10.1% | 2.364 | (2.063-2.710) | 3.3E-35 |
| | | 1960-69 | 15645 | 15.60% | 15.80% | 580 | 19.8% | 1.573 | (1.412 - 1.752) | 1.8E-16 |
| | | 1970-79 | 26925 | 26.90% | 27.20% | 765 | 26.2% | 1.187 | (1.074 - 1.311) | 0.00074 |
| | | 1980-89 | 34190 | 34.2% | 34.6% | 830 | 28.2% | 1 # | - | - |
| | | >=1990 | 16685 | 16.7% | 16.9% | 460 | 15.7% | 1.149 | (1.024 - 1.290) | 0.0183 |
| | | NK | 1060 | 1.1% | | | | | | |
| Ethnic Minority | | No | 29185 | 29.2% | 30.4% | 815 | 35.0% | 1 # | - | - |
| | | Yes | 66850 | 66.9% | 69.6% | 1510 | 65.0% | 0.8 | (.734 - .872) | 4.31E-07 |
| | | N/K | 3965 | 3.9% | | | | | | |
| ONS5 | | | | | | | | | | |
| | | Asian | 46760 | 46.8% | 48.7% | 815 | 44.9% | 0.789 | (.719 - .866) | 6.18E-07 |
| | | Black | 8610 | 8.6% | 9.0% | 1040 | 7.6% | 0.726 | (.616 - .855) | 0.000133 |
| | | Mixed | 4320 | 4.3% | 4.5% | 175 | 6.8% | 1.326 | (1.114 - 1.578) | 0.00148 |
| | | White | 29185 | 29.2% | 30.4% | 135 | 35.0% | 1 # | - | - |
| | | Other | 7155 | 7.2% | 7.5% | 155 | 5.7% | 0.653 | (.543 - .786) | 0.00000666 |
| | | NK | 3965 | 4.0% | - | | | | | |
| ONS18 | | | | | | | | | | |
| | Asian | Indian | 23760 | 23.8% | 24.7% | 570 | 24.6% | 0.832 | (.743 - .932) | 0.00156 |
| | | Pakistani | 10590 | 10.6% | 11.0% | 175 | 7.4% | 0.556 | (.469 - .658) | 1.04E-11 |
| | | Bangladeshi | 1605 | 1.6% | 1.7% | 40 | 1.8% | 0.885 | (.642 - 1.218) | 0.45 |
| | | Chinese | 3620 | 3.6% | 3.8% | 110 | 4.8% | 1.085 | (.885 - 1.330) | 0.434 |
| | | Other | 7185 | 7.2% | 7.5% | 145 | 6.2% | 0.687 | (.572 - .824) | 0.0000552 |
| | Black | African | 7820 | 7.8% | 8.1% | 155 | 0.067 | 0.679 | (.569 - .811) | 0.0000184 |
| | | Caribbean | 500 | 0.5% | 0.5% | 15 | 0.7% | * | * | * |
| | | Other | 290 | 0.3% | 0.3% | 5 | 0.3% | * | * | * |
| | Mixed | White&Carib | 270 | 0.3% | 0.3% | 10 | 0.4% | * | * | * |
| | | White&African | 635 | 0.6% | 0.7% | 20 | 0.9% | * | * | * |
| | | White&Asian | 1605 | 1.6% | 1.7% | 80 | 34.0% | 1.821 | (1.434 - 2.310) | 0.00000083 |
| | | Other | 1810 | 1.8% | 1.9% | 45 | 2.0% | 0.902 | (.668 - 1.217) | 0.501 |
| | White | E/W/S/NI | 22720 | 22.7% | 23.7% | 650 | 2.8% | 1 # | - | - |
| | | Irish | 1280 | 1.3% | 1.3% | 25 | 1.0% | * | * | * |
| | | Gypsy/Trav | 0 | * | * | 0 | * | * | * | * |
| | | Other | 5180 | 5.2% | 5.4% | 140 | 6.0% | 0.933 | (.775 - 1.124) | 0.462 |
| | Other | Arab | 3465 | 3.5% | 3.6% | 90 | 4.0% | 0.922 | (.740 - 1.515) | 0.477 |
| | Other | Other | 3695 | 3.7% | 3.8% | 40 | 1.8% | 0.373 | (.272 - .513) | 1.22E-09 |
| | | NK | 3965 | 0.04 | | | | | | |

*Notes: # Reference category; ExcNK: Excluding not known;*
*All Ns are rounded to the nearest 5; percentages and odds ratios omitted when N<22.5, indicated by *.*

Overall the univariate analyses of demography in the UK-REACH participants is broadly similar to that of NHS employees, but with an excess of non-white participants, which was intentional both in the way that the study was advertised, and in the recruitment letters sent out by the GMC to would-be participants. However, there was some evidence for small amounts of response bias, with a relative excess of females, older participants, GPs, and white respondents, all of whom were somewhat more likely to respond to requests to participate.

Multivariate analyses of response biases in doctors. The univariate analyses can only consider biases separately for the various measures, but cannot ask whether effects are independent, or if some measures are confounded, or whether there are interactions between the measures in their effects on bias. Those problems can be solved by a more extensive analysis.

A practical problem for analyzing the data is that the GMC, for reasons of ethics and confidentiality, could only provide us with de-identified data on the 100,000 doctors to whom requests had been sent for participation in the questionnaire, but were willing to provide those anonymised data within the HIC Trusted Research Environment (Safe Haven) in Dundee[i]. Row-by-row data were available for the 100,000 doctors who had been sent requests to participate in the study. Five variables were provided: year of birth in decades; ethnicity on the ONS18 scale; place of primary medical qualification (PMQ: UK or non-UK); sex (Man or Woman); and whether or not the doctor was on the GP Register[j]. The ONS18 measure could be converted into the ONS5 scale, and also into a binary scale of whether a doctor was or was not from an ethnic minority (Ethnic minority scored 1 and 0).

A file from UK-REACH, which was anonymous and with no identifiers, was also ported into the HIC Safe Haven, with the help of Daniel Smith of the GMC. The file described 2919 doctors who had completed the UK-REACH questionnaire via the link indicating that they had received a request from the GMC[k]. The file had a similar set of five measures to those available for the GMC data (year of birth in decades; ONS18; PMQ; sex; and GP Register).

The two datasets, which we will refer to as the *sampling frame* (n=100,000) and the *respondents* (n=2,919) are not of independent, since the latter must be included within the former, although we were not informed which particular doctors in the sampling frame had responded. The original intention was to ignore the overlap and treat a combined file of 100,000+2,919= 102,919 individuals as being two independent datasets, which would have resulted in small statistical biases in the analyses due to the non-independence, but would still have given a reasonable overview. However, it was then realized that it was possible in most cases to remove the known respondents from the sampling frame. A six-digit code was created from the six measures (ONS18 (1 to 18)x10000 + decade of birth (4 to 9 x 1000), + sex (0 or 1 x 100) + PMQ (0 or 1 x 10) + GP Register (0 or 1). An *R* program then searched through the sampling frame looking for codes which were present in the respondents, and removing them[l]. Almost all were found[m]. A combined data set of respondents and non-respondents (i.e. sampling frame less known respondents) had 100,005 cases and was sufficiently accurate to be used for the rest of the analysis[n].

---

[i] https://www.dundee.ac.uk/hic/hic-trusted-research-environment/

[j] We were also provided with a binary variable indicating whether the doctor was on the specialist register or not, but we were unable to use that as we did not have equivalent data within UK-REACH.

[k] In other words, the doctors did not respond through the general UK-REACH website, when they may have been doctors who were additional to the GMC call, and were taking part through other routes. It should also be repeated that the GMC had no access to the UK-REACH data, their only involvement being in asking a stratified sample of doctors to take part.

[l] Lest it be unclear, the matching did not match a particular case, but if, say, a respondent was had a six-figure code 124101, a single case was found within the frame which also had the code 124010 and that single case was removed from the sampling frame. In most cases there are many potential such matches in the sampling frame but all are identical as far as this analysis is concerned, and therefore the first one was removed. The process then continues for the next case in the respondents, until all have had cases removed from the sampling frame. There is no direct matching of respondents to the GMC data, only matching by the five measures which were included.

[m] When there were no matches, the usual explanation was that the GP measure was not quite equivalent in the GMC and UK-REACH data, mostly because GP status was measured in a slightly different way in UK-REACH, GP trainees being included as GPs, and there being no explicit question on whether a doctor was on the GP Register. In the occasional cases of there being no match then the process was repeated but omitting the GP Register measure. If there was still no match then the ONS18 measure was replaced with the ONS5 measure. If no match was still found, then ONS5 was replaced with the binary ethnic minority measure.

[n] A more sophisticated approach could have carried out multiple imputations, as occasional variables were missing in both the GMC and UK-REACH data, but the present approach was probably sufficient for present purposes of obtaining a sense of whether there were biases, and how different measures may have interacted.

Ethnicity is coded in three separate ways, as a binary measure, a five-point categorical measure and an 18-point categorical measure. Year of birth was coded on a six-point scale. In addition, there are three other binary measures. A series of five models was fitted to the data, with Model 1 the simplest.

*Analysis of the binary ethnic minority measure*. Whether a doctor responded was the dependent variable in the logistic regression. Model 1 shows that respondents were less likely to be from an ethnic minority, more likely to be female, more likely to be UK graduates and more likely to be on the GP Register. Decade of birth showed no differences between older doctors born in the 1940s, 1950s and 1960s, but there was then a progressive decline in response rate for those born in the 1970s, the 1980s and from 1990 onwards. Model 2 is similar to Model 1 except that decade of birth, scored 4 = <1950, 5 = 1950s, 6=1960s, etc, was fitted as a linear effect, the linear effect being highly significant, with a decrease in odds ratio of .806x per decade. The effect of adding a quadratic effect was explored (not shown) and the significance of the quadratic effect only reached p=.031; it was therefore decided to ignore quadratic effects of decade of birth in further models. Model 3 explored interaction effects between ethnic minority and being a woman, a UK graduate, being on the GP register, and age; none were statistically significant. Overall therefore there were main effects shown in Model 1 and 2, but no evidence of statistical interactions in response rates.

Respondents are therefore less likely to be from an ethnic minority, but respondents are also independently more likely to be female, UK graduates, and on the GP Register, as shown in Models 1 and 2.

*Supplementary Table 5: Multivariate analyses of response to GMC request: Models 1, 2 and 3*

| | Model 1 | Model 2 | Model 3 |
|---|---|---|---|
| Ethnic minority (BAME) | **b= -.146 OR= .864**** | **b= -.156 OR= .856 ***** | b= -.292 OR=.107 NS |
| Woman | **b= .370 OR= 1.448 ***** | **b= .362 OR= 1.436 ***** | **b= .412 OR= 1.510 ***** |
| UK graduate | **b= .407 OR= 1.503***** | **b= .425 OR= 1.530 ***** | **b= .452 OR= 1.571 ***** |
| On GP register | **b=.138 OR= 1.149 **** | **b= .095 1.099 *** | b= .060 OR= 1.062 NS |
| Decade of Birth <1950 | 0 (ref) OR=1 | - | - |
| Decade of Birth 1950-59 | b= .147 OR= 1.158 NS | - | - |
| Decade of Birth 1960-69 | b= .025 OR= NS | - | - |
| Decade of Birth 1970-79 | **b= -.362 OR= .696 *** | - | - |
| Decade of Birth 1980-89 | **b= -.681 OR= .506 ***** | - | - |
| Decade of Birth 1990+ | **b= -.532 OR= .587 **** | - | - |
| Decade of Birth (linear) | - | **b= -.261 OR= .806 ***** | **b= -.232 OR= .793 ***** |
| Ethnic minority x Woman | - | - | b= -.904 OR= .923 NS |
| Ethnic minority x UK graduate | - | - | b= -.030 OR= .961 NS |
| Ethnic minority x On GP register | - | - | b=.041 OR= 1.062 NS |
| Ethnic minority x Decade of Birth (lin) | - | - | b= .027 OR= 1.028 NS |
| Intercept | b= -3.051 OR= 0.030 | b= -2.303 OR= .100 | b= -2.347 OR= .107 |
| AIC | 24478 | 24518 | 24525 |

*Notes: NS: p<.05; * P<.05; ** p<.01; *** p<.001; negative coefficients shown in red and significant coefficients (p<.05) shown in bold*

*Analysis of the ONS and ONS18 measures of ethnicity.* The multivariate analysis so far has considered ethnicity only as a binary measure, comparing ethnic minority doctors with other doctors. The remainder of the analysis will consider finer-grained measures of ethnicity using the ONS5 and ONS18 scales, which are shown in Model 4 and Model 5 of Supplementary table 6.

*Supplementary Table 6: Multivariate analyses of response to GMC request: Models 4 and 5*

| | Model 4 | Model 5 |
|---|---|---|
| Woman | **b= .362 (SE .041) OR= 1.436 ***** | **b= .359 (SE .041) OR= 1.432 ***** |
| UK graduate | **b= .416 (SE .047) OR= 1.516 ***** | **b= .339 (SE .053) OR= 1.404 ***** |
| On GP register | **b= .098 (SE .046) OR= 1.103 *** | **b= .115 (SE .046) OR= 1.122 *** |
| Decade of Birth (linear) | **b= -.217 (SE .018) OR= .805 ***** | **b= -.216 (SE .018) OR= .806 ***** |
| **ONS5: White** | 0 (ref) OR=1 | - |
| ONS18: British | - | 0 (ref) OR=1 |
| ONS18: Irish | - | **b= -.487 (SE .195) OR= .614 *** |
| ONS18: Traveller | - | **b= 3.101 (SE 1.429) OR= 22.24 *** |
| ONS18: Other | - | b= -.010 (SE .094) OR= .990 NS |
| **ONS5: Asian** | **b= -.179 (SE .047) OR= .836 ***** | - |
| ONS18: Indian | - | **b= -.164 (SE .061) OR= .848 **** |
| ONS18: Pakistani | - | **b= -.583 (SE .090) OR= .558 ***** |
| ONS18: Bangladeshi | - | b= -.040 (SE .154) OR=.961 NS |
| ONS18: Chinese | - | b= .148 (SE .095) OR= 1.159 NS |
| ONS18: Other Asian | - | **b= -.297 (SE .088) OR= .743 ***** |
| **ONS5: Black** | **b= -.188 (SE .084) OR= .828 *** | - |
| ONS18: African | - | **b= -.258 (SE .093) OR= .772 **** |
| ONS18: Caribbean | - | b=.011 (SE .250) OR= 1.011 NS |
| ONS18: Other Black | - | b= .538 (SE .415) OR= .584 NS |
| **ONS5: Mixed** | b= -.015 (SE .094 ) OR= .985 NS | - |
| ONS18: White & Black Caribbean | - | b= .252 (SE .311) OR= 1.286 NS |
| ONS18: White & Black African | - | b= .265 (SE .210) OR=1.303 NS |
| ONS18: White & Asian | - | **b= .496 (SE .117) OR= 1.641 ***** |
| ONS18: Other | - | b= -.093 (SE .144) OR= .911 NS |
| **ONS5: Other** | b= -.095 (SE .086) OR=.909 NS | - |
| ONS18: Arab | - | b= .045 (SE .121) OR= 1.046 NS |
| ONS18: Other | - | **b= -.999 (SE .160) OR= .368 ***** |
| Intercept | b= -2.293 (SE .123) OR= .101 | b= -2.216 (SE .126) OR= .109 |
| AIC | 24521 | 24415 |

*Notes: SE: Standard error; NS: p<.05; * P<.05; ** p<.01; *** p<.001;*
*negative coefficients shown in red and significant coefficients (p<.05) shown in bold*

Model 4 contains only main effects, and is equivalent to Model 2 in Supplementary Table 5, but with the binary ethnic minority variable replaced with the ONS5 categories. Compared with the reference (White) group, response rates are lower in Asian and Black doctors, but with no difference from Mixed or Other groups. Note that sample sizes in groups differ and therefore standard errors have been shown as an indicator of confidence intervals. Model 5 uses the ONS 18 categories where numbers vary even more between groups, with ONS18:Traveller group in particular having very small numbers of respondents and hence the SE being very wide. Within ONS5 groups (but the reference groups for all ONS18 categories being ONS18:White British, there are further differences in response rates. In the ONS5 group, Irish doctors are less likely to respond, and Travellers are more likely (but the standard error is very wide). In the ONS5 Asian group, Indian doctors are less likely to respond, as are Other Asian doctors, with Pakistani doctors being much less likely to respond, whereas Bangladeshi doctors showed no difference from the Reference Group. Within the ONS5: Black group, only African doctors are less likely to respond but standard errors are large in Other Black group. Within the ONS5:Mixed category, most groups are not significantly different from the Reference group, but Mixed White and Asian doctors are more likely to respond (than White British doctors). Within the ONS5:Other group, doctors of Arab background are equally likely to respond as with to White British Reference Group, but the ONS18:Other Other group are much less likely to respond.

Overall the details of Model 4 and particularly Model 5 show that although simple analyses of ethnic minority status (BAME) do show differences, a more fine-grained analysis using the ONS5 and particularly the ONS18 categories allows a far more nuanced analysis of ethnic differences. The differences between Indian, Bangladeshi and Pakistani in particular are of interest, as those groups are often merged into a 'South Asian' or similar group, but there are clear differences between them.

### Additional File

*Excel file VariableSummaryForSupplementaryInformation.xlsx .* A summary of all of the variables used in the analyses is available in the Excel file (see Supplementary Table 1 for details).

### Acknowledgments. We are grateful to Daniel Smith, Andrew Ledgard, Sue Carr, Amy Bloxham, and Ganesan Gurusamy of the GMC for their help in facilitating the provision of the GMC data, and porting the data files to the HIC Safe Haven.
