## Supplementary material for "Vaccine hesitancy for COVID-19 explored in a phenomic study of 259 socio-cognitive-behavioural measures in the UK-REACH study of 12,431 UK healthcare workers": PDF for looking at XLS file of variables

### This is a PDF of the proper spreadsheet as medRxiv will not allow uploading of .xlsx files.

| Order | Section | Raw | Derived | Marker | SPSSvariable | Impn | is.lasso | Figure3 | Fig 3 Label | PathLevel | Code | Type | Label |
|---|---|---|---|---|---|---|---|---|---|---|---|---|---|
| 0 | 1153 | 785 | 392 | 25 | COUNTS | 214 | 260 | 44 | | 260 | | | |
| 1 | 0 | 1 | 0 | 0 | DataFileName | | | | | | | String | Version of Redcap files that has been used |
| 2 | 0 | 1 | 0 | 0 | ID | | | | | | | Numeric | |
| 3 | 0 | 1 | 0 | 0 | ExtractionDate | | | | | | | String | |
| 4 | 0 | 1 | 0 | 0 | WorkingFileVersion | | | | | | | String | |
| 5 | 0 | 1 | 0 | 0 | WorkingFileDate | | | | | | | String | |
| 6 | 0 | 0 | 0 | 1 | Section0_Raw_RegistrationEtc______________Raw | | | | | | | Numeric | |
| 7 | 0 | 1 | 0 | 0 | ukreach_questionnaire_complete | | | | | | | Numeric | Complete? |
| 8 | 0 | 1 | 0 | 0 | regulator | | | | | | | String | |
| 9 | 0 | 1 | 0 | 0 | age_raw | | | | | | | Numeric | Age in years on date of registration [NB differs from 2021-01-19 version which used date of download] |
| 10 | 0 | 1 | 0 | 0 | date.of.consent | | | | | | | String | |
| 11 | 0 | 1 | 0 | 0 | date_quest_complete | | | | | | | String | |
| 12 | 0 | 1 | 0 | 0 | Index.of.Multiple.Deprivation.Decile | 1 | 1 | | | 7 | 179 | Numeric | |
| 13 | 0 | 1 | 0 | 0 | Income.Decile | 1 | 1 | | | 7 | 178 | Numeric | |
| 14 | 0 | 1 | 0 | 0 | Employment.Decile | 1 | 1 | | | 7 | 96 | Numeric | |
| 15 | 0 | 1 | 0 | 0 | Education.and.Skills.Decile | 1 | 1 | | | 7 | 95 | Numeric | |
| 16 | 0 | 1 | 0 | 0 | Health.and.Disability.Decile | 1 | 1 | | | 7 | 131 | Numeric | |
| 17 | 0 | 1 | 0 | 0 | Crime.Decile | 1 | 1 | | | 7 | 86 | Numeric | |
| 18 | 0 | 1 | 0 | 0 | Barriers.to.Housing.and.Services.Decile | 1 | 1 | | | 7 | 17 | Numeric | |
| 19 | 0 | 1 | 0 | 0 | Living.Environment.Decile | 1 | 1 | | | 7 | 185 | Numeric | |
| 20 | 0 | 1 | 0 | 0 | IDACI.Decile | 1 | 1 | | | 7 | 176 | Numeric | |
| 21 | 0 | 1 | 0 | 0 | IDAOPI.Decile | 1 | 1 | | | 7 | 177 | Numeric | |
| 22 | 1 | 0 | 0 | 1 | Section1_Raw_DemographicsEtc______________Raw | | | | | | | Numeric | |
| 23 | 1 | 1 | 0 | 0 | gender | | | | | | | Numeric | Which of the following best describes you? |
| 24 | 1 | 1 | 0 | 0 | gender_other | | | | | | | String | Please enter the term you use to describe your gender, or enter Prefer not to answer. |
| 25 | 1 | 1 | 0 | 0 | sex | | | | | | | Numeric | What was your sex assigned at birth? |
| 26 | 1 | 1 | 0 | 0 | rel | | | | | | | Numeric | Which of the following best describes your marital status?You will be asked more about who you liveÂ with laterÂ in the questionnaire. |
| 27 | 2 | 0 | 0 | 1 | Section2_Raw_Job______________Raw | | | | | | | Numeric | |
| 28 | 2 | 1 | 0 | 0 | jobrole | | | | | | | Numeric | What is your main job/role? Please choose the best fit and specify further if you wish.Â If you are not currently working, please answer about your most recent role. |
| 29 | 2 | 1 | 0 | 0 | jobrole_whc_other | | | | | | | String | Please specify your wider healthcare role: |
| 30 | 2 | 1 | 0 | 0 | jobrole_ahp_other | | | | | | | String | Please specify your Allied Health Professional role: |
| 31 | 2 | 1 | 0 | 0 | jobrole_amb_other | | | | | | | String | Please specify your ambulance role: |
| 32 | 2 | 1 | 0 | 0 | jobrole_css_other | | | | | | | String | Please specify your clinical support staff role: |
| 33 | 2 | 1 | 0 | 0 | jobrole_den_other | | | | | | | String | Please specify your dental role: |
| 34 | 2 | 1 | 0 | 0 | jobrole_ma_other | | | | | | | String | Please specify your medical associates role: |
| 35 | 2 | 1 | 0 | 0 | jobrole_nam_other | | | | | | | String | Please specify your nursing and midwifery role: |
| 36 | 2 | 1 | 0 | 0 | jobrole_pharm_other | | | | | | | String | Please specify your pharmacy role: |
| 37 | 2 | 1 | 0 | 0 | jobrole_opt_other | | | | | | | String | Please specify your optical role: |
| 38 | 2 | 1 | 0 | 0 | jobrole_other | | | | | | | String | Please specify your job role: |
| 39 | 2 | 1 | 0 | 0 | jobwork_now | | | | | | | Numeric | Now |
| 40 | 2 | 1 | 0 | 0 | jobwork_ld | | | | | | | Numeric | In the first month after the start of the UK national lockdown on 23rd March 2020 |
| 41 | 2 | 1 | 0 | 0 | jobnotwork_now___1 | | | | | | | Numeric | Please indicate the reason(s) you are not working now (Select all that apply): By shielding we mean taking extra steps to protect yourself, by minimising interactions between yourself and others because you are at high risk of severe illness from coronavir |
| 42 | 2 | 1 | 0 | 0 | jobnotwork_now___2 | | | | | | | Numeric | Please indicate the reason(s) you are not working now (Select all that apply): By shielding we mean taking extra steps to protect yourself, by minimising interactions between yourself and others because you are at high risk of severe illness from coronavir |
| 43 | 2 | 1 | 0 | 0 | jobnotwork_now___3 | | | | | | | Numeric | Please indicate the reason(s) you are not working now (Select all that apply): By shielding we mean taking extra steps to protect yourself, by minimising interactions between yourself and others because you are at high risk of severe illness from coronavir |
| 44 | 2 | 1 | 0 | 0 | jobnotwork_now___4 | | | | | | | Numeric | Please indicate the reason(s) you are not working now (Select all that apply): By shielding we mean taking extra steps to protect yourself, by minimising interactions between yourself and others because you are at high risk of severe illness from coronavir |
| 45 | 2 | 1 | 0 | 0 | jobnotwork_now___5 | | | | | | | Numeric | Please indicate the reason(s) you are not working now (Select all that apply): By shielding we mean taking extra steps to protect yourself, by minimising interactions between yourself and others because you are at high risk of severe illness from coronavir |
| 46 | 2 | 1 | 0 | 0 | jobnotwork_now___6 | | | | | | | Numeric | Please indicate the reason(s) you are not working now (Select all that apply): By shielding we mean taking extra steps to protect yourself, by minimising interactions between yourself and others because you are at high risk of severe illness from coronavir |
| 47 | 2 | 1 | 0 | 0 | jobnotwork_now___7 | | | | | | | Numeric | Please indicate the reason(s) you are not working now (Select all that apply): By shielding we mean taking extra steps to protect yourself, by minimising interactions between yourself and others because you are at high risk of severe illness from coronavir |
| 48 | 2 | 1 | 0 | 0 | jobnotwork_now___8 | | | | | | | Numeric | Please indicate the reason(s) you are not working now (Select all that apply): By shielding we mean taking extra steps to protect yourself, by minimising interactions between yourself and others because you are at high risk of severe illness from coronavir |
| 49 | 2 | 1 | 0 | 0 | jobnotwork_now___99 | | | | | | | Numeric | Please indicate the reason(s) you are not working now (Select all that apply): By shielding we mean taking extra steps to protect yourself, by minimising interactions between yourself and others because you are at high risk of severe illness from coronavir |
| 50 | 2 | 1 | 0 | 0 | jobnotwork_now_other | | | | | | | String | Please specify why you are not currently working: |
| 51 | 2 | 1 | 0 | 0 | jobnotwork_ld___1 | | | | | | | Numeric | Please indicate the reason(s) you were not working at the start of the UK national lockdown on 23rd March 2020 (select all that apply): By shielding we mean taking extra steps to protect yourself, by minimising interactions between yourself and others beca |
| 52 | 2 | 1 | 0 | 0 | jobnotwork_ld___2 | | | | | | | Numeric | Please indicate the reason(s) you were not working at the start of the UK national lockdown on 23rd March 2020 (select all that apply): By shielding we mean taking extra steps to protect yourself, by minimising interactions between yourself and others beca |
| 53 | 2 | 1 | 0 | 0 | jobnotwork_ld___3 | | | | | | | Numeric | Please indicate the reason(s) you were not working at the start of the UK national lockdown on 23rd March 2020 (select all that apply): By shielding we mean taking extra steps to protect yourself, by minimising interactions between yourself and others beca |
| 54 | 2 | 1 | 0 | 0 | jobnotwork_ld___4 | | | | | | | Numeric | Please indicate the reason(s) you were not working at the start of the UK national lockdown on 23rd March 2020 (select all that apply): By shielding we mean taking extra steps to protect yourself, by minimising interactions between yourself and others beca |
| 55 | 2 | 1 | 0 | 0 | jobnotwork_ld___5 | | | | | | | Numeric | Please indicate the reason(s) you were not working at the start of the UK national lockdown on 23rd March 2020 (select all that apply): By shielding we mean taking extra steps to protect yourself, by minimising interactions between yourself and others beca |
| 56 | 2 | 1 | 0 | 0 | jobnotwork_ld___6 | | | | | | | Numeric | Please indicate the reason(s) you were not working at the start of the UK national lockdown on 23rd March 2020 (select all that apply): By shielding we mean taking extra steps to protect yourself, by minimising interactions between yourself and others beca |
| 57 | 2 | 1 | 0 | 0 | jobnotwork_ld___7 | | | | | | | Numeric | Please indicate the reason(s) you were not working at the start of the UK national lockdown on 23rd March 2020 (select all that apply): By shielding we mean taking extra steps to protect yourself, by minimising interactions between yourself and others beca |
| 58 | 2 | 1 | 0 | 0 | jobnotwork_ld___8 | | | | | | | Numeric | Please indicate the reason(s) you were not working at the start of the UK national lockdown on 23rd March 2020 (select all that apply): By shielding we mean taking extra steps to protect yourself, by minimising interactions between yourself and others beca |
| 59 | 2 | 1 | 0 | 0 | jobnotwork_ld___99 | | | | | | | Numeric | Please indicate the reason(s) you were not working at the start of the UK national lockdown on 23rd March 2020 (select all that apply): By shielding we mean taking extra steps to protect yourself, by minimising interactions between yourself and others beca |

| | | | | | | | |
|---|---|---|---|---|---|---|---|
| 60 | 2 | 1 | 0 | 0 | **jobnotwork_ld_other** | String | Please specify why you were not working at the start of the UK national lockdown on 23rd March 2020: |
| 61 | 2 | 1 | 0 | 0 | **jobnhs___1** | Numeric | In which of the following sectors is your current main job/ role?If not currently working, please answer for your most recent main job/role.Select all that apply. (choice=NHS) |
| 62 | 2 | 1 | 0 | 0 | **jobnhs___2** | Numeric | In which of the following sectors is your current main job/ role?If not currently working, please answer for your most recent main job/role.Select all that apply. (choice=Other public sector (e.g. local or national government)) |
| 63 | 2 | 1 | 0 | 0 | **jobnhs___3** | Numeric | In which of the following sectors is your current main job/ role?If not currently working, please answer for your most recent main job/role.Select all that apply. (choice=Private sector) |
| 64 | 2 | 1 | 0 | 0 | **jobnhs___4** | Numeric | In which of the following sectors is your current main job/ role?If not currently working, please answer for your most recent main job/role.Select all that apply. (choice=Private facility temporarily used by the NHS) |
| 65 | 2 | 1 | 0 | 0 | **jobnhs___5** | Numeric | In which of the following sectors is your current main job/ role?If not currently working, please answer for your most recent main job/role.Select all that apply. (choice=University / higher education) |
| 66 | 2 | 1 | 0 | 0 | **jobnhs___99** | Numeric | In which of the following sectors is your current main job/ role?If not currently working, please answer for your most recent main job/role.Select all that apply. (choice=Prefer not to answer) |
| 67 | 2 | 1 | 0 | 0 | **job_pc** | String | It would be helpful to us to know where in the UK your main job/role is located. Please type the first part of the postcode (e.g. W1G, CF24, BT12 or EH16).Â If you cant remember the number at the end, just type the first letters (e.g. CF, BT or EH).If no |
| 68 | 2 | 1 | 0 | 0 | **jobnhs_ld___1** | Numeric | In which of the following sectors was your main job/role in the first month after the start of the UK national lockdown on 23rd March 2020?Select all that apply. (choice=NHS) |
| 69 | 2 | 1 | 0 | 0 | **jobnhs_ld___2** | Numeric | In which of the following sectors was your main job/role in the first month after the start of the UK national lockdown on 23rd March 2020?Select all that apply. (choice=Other public sector (e.g. local or national government)) |
| 70 | 2 | 1 | 0 | 0 | **jobnhs_ld___3** | Numeric | In which of the following sectors was your main job/role in the first month after the start of the UK national lockdown on 23rd March 2020?Select all that apply. (choice=Private sector) |
| 71 | 2 | 1 | 0 | 0 | **jobnhs_ld___4** | Numeric | In which of the following sectors was your main job/role in the first month after the start of the UK national lockdown on 23rd March 2020?Select all that apply. (choice=Private facility temporarily used by the NHS) |
| 72 | 2 | 1 | 0 | 0 | **jobnhs_ld___5** | Numeric | In which of the following sectors was your main job/role in the first month after the start of the UK national lockdown on 23rd March 2020?Select all that apply. (choice=University / higher education) |
| 73 | 2 | 1 | 0 | 0 | **jobnhs_ld___99** | Numeric | In which of the following sectors was your main job/role in the first month after the start of the UK national lockdown on 23rd March 2020?Select all that apply. (choice=Prefer not to answer) |
| 74 | 2 | 1 | 0 | 0 | **jobdrgrade** | Numeric | What is your current or most recent grade? |
| 75 | 2 | 1 | 0 | 0 | **jobdrgrade_other** | String | Please specify your current or most recent grade: |
| 76 | 2 | 1 | 0 | 0 | **jobdrgrade_ld** | Numeric | What was your grade at the start of the UK national lockdown on 23rd March 2020? |
| 77 | 2 | 1 | 0 | 0 | **jobdrgrade_other_2** | String | Please specify your grade: |
| 78 | 2 | 1 | 0 | 0 | **jobdrspec** | Numeric | What is your current or most recent specialty? |
| 79 | 2 | 1 | 0 | 0 | **jobdrspec_2** | Numeric | What was your specialty in the first month after the start of the UK national lockdown on 23rd March 2020? |
| 80 | 2 | 1 | 0 | 0 | **jobband** | Numeric | What is your current or most recent NHS band? |
| 81 | 2 | 1 | 0 | 0 | **jobband_2** | Numeric | What was your NHS band at the start of the UK national lockdown on 23rd March 2020? |
| 82 | 2 | 1 | 0 | 0 | **jobrnfield** | Numeric | What is your registered field of nursing? |
| 83 | 2 | 1 | 0 | 0 | **jobrnfield_other** | String | Please specify the two fields in which you practice: |
| 84 | 2 | 1 | 0 | 0 | **job_areas_1___1** | Numeric | Ambulance (inc air ambulance) (choice=Now) |
| 85 | 2 | 1 | 0 | 0 | **job_areas_1___2** | Numeric | Ambulance (inc air ambulance) (choice=UK national lockdown) |
| 86 | 2 | 1 | 0 | 0 | **job_areas_1___99** | Numeric | Ambulance (inc air ambulance) (choice=Prefer not to answer) |
| 87 | 2 | 1 | 0 | 0 | **job_areas_2___1** | Numeric | Armed forces (choice=Now) |
| 88 | 2 | 1 | 0 | 0 | **job_areas_2___2** | Numeric | Armed forces (choice=UK national lockdown) |
| 89 | 2 | 1 | 0 | 0 | **job_areas_2___99** | Numeric | Armed forces (choice=Prefer not to answer) |
| 90 | 2 | 1 | 0 | 0 | **job_areas_3___1** | Numeric | Community clinical / primary care setting (choice=Now) |
| 91 | 2 | 1 | 0 | 0 | **job_areas_3___2** | Numeric | Community clinical / primary care setting (choice=UK national lockdown) |
| 92 | 2 | 1 | 0 | 0 | **job_areas_3___99** | Numeric | Community clinical / primary care setting (choice=Prefer not to answer) |
| 93 | 2 | 1 | 0 | 0 | **job_areas_4___1** | Numeric | Community non-clinical settings (choice=Now) |
| 94 | 2 | 1 | 0 | 0 | **job_areas_4___2** | Numeric | Community non-clinical settings (choice=UK national lockdown) |
| 95 | 2 | 1 | 0 | 0 | **job_areas_4___99** | Numeric | Community non-clinical settings (choice=Prefer not to answer) |
| 96 | 2 | 1 | 0 | 0 | **job_areas_5___1** | Numeric | Hospitals - Emergency Department (choice=Now) |
| 97 | 2 | 1 | 0 | 0 | **job_areas_5___2** | Numeric | Hospitals - Emergency Department (choice=UK national lockdown) |
| 98 | 2 | 1 | 0 | 0 | **job_areas_5___99** | Numeric | Hospitals - Emergency Department (choice=Prefer not to answer) |
| 99 | 2 | 1 | 0 | 0 | **job_areas_6___1** | Numeric | Hospital - Intensive Care Unit (choice=Now) |
| 100 | 2 | 1 | 0 | 0 | **job_areas_6___2** | Numeric | Hospital - Intensive Care Unit (choice=UK national lockdown) |
| 101 | 2 | 1 | 0 | 0 | **job_areas_6___99** | Numeric | Hospital - Intensive Care Unit (choice=Prefer not to answer) |
| 102 | 2 | 1 | 0 | 0 | **job_areas_7___1** | Numeric | Hospital - other inpatient setting (choice=Now) |
| 103 | 2 | 1 | 0 | 0 | **job_areas_7___2** | Numeric | Hospital - other inpatient setting (choice=UK national lockdown) |
| 104 | 2 | 1 | 0 | 0 | **job_areas_7___99** | Numeric | Hospital - other inpatient setting (choice=Prefer not to answer) |
| 105 | 2 | 1 | 0 | 0 | **job_areas_8___1** | Numeric | Hospital - outpatients (choice=Now) |
| 106 | 2 | 1 | 0 | 0 | **job_areas_8___2** | Numeric | Hospital - outpatients (choice=UK national lockdown) |
| 107 | 2 | 1 | 0 | 0 | **job_areas_8___99** | Numeric | Hospital - outpatients (choice=Prefer not to answer) |
| 108 | 2 | 1 | 0 | 0 | **job_areas_9___1** | Numeric | Hospital - other clinical setting (choice=Now) |
| 109 | 2 | 1 | 0 | 0 | **job_areas_9___2** | Numeric | Hospital - other clinical setting (choice=UK national lockdown) |
| 110 | 2 | 1 | 0 | 0 | **job_areas_9___99** | Numeric | Hospital - other clinical setting (choice=Prefer not to answer) |
| 111 | 2 | 1 | 0 | 0 | **job_areas_10___1** | Numeric | Hospital - other non-clinical setting (choice=Now) |
| 112 | 2 | 1 | 0 | 0 | **job_areas_10___2** | Numeric | Hospital - other non-clinical setting (choice=UK national lockdown) |
| 113 | 2 | 1 | 0 | 0 | **job_areas_10___99** | Numeric | Hospital - other non-clinical setting (choice=Prefer not to answer) |
| 114 | 2 | 1 | 0 | 0 | **job_areas_11___1** | Numeric | Hospital - public / communal areas (choice=Now) |
| 115 | 2 | 1 | 0 | 0 | **job_areas_11___2** | Numeric | Hospital - public / communal areas (choice=UK national lockdown) |
| 116 | 2 | 1 | 0 | 0 | **job_areas_11___99** | Numeric | Hospital - public / communal areas (choice=Prefer not to answer) |
| 117 | 2 | 1 | 0 | 0 | **job_areas_12___1** | Numeric | Laboratory (choice=Now) |
| 118 | 2 | 1 | 0 | 0 | **job_areas_12___2** | Numeric | Laboratory (choice=UK national lockdown) |
| 119 | 2 | 1 | 0 | 0 | **job_areas_12___99** | Numeric | Laboratory (choice=Prefer not to answer) |
| 120 | 2 | 1 | 0 | 0 | **job_areas_13___1** | Numeric | Maternity (choice=Now) |
| 121 | 2 | 1 | 0 | 0 | **job_areas_13___2** | Numeric | Maternity (choice=UK national lockdown) |
| 122 | 2 | 1 | 0 | 0 | **job_areas_13___99** | Numeric | Maternity (choice=Prefer not to answer) |
| 123 | 2 | 1 | 0 | 0 | **job_areas_14___1** | Numeric | Mobile across areas (choice=Now) |

| | | | | | | | |
|---|---|---|---|---|---|---|---|
| 124 | 2 | 1 | 0 | 0 | job_areas_14___2 | Numeric | Mobile across areas (choice=UK national lockdown) |
| 125 | 2 | 1 | 0 | 0 | job_areas_14___99 | Numeric | Mobile across areas (choice=Prefer not to answer) |
| 126 | 2 | 1 | 0 | 0 | job_areas_15___1 | Numeric | Nursing or care home (choice=Now) |
| 127 | 2 | 1 | 0 | 0 | job_areas_15___2 | Numeric | Nursing or care home (choice=UK national lockdown) |
| 128 | 2 | 1 | 0 | 0 | job_areas_15___99 | Numeric | Nursing or care home (choice=Prefer not to answer) |
| 129 | 2 | 1 | 0 | 0 | job_areas_16___1 | Numeric | Prison (choice=Now) |
| 130 | 2 | 1 | 0 | 0 | job_areas_16___2 | Numeric | Prison (choice=UK national lockdown) |
| 131 | 2 | 1 | 0 | 0 | job_areas_16___99 | Numeric | Prison (choice=Prefer not to answer) |
| 132 | 2 | 1 | 0 | 0 | job_areas_17___1 | Numeric | Psychiatric hospital or inpatient unit (choice=Now) |
| 133 | 2 | 1 | 0 | 0 | job_areas_17___2 | Numeric | Psychiatric hospital or inpatient unit (choice=UK national lockdown) |
| 134 | 2 | 1 | 0 | 0 | job_areas_17___99 | Numeric | Psychiatric hospital or inpatient unit (choice=Prefer not to answer) |
| 135 | 2 | 1 | 0 | 0 | job_areas_18___1 | Numeric | University (choice=Now) |
| 136 | 2 | 1 | 0 | 0 | job_areas_18___2 | Numeric | University (choice=UK national lockdown) |
| 137 | 2 | 1 | 0 | 0 | job_areas_18___99 | Numeric | University (choice=Prefer not to answer) |
| 138 | 2 | 1 | 0 | 0 | job_areas_19___1 | Numeric | Your home (choice=Now) |
| 139 | 2 | 1 | 0 | 0 | job_areas_19___2 | Numeric | Your home (choice=UK national lockdown) |
| 140 | 2 | 1 | 0 | 0 | job_areas_19___99 | Numeric | Your home (choice=Prefer not to answer) |
| 141 | 2 | 1 | 0 | 0 | job_areas_20___1 | Numeric | Other (Please specify) (choice=Now) |
| 142 | 2 | 1 | 0 | 0 | job_areas_20___2 | Numeric | Other (Please specify) (choice=UK national lockdown) |
| 143 | 2 | 1 | 0 | 0 | job_areas_20___99 | Numeric | Other (Please specify) (choice=Prefer not to answer) |
| 144 | 2 | 1 | 0 | 0 | job_areas_other | String | Please specify the workplace for which you selected Other: |
| 145 | 2 | 1 | 0 | 0 | work_hrsnow | Numeric | At present, how many hours do you workÂ in a typical week? |
| 146 | 2 | 1 | 0 | 0 | work_hrsld | Numeric | In the first month after the start of the UK national lockdown on 23rd March 2020, how many hours did you workÂ in a typical week? |
| 147 | 2 | 1 | 0 | 0 | job_nights1 | Numeric | Now |
| 148 | 2 | 1 | 0 | 0 | job_nights2 | Numeric | In the early months following the start of the UK national lockdown on 23rd March 2020 |
| 149 | 2 | 1 | 0 | 0 | jobncommnowwrempts | Numeric | Number of patients |
| 150 | 2 | 1 | 0 | 0 | jobncommnowwremcol | Numeric | Number of colleagues |
| 151 | 2 | 1 | 0 | 0 | jobncommnowwremoth | Numeric | Number of others (not patients or colleagues) |
| 152 | 2 | 1 | 0 | 0 | jobncommnowwftfpwc | Numeric | Number of patients with confirmed or suspected COVID-19 |
| 153 | 2 | 1 | 0 | 0 | jobncommnowwftfpwoc | Numeric | Number of other patients |
| 154 | 2 | 1 | 0 | 0 | jobncommnowwftfcol | Numeric | Number of colleagues |
| 155 | 2 | 1 | 0 | 0 | jobncommnowwftfoth | Numeric | Number of others (not patients or colleagues) |
| 156 | 2 | 1 | 0 | 0 | jobncommnowwwpcpwc | Numeric | Number of patients with confirmed or suspected COVID-19 |
| 157 | 2 | 1 | 0 | 0 | jobncommnowwwpcpwoc | Numeric | Number of other patients |
| 158 | 2 | 1 | 0 | 0 | jobncommnowwwpccol | Numeric | Number of colleagues |
| 159 | 2 | 1 | 0 | 0 | jobncommnowwwpcoth | Numeric | Number of others (not patients or colleagues) |
| 160 | 2 | 1 | 0 | 0 | pastworkncommwrempts | Numeric | Number of patients |
| 161 | 2 | 1 | 0 | 0 | pastworkncommwremcol | Numeric | Number of colleagues |
| 162 | 2 | 1 | 0 | 0 | pastworkncommwremoth | Numeric | Number of others (not patients or colleagues) |
| 163 | 2 | 1 | 0 | 0 | pastworkncommwftfpwc | Numeric | Number of patients with confirmed or suspected COVID-19 |
| 164 | 2 | 1 | 0 | 0 | pastworkncommwftfpwoc | Numeric | Number of other patients |
| 165 | 2 | 1 | 0 | 0 | pastworkncommwftfcol | Numeric | Number of colleagues |
| 166 | 2 | 1 | 0 | 0 | pastworkncommwftfoth | Numeric | Number of others (not patients or colleagues) |
| 167 | 2 | 1 | 0 | 0 | pastworkncommwwpcpwc | Numeric | Number of patients with confirmed or suspected COVID-19 |
| 168 | 2 | 1 | 0 | 0 | pastworkncommwwpcpwoc | Numeric | Number of other patients |
| 169 | 2 | 1 | 0 | 0 | pastworkncommwwpccol | Numeric | Number of colleagues |
| 170 | 2 | 1 | 0 | 0 | pastworkncommwwpcoth | Numeric | Number of others (not patients or colleagues) |
| 171 | 2 | 1 | 0 | 0 | jobtraveltime_now | Numeric | Typical working day over the past month |
| 172 | 2 | 1 | 0 | 0 | jobtraveltime_ld | Numeric | Typical working day during the first month after the UK national lockdown on 23rd March 2020 |
| 173 | 2 | 1 | 0 | 0 | jobtravelmode_now___1 | Numeric | Which of the following modes of transport do you use to commute on a typical working day over the past month? Please select all that apply.Â (choice=Car, alone or with member of household) |
| 174 | 2 | 1 | 0 | 0 | jobtravelmode_now___2 | Numeric | Which of the following modes of transport do you use to commute on a typical working day over the past month? Please select all that apply.Â (choice=Car share, with a small pool of people outside of household) |
| 175 | 2 | 1 | 0 | 0 | jobtravelmode_now___3 | Numeric | Which of the following modes of transport do you use to commute on a typical working day over the past month? Please select all that apply.Â (choice=Taxi or private hire vehicle) |
| 176 | 2 | 1 | 0 | 0 | jobtravelmode_now___4 | Numeric | Which of the following modes of transport do you use to commute on a typical working day over the past month? Please select all that apply.Â (choice=Public transport (e.g. bus, train, tram, underground)) |
| 177 | 2 | 1 | 0 | 0 | jobtravelmode_now___5 | Numeric | Which of the following modes of transport do you use to commute on a typical working day over the past month? Please select all that apply.Â (choice=Motorcycle, scooter or moped) |
| 178 | 2 | 1 | 0 | 0 | jobtravelmode_now___6 | Numeric | Which of the following modes of transport do you use to commute on a typical working day over the past month? Please select all that apply.Â (choice=Bicycle) |
| 179 | 2 | 1 | 0 | 0 | jobtravelmode_now___7 | Numeric | Which of the following modes of transport do you use to commute on a typical working day over the past month? Please select all that apply.Â (choice=On foot) |
| 180 | 2 | 1 | 0 | 0 | jobtravelmode_now___8 | Numeric | Which of the following modes of transport do you use to commute on a typical working day over the past month? Please select all that apply.Â (choice=Other) |
| 181 | 2 | 1 | 0 | 0 | jobtravelmode_now_other | String | Please enter the mode of transport for which you selected Other: |
| 182 | 2 | 1 | 0 | 0 | jobtravelmode_ld___1 | Numeric | Which of the following modes of transport did you use to commute on a typical working day, during the first monthÂ afterÂ the start of the UK national lockdown on 23 March 2020? Please select all that apply.Â (choice=Car, alone or with member of hou |
| 183 | 2 | 1 | 0 | 0 | jobtravelmode_ld___2 | Numeric | Which of the following modes of transport did you use to commute on a typical working day, during the first monthÂ afterÂ the start of the UK national lockdown on 23 March 2020? Please select all that apply.Â (choice=Car share, with a small pool of |
| 184 | 2 | 1 | 0 | 0 | jobtravelmode_ld___3 | Numeric | Which of the following modes of transport did you use to commute on a typical working day, during the first monthÂ afterÂ the start of the UK national lockdown on 23 March 2020? Please select all that apply.Â (choice=Taxi or private hire vehicle) |
| 185 | 2 | 1 | 0 | 0 | jobtravelmode_ld___4 | Numeric | Which of the following modes of transport did you use to commute on a typical working day, during the first monthÂ afterÂ the start of the UK national lockdown on 23 March 2020? Please select all that apply.Â (choice=Public transport (e.g. bus, trai |
| 186 | 2 | 1 | 0 | 0 | jobtravelmode_ld___5 | Numeric | Which of the following modes of transport did you use to commute on a typical working day, during the first monthÂ afterÂ the start of the UK national lockdown on 23 March 2020? Please select all that apply.Â (choice=Motorcycle, scooter or moped) |
| 187 | 2 | 1 | 0 | 0 | jobtravelmode_ld___6 | Numeric | Which of the following modes of transport did you use to commute on a typical working day, during the first monthÂ afterÂ the start of the UK national lockdown on 23 March 2020? Please select all that apply.Â (choice=Bicycle) |

| | | | | | | | |
|---|---|---|---|---|---|---|---|
| 188 | 2 | 1 | 0 | 0 | jobtravelmode_ld___7 | Numeric | Which of the following modes of transport did you use to commute on a typical working day, during the first monthÂ afterÂ the start of the UK national lockdown on 23 March 2020? Please select all that apply.Â (choice=On foot) |
| 189 | 2 | 1 | 0 | 0 | jobtravelmode_ld___8 | Numeric | Which of the following modes of transport did you use to commute on a typical working day, during the first monthÂ afterÂ the start of the UK national lockdown on 23 March 2020? Please select all that apply.Â (choice=Other) |
| 190 | 2 | 1 | 0 | 0 | jobtravelmode_ld_other | String | Please enter the mode of transport for which you selected Other: |
| 191 | 2 | 1 | 0 | 0 | workppeacc_new_now | Numeric | At present, do you have access to appropriate personal protective equipment (PPE) at work? |
| 192 | 2 | 1 | 0 | 0 | workppeacc_new_ld | Numeric | In the first month after the start of the UK national lockdown on 23rd March 2020, did you have access to appropriate personal protective equipment (PPE) at work? |
| 193 | 2 | 1 | 0 | 0 | workppetrain_new_1___1 | Numeric | Have you received training in the use of personal protective equipment (PPE) for your current work? (choice=Not applicable) |
| 194 | 2 | 1 | 0 | 0 | workppetrain_new_1___2 | Numeric | Have you received training in the use of personal protective equipment (PPE) for your current work? (choice=Formal training in person) |
| 195 | 2 | 1 | 0 | 0 | workppetrain_new_1___3 | Numeric | Have you received training in the use of personal protective equipment (PPE) for your current work? (choice=Formal training online) |
| 196 | 2 | 1 | 0 | 0 | workppetrain_new_1___4 | Numeric | Have you received training in the use of personal protective equipment (PPE) for your current work? (choice=Informal training) |
| 197 | 2 | 1 | 0 | 0 | workppetrain_new_1___5 | Numeric | Have you received training in the use of personal protective equipment (PPE) for your current work? (choice=No training) |
| 198 | 2 | 1 | 0 | 0 | workppetrain_new_1___99 | Numeric | Have you received training in the use of personal protective equipment (PPE) for your current work? (choice=Prefer not to answer) |
| 199 | 2 | 1 | 0 | 0 | workppetrain_new_2___1 | Numeric | In the first month after the start of the UK national lockdown on 23rd March 2020, did you receive training in the use of personal protective equipment (PPE) for your work? (choice=Not applicable) |
| 200 | 2 | 1 | 0 | 0 | workppetrain_new_2___2 | Numeric | In the first month after the start of the UK national lockdown on 23rd March 2020, did you receive training in the use of personal protective equipment (PPE) for your work? (choice=Formal training in person) |
| 201 | 2 | 1 | 0 | 0 | workppetrain_new_2___3 | Numeric | In the first month after the start of the UK national lockdown on 23rd March 2020, did you receive training in the use of personal protective equipment (PPE) for your work? (choice=Formal training online) |
| 202 | 2 | 1 | 0 | 0 | workppetrain_new_2___4 | Numeric | In the first month after the start of the UK national lockdown on 23rd March 2020, did you receive training in the use of personal protective equipment (PPE) for your work? (choice=Informal training) |
| 203 | 2 | 1 | 0 | 0 | workppetrain_new_2___5 | Numeric | In the first month after the start of the UK national lockdown on 23rd March 2020, did you receive training in the use of personal protective equipment (PPE) for your work? (choice=No training) |
| 204 | 2 | 1 | 0 | 0 | workppetrain_new_2___99 | Numeric | In the first month after the start of the UK national lockdown on 23rd March 2020, did you receive training in the use of personal protective equipment (PPE) for your work? (choice=Prefer not to answer) |
| 205 | 2 | 1 | 0 | 0 | workagp_new_1 | Numeric | At present, how often are you in a room where aerosol-generating procedures are performed? |
| 206 | 2 | 1 | 0 | 0 | workagp_new_2 | Numeric | In the first month after the start of the UK national lockdown on 23rd March 2020, how often were you in a room where aerosol-generating procedures are performed? |
| 207 | 2 | 1 | 0 | 0 | workrskassoff | Numeric | Have you been offered an NHS COVID-19 risk assessment at work? |
| 208 | 2 | 1 | 0 | 0 | workrskassch | Numeric | Did your work change as a result of the NHS COVID-19 risk assessment result? (Select the best answer) Â |
| 209 | 2 | 1 | 0 | 0 | workrskassch_other | String | Please specify how your work changed as a result of the NHS COVID-19 risk assessment result: |
| 210 | 2 | 1 | 0 | 0 | workconcern_1 | Numeric | I would feel secure raising concerns about unsafe clinical practice |
| 211 | 2 | 1 | 0 | 0 | workconcern_2 | Numeric | I am confident that my organisation would address my concern |
| 212 | 2 | 1 | 0 | 0 | pastjobredep | Numeric | During the UK national lockdown that began on 23rd March 2020, were you redeployed to a different role because of the pandemic? |
| 213 | 2 | 1 | 0 | 0 | pastjobredep2 | Numeric | Compared to your role before the start of UK national lockdown on 23 March 2020, how much direct patient contact is there or was there in your redeployed role? |
| 214 | 2 | 1 | 0 | 0 | pastjobredeptrain___1 | Numeric | Did you have any of the following in your redeployment?Select all that apply (choice=Formal training face to face) |
| 215 | 2 | 1 | 0 | 0 | pastjobredeptrain___2 | Numeric | Did you have any of the following in your redeployment?Select all that apply (choice=Formal training online) |
| 216 | 2 | 1 | 0 | 0 | pastjobredeptrain___3 | Numeric | Did you have any of the following in your redeployment?Select all that apply (choice=Informal training) |
| 217 | 2 | 1 | 0 | 0 | pastjobredeptrain___4 | Numeric | Did you have any of the following in your redeployment?Select all that apply (choice=No training) |
| 218 | 2 | 1 | 0 | 0 | pastjobredeptrain___99 | Numeric | Did you have any of the following in your redeployment?Select all that apply (choice=Prefer not to answer) |
| 219 | 2 | 1 | 0 | 0 | pastjobredepsup___1 | Numeric | Did you have any of the following types of supervision in your redeployment? Select all that apply (choice=Formal supervision) |
| 220 | 2 | 1 | 0 | 0 | pastjobredepsup___2 | Numeric | Did you have any of the following types of supervision in your redeployment? Select all that apply (choice=Informal supervision) |
| 221 | 2 | 1 | 0 | 0 | pastjobredepsup___3 | Numeric | Did you have any of the following types of supervision in your redeployment? Select all that apply (choice=No supervision) |
| 222 | 2 | 1 | 0 | 0 | pastjobredepsup___99 | Numeric | Did you have any of the following types of supervision in your redeployment? Select all that apply (choice=Prefer not to answer) |
| 223 | 3 | 0 | 0 | 1 | Section3_Raw_EthnicityEtc______________Raw | Numeric | |
| 224 | 3 | 1 | 0 | 0 | ethown | Numeric | What is your ethnic group?Select the one that best describes your ethnic group or background. The categories are the ethnic groups used in the UK National Census. |
| 225 | 3 | 1 | 0 | 0 | ethown_other | String | Please specify your ethnic group: |
| 226 | 3 | 1 | 0 | 0 | ethbornuk | Numeric | Were you born in the UK? |
| 227 | 3 | 1 | 0 | 0 | ethnborn | Numeric | In which country were you born? |
| 228 | 3 | 1 | 0 | 0 | ethnborn_other | String | Please specify the country in which you were born: |
| 229 | 3 | 1 | 0 | 0 | ethyearuk | Numeric | In which year did you move to the UK?If you are unsure, please give your best estimate. |
| 230 | 3 | 1 | 0 | 0 | eth_nat_1 | Numeric | Nationality 1: |
| 231 | 3 | 1 | 0 | 0 | eth_nat_1_other | String | Please specify nationality 1: |
| 232 | 3 | 1 | 0 | 0 | eth_nat_2 | Numeric | Nationality 2: |
| 233 | 3 | 1 | 0 | 0 | eth_nat_2_other | String | Please specify nationality 2: |
| 234 | 3 | 1 | 0 | 0 | eth_nat_3 | Numeric | Nationality 3: |
| 235 | 3 | 1 | 0 | 0 | eth_nat_3_other | Numeric | Please specify nationality 3: |
| 236 | 3 | 1 | 0 | 0 | ethp | Numeric | What is or was your partners ethnic group?The following categories are the ethnic groups used in the UK National Census. |
| 237 | 3 | 1 | 0 | 0 | ethp_other_2 | String | Please specify your partners ethnic group: |
| 238 | 3 | 1 | 0 | 0 | ethm | Numeric | What is or was your mothers ethnic group? The following categories are the ethnic groups used in the UK National Census. |
| 239 | 3 | 1 | 0 | 0 | ethm_other | String | Please specify your mothers ethnic group: |
| 240 | 3 | 1 | 0 | 0 | ethprntukm | Numeric | Was your mother born in the UK? |
| 241 | 3 | 1 | 0 | 0 | ethprntukmc | Numeric | In which country was your mother born? |
| 242 | 3 | 1 | 0 | 0 | ethnprntmc_other | String | Please specify the country in which your mother was born: |
| 243 | 3 | 1 | 0 | 0 | ethf | Numeric | What is or was your fathers ethnic group?The following categories are the ethnic groups used in the UK National Census. |
| 244 | 3 | 1 | 0 | 0 | ethf_other | String | Please specify your fathers ethnic group: |
| 245 | 3 | 1 | 0 | 0 | ethprntukf | Numeric | Was your father born in the UK? |
| 246 | 3 | 1 | 0 | 0 | ethprntukfc | Numeric | In which country was your father born? |
| 247 | 3 | 1 | 0 | 0 | ethnprntfc_other | String | Please specify the country in which your father was born: |
| 248 | 3 | 1 | 0 | 0 | ethgprntukmatgm | Numeric | Your mothers mother |
| 249 | 3 | 1 | 0 | 0 | ethgprntukmatgf | Numeric | Your mothers father |
| 250 | 3 | 1 | 0 | 0 | ethgprntukpatgm | Numeric | Your fathers mother |
| 251 | 3 | 1 | 0 | 0 | ethgprntukpatgf | Numeric | Your fathers father |

| | | | | | | | |
|---|---|---|---|---|---|---|---|
| 252 | 3 | 1 | 0 | 0 | **langedeng** | Numeric | How old were you when you first learned English? |
| 253 | 3 | 1 | 0 | 0 | **langedhome_1** | Numeric | Language 1 |
| 254 | 3 | 1 | 0 | 0 | **langedhome_1_other** | String | Please specify language 1: |
| 255 | 3 | 1 | 0 | 0 | **langedhome_2** | Numeric | Language 2 |
| 256 | 3 | 1 | 0 | 0 | **langedhome_2_other** | String | Please specify language 2: |
| 257 | 3 | 1 | 0 | 0 | **langedch___11** | Numeric | What language was spoken in your home when you were a child? If more than one language was spoken, please give them all. (choice=English) |
| 258 | 3 | 1 | 0 | 0 | **langedch___1** | Numeric | What language was spoken in your home when you were a child? If more than one language was spoken, please give them all. (choice=Afrikaans) |
| 259 | 3 | 1 | 0 | 0 | **langedch___2** | Numeric | What language was spoken in your home when you were a child? If more than one language was spoken, please give them all. (choice=Akan) |
| 260 | 3 | 1 | 0 | 0 | **langedch___3** | Numeric | What language was spoken in your home when you were a child? If more than one language was spoken, please give them all. (choice=Arabic) |
| 261 | 3 | 1 | 0 | 0 | **langedch___4** | Numeric | What language was spoken in your home when you were a child? If more than one language was spoken, please give them all. (choice=Bengali) |
| 262 | 3 | 1 | 0 | 0 | **langedch___5** | Numeric | What language was spoken in your home when you were a child? If more than one language was spoken, please give them all. (choice=Bulgarian) |
| 263 | 3 | 1 | 0 | 0 | **langedch___6** | Numeric | What language was spoken in your home when you were a child? If more than one language was spoken, please give them all. (choice=Burmese) |
| 264 | 3 | 1 | 0 | 0 | **langedch___7** | Numeric | What language was spoken in your home when you were a child? If more than one language was spoken, please give them all. (choice=Cebuano) |
| 265 | 3 | 1 | 0 | 0 | **langedch___8** | Numeric | What language was spoken in your home when you were a child? If more than one language was spoken, please give them all. (choice=Chinese) |
| 266 | 3 | 1 | 0 | 0 | **langedch___9** | Numeric | What language was spoken in your home when you were a child? If more than one language was spoken, please give them all. (choice=Czech) |
| 267 | 3 | 1 | 0 | 0 | **langedch___10** | Numeric | What language was spoken in your home when you were a child? If more than one language was spoken, please give them all. (choice=Dutch) |
| 268 | 3 | 1 | 0 | 0 | **langedch___12** | Numeric | What language was spoken in your home when you were a child? If more than one language was spoken, please give them all. (choice=French) |
| 269 | 3 | 1 | 0 | 0 | **langedch___13** | Numeric | What language was spoken in your home when you were a child? If more than one language was spoken, please give them all. (choice=German) |
| 270 | 3 | 1 | 0 | 0 | **langedch___14** | Numeric | What language was spoken in your home when you were a child? If more than one language was spoken, please give them all. (choice=Greek) |
| 271 | 3 | 1 | 0 | 0 | **langedch___15** | Numeric | What language was spoken in your home when you were a child? If more than one language was spoken, please give them all. (choice=Gujarati) |
| 272 | 3 | 1 | 0 | 0 | **langedch___16** | Numeric | What language was spoken in your home when you were a child? If more than one language was spoken, please give them all. (choice=Hausa) |
| 273 | 3 | 1 | 0 | 0 | **langedch___17** | Numeric | What language was spoken in your home when you were a child? If more than one language was spoken, please give them all. (choice=Hindi) |
| 274 | 3 | 1 | 0 | 0 | **langedch___18** | Numeric | What language was spoken in your home when you were a child? If more than one language was spoken, please give them all. (choice=Hungarian) |
| 275 | 3 | 1 | 0 | 0 | **langedch___19** | Numeric | What language was spoken in your home when you were a child? If more than one language was spoken, please give them all. (choice=Igbo) |
| 276 | 3 | 1 | 0 | 0 | **langedch___20** | Numeric | What language was spoken in your home when you were a child? If more than one language was spoken, please give them all. (choice=Irish) |
| 277 | 3 | 1 | 0 | 0 | **langedch___21** | Numeric | What language was spoken in your home when you were a child? If more than one language was spoken, please give them all. (choice=Italian) |
| 278 | 3 | 1 | 0 | 0 | **langedch___22** | Numeric | What language was spoken in your home when you were a child? If more than one language was spoken, please give them all. (choice=Kurdish) |
| 279 | 3 | 1 | 0 | 0 | **langedch___50** | Numeric | What language was spoken in your home when you were a child? If more than one language was spoken, please give them all. (choice=Latvian) |
| 280 | 3 | 1 | 0 | 0 | **langedch___23** | Numeric | What language was spoken in your home when you were a child? If more than one language was spoken, please give them all. (choice=Lithuanian) |
| 281 | 3 | 1 | 0 | 0 | **langedch___24** | Numeric | What language was spoken in your home when you were a child? If more than one language was spoken, please give them all. (choice=Malay) |
| 282 | 3 | 1 | 0 | 0 | **langedch___25** | Numeric | What language was spoken in your home when you were a child? If more than one language was spoken, please give them all. (choice=Malayalam) |
| 283 | 3 | 1 | 0 | 0 | **langedch___26** | Numeric | What language was spoken in your home when you were a child? If more than one language was spoken, please give them all. (choice=Maltese) |
| 284 | 3 | 1 | 0 | 0 | **langedch___51** | Numeric | What language was spoken in your home when you were a child? If more than one language was spoken, please give them all. (choice=Maori) |
| 285 | 3 | 1 | 0 | 0 | **langedch___27** | Numeric | What language was spoken in your home when you were a child? If more than one language was spoken, please give them all. (choice=Ndebele) |
| 286 | 3 | 1 | 0 | 0 | **langedch___28** | Numeric | What language was spoken in your home when you were a child? If more than one language was spoken, please give them all. (choice=Nepali) |
| 287 | 3 | 1 | 0 | 0 | **langedch___29** | Numeric | What language was spoken in your home when you were a child? If more than one language was spoken, please give them all. (choice=Persian/Farsi) |
| 288 | 3 | 1 | 0 | 0 | **langedch___30** | Numeric | What language was spoken in your home when you were a child? If more than one language was spoken, please give them all. (choice=Polish) |
| 289 | 3 | 1 | 0 | 0 | **langedch___31** | Numeric | What language was spoken in your home when you were a child? If more than one language was spoken, please give them all. (choice=Portuguese) |
| 290 | 3 | 1 | 0 | 0 | **langedch___32** | Numeric | What language was spoken in your home when you were a child? If more than one language was spoken, please give them all. (choice=Punjabi) |
| 291 | 3 | 1 | 0 | 0 | **langedch___33** | Numeric | What language was spoken in your home when you were a child? If more than one language was spoken, please give them all. (choice=Romanian) |
| 292 | 3 | 1 | 0 | 0 | **langedch___34** | Numeric | What language was spoken in your home when you were a child? If more than one language was spoken, please give them all. (choice=Russian) |
| 293 | 3 | 1 | 0 | 0 | **langedch___35** | Numeric | What language was spoken in your home when you were a child? If more than one language was spoken, please give them all. (choice=Shona) |
| 294 | 3 | 1 | 0 | 0 | **langedch___36** | Numeric | What language was spoken in your home when you were a child? If more than one language was spoken, please give them all. (choice=Sinhala) |
| 295 | 3 | 1 | 0 | 0 | **langedch___52** | Numeric | What language was spoken in your home when you were a child? If more than one language was spoken, please give them all. (choice=Slovakian) |
| 296 | 3 | 1 | 0 | 0 | **langedch___37** | Numeric | What language was spoken in your home when you were a child? If more than one language was spoken, please give them all. (choice=Somali) |
| 297 | 3 | 1 | 0 | 0 | **langedch___38** | Numeric | What language was spoken in your home when you were a child? If more than one language was spoken, please give them all. (choice=Spanish) |
| 298 | 3 | 1 | 0 | 0 | **langedch___49** | Numeric | What language was spoken in your home when you were a child? If more than one language was spoken, please give them all. (choice=Swahili) |
| 299 | 3 | 1 | 0 | 0 | **langedch___53** | Numeric | What language was spoken in your home when you were a child? If more than one language was spoken, please give them all. (choice=Swedish) |
| 300 | 3 | 1 | 0 | 0 | **langedch___39** | Numeric | What language was spoken in your home when you were a child? If more than one language was spoken, please give them all. (choice=Tagalog) |
| 301 | 3 | 1 | 0 | 0 | **langedch___40** | Numeric | What language was spoken in your home when you were a child? If more than one language was spoken, please give them all. (choice=Tamil) |
| 302 | 3 | 1 | 0 | 0 | **langedch___41** | Numeric | What language was spoken in your home when you were a child? If more than one language was spoken, please give them all. (choice=Turkish) |
| 303 | 3 | 1 | 0 | 0 | **langedch___54** | Numeric | What language was spoken in your home when you were a child? If more than one language was spoken, please give them all. (choice=Twi) |
| 304 | 3 | 1 | 0 | 0 | **langedch___42** | Numeric | What language was spoken in your home when you were a child? If more than one language was spoken, please give them all. (choice=Ukrainian) |
| 305 | 3 | 1 | 0 | 0 | **langedch___43** | Numeric | What language was spoken in your home when you were a child? If more than one language was spoken, please give them all. (choice=Urdu) |
| 306 | 3 | 1 | 0 | 0 | **langedch___44** | Numeric | What language was spoken in your home when you were a child? If more than one language was spoken, please give them all. (choice=Welsh) |
| 307 | 3 | 1 | 0 | 0 | **langedch___45** | Numeric | What language was spoken in your home when you were a child? If more than one language was spoken, please give them all. (choice=Xhosa) |
| 308 | 3 | 1 | 0 | 0 | **langedch___46** | Numeric | What language was spoken in your home when you were a child? If more than one language was spoken, please give them all. (choice=Yoruba) |
| 309 | 3 | 1 | 0 | 0 | **langedch___47** | Numeric | What language was spoken in your home when you were a child? If more than one language was spoken, please give them all. (choice=Zulu) |
| 310 | 3 | 1 | 0 | 0 | **langedch___48** | Numeric | What language was spoken in your home when you were a child? If more than one language was spoken, please give them all. (choice=Other (Please specify)) |
| 311 | 3 | 1 | 0 | 0 | **langedch___99** | Numeric | What language was spoken in your home when you were a child? If more than one language was spoken, please give them all. (choice=Prefer not to answer) |
| 312 | 3 | 1 | 0 | 0 | **langedch_other** | String | Please specify the language spoken in your home when you were a child for which you selected Other: |
| 313 | 3 | 1 | 0 | 0 | **relig** | Numeric | What is your religion? This is the question that was asked in the 2011 UK Census. |
| 314 | 3 | 1 | 0 | 0 | **rel_other** | String | Please specify your religion: |
| 315 | 3 | 1 | 0 | 0 | **relimp1** | Numeric | How important is religion to you in your everyday life? |

| | | | | | | | |
|---|---|---|---|---|---|---|---|
| 316 | 3 | 1 | 0 | 0 | relimp2 | Numeric | How important was religion in your upbringing? |
| 317 | 3 | 1 | 0 | 0 | relobs1 | Numeric | How often would you usually attend a holy place or a place of worship outside your home? |
| 318 | 3 | 1 | 0 | 0 | ethimp | Numeric | How important is yourÂ ethnic and cultural background to your identity?Use the scale of 0 to 10, where 0 means not at all important, and 10 means extremely important. |
| 319 | 3 | 1 | 0 | 0 | ethimpm | Numeric | How important is/wasÂ your mothers ethnic and cultural background to your identity?Use the scale of 0 to 10, where 0 means not at all important, and 10 means extremely important. |
| 320 | 3 | 1 | 0 | 0 | ethimpf | Numeric | How important is/wasÂ your fathers ethnic and cultural background to your identity?Use the scale of 0 to 10, where 0 means not at all important, and 10 means extremely important. |
| 321 | 3 | 1 | 0 | 0 | ethimpp | Numeric | How important is/wasÂ your partners ethnic and cultural background to your identity?Use the scale of 0 to 10, where 0 means not at all important, and 10 means extremely important. |
| 322 | 3 | 1 | 0 | 0 | langedpmq | Numeric | In which country did you gain your primary professional qualification?(The qualification used for registration with your professional regulator, such as the GDC, GMC, GOC, GPhC, PSNI, HCPC, NMC) |
| 323 | 3 | 1 | 0 | 0 | langedpmq_other | String | Please specify the country in which you gained your primary professional qualification: |
| 324 | 3 | 1 | 0 | 0 | langedyear | Numeric | In which year did you obtain your primary professional qualification? |
| 325 | 3 | 1 | 0 | 0 | langedqual | Numeric | What is the highest level of education you have completed? |
| 326 | 3 | 1 | 0 | 0 | langedqualm | Numeric | What is the highest level of education your mother has completed? |
| 327 | 3 | 1 | 0 | 0 | langedqualf | Numeric | What is the highest level of education your father has completed? |
| 328 | 3 | 1 | 0 | 0 | workethn | Numeric | In your current main job/role, what proportion of colleagues who are senior to you are of the same ethnic group as yourself? |
| 329 | 3 | 1 | 0 | 0 | workwhite | Numeric | In your current main job/role, what proportion of your colleagues who are senior to you are White? |
| 330 | 3 | 1 | 0 | 0 | workfair | Numeric | Thinking about where you work in your current main job/role, does your organisation act fairly with regard to career progression / promotion, regardless of ethnic background, gender, religion, sexual orientation, disability or age? |
| 331 | 4 | 0 | 0 | 1 | Section4_Raw_HomeAndFamilyLife_______________Raw | Numeric | |
| 332 | 4 | 1 | 0 | 0 | hf_sup_bub | Numeric | Do you have a âsupport bubbleâ (in England or Northern Ireland) or âextended householdâ (in Wales or Scotland) which includes people who usually live at a different address? |
| 333 | 4 | 1 | 0 | 0 | hf_sup_bub_2 | Numeric | How many people are in this support bubble? Only count those who usually live at a different address. |
| 334 | 4 | 1 | 0 | 0 | hf_chld_bub | Numeric | Do you have a âchildcare bubbleâ which includes people who usually live at a different address? Do not include anyone already counted in the support bubble in the previous question. |
| 335 | 4 | 1 | 0 | 0 | hf_chld_bub_2 | Numeric | How many people are in this childcare bubble? Only count those who usually live at a different address. |
| 336 | 4 | 1 | 0 | 0 | hf_nhhold | Numeric | Apart from you, how many other people are in your household? |
| 337 | 4 | 1 | 0 | 0 | hfp1rel | Numeric | Person 1 - What best describes this persons relationship to you? Please select from the list provided. |
| 338 | 4 | 1 | 0 | 0 | hfp1age | Numeric | Person 1 - How old is this person? |
| 339 | 4 | 1 | 0 | 0 | hfp2rel | Numeric | Person 2 - What best describes this persons relationship to you? |
| 340 | 4 | 1 | 0 | 0 | hfp2age | Numeric | Person 2 - How old is this person? |
| 341 | 4 | 1 | 0 | 0 | hfp3rel | Numeric | Person 3 - What best describes this persons relationship to you? Please select from the list provided. |
| 342 | 4 | 1 | 0 | 0 | hfp3age | Numeric | Person 3 - How old is this person? |
| 343 | 4 | 1 | 0 | 0 | hfp4rel | Numeric | Person 4 - What best describes this persons relationship to you? Please select from the list provided. |
| 344 | 4 | 1 | 0 | 0 | hfp4age | Numeric | Person 4 - How old is this person? |
| 345 | 4 | 1 | 0 | 0 | hfp5rel | Numeric | Person 5 - What best describes this persons relationship to you? Please select from the list provided. |
| 346 | 4 | 1 | 0 | 0 | hfp5age | Numeric | Person 5 - How old is this person? |
| 347 | 4 | 1 | 0 | 0 | hfp6rel | Numeric | Person 6 - What best describes this persons relationship to you? Please select from the list provided. |
| 348 | 4 | 1 | 0 | 0 | hfp6age | Numeric | Person 6 - How old is this person? |
| 349 | 4 | 1 | 0 | 0 | hfp7rel | Numeric | Person 7 - What best describes this persons relationship to you? Please select from the list provided. |
| 350 | 4 | 1 | 0 | 0 | hfp7age | Numeric | Person 7 - How old is this person? |
| 351 | 4 | 1 | 0 | 0 | hfp8rel | Numeric | Person 8 - What best describes this persons relationship to you? Please select from the list provided. |
| 352 | 4 | 1 | 0 | 0 | hfp8age | Numeric | Person 8 - How old is this person? |
| 353 | 4 | 1 | 0 | 0 | hfp9rel | Numeric | Person 9 - What best describes this persons relationship to you? Please select from the list provided. |
| 354 | 4 | 1 | 0 | 0 | hfp9age | Numeric | Person 9 - How old is this person? |
| 355 | 4 | 1 | 0 | 0 | hfp10rel | Numeric | Person 10 - What best describes this persons relationship to you? Please select from the list provided. |
| 356 | 4 | 1 | 0 | 0 | hfp10age | Numeric | Person 10 - How old is this person? |
| 357 | 4 | 1 | 0 | 0 | hfp11rel | Numeric | Person 11 - What best describes this persons relationship to you? Please select from the list provided. |
| 358 | 4 | 1 | 0 | 0 | hfp11age | Numeric | Person 11 - How old is this person? |
| 359 | 4 | 1 | 0 | 0 | hfp12rel | Numeric | Person 12 - What best describes this persons relationship to you? Please select from the list provided. |
| 360 | 4 | 1 | 0 | 0 | hfp12age | Numeric | Person 12 - How old is this person? |
| 361 | 4 | 1 | 0 | 0 | hfp12more | String | If you live with more than 12 people, please state the relationship to you and ages for the others in this box separated by a comma, e.g.: Aunt 56, Colleague 25 |
| 362 | 4 | 1 | 0 | 0 | hfriskjob | Numeric | Apart from yourself, how many people in your household travel to work using public transport? |
| 363 | 4 | 1 | 0 | 0 | hfriskjob_2 | Numeric | Apart from yourself, how many people in your household work in jobs that often bring them into close physical contact (within 2 metres) with others? Some examples include: bus driver, carer, cleaner, doctor, supermarket checkout worker, teacher. |
| 364 | 4 | 1 | 0 | 0 | hfcuradd | Numeric | In which year did you move to your current address? |
| 365 | 4 | 1 | 0 | 0 | hfcuradd_months | Numeric | In which month did you move to your current address? |
| 366 | 4 | 1 | 0 | 0 | hfacctype | Numeric | What type of accommodation are you currently living in? |
| 367 | 4 | 1 | 0 | 0 | hfacctype_other | String | Please specify what type of accommodation you live in: |
| 368 | 4 | 1 | 0 | 0 | hfaccemp | Numeric | Is your current accommodation provided by or linked to your employer, e.g. hospital staff accommodation? |
| 369 | 4 | 1 | 0 | 0 | hfaccrooms | Numeric | How many rooms are in your accommodation (not including the kitchen and bathroom(s))? |
| 370 | 4 | 1 | 0 | 0 | hfaccshbath___4 | Numeric | Do you share any of the following rooms with people you do not consider to be a part of your household? You may select more than one answer. If you do not share any of the rooms listed, please select None. (choice=None) |
| 371 | 4 | 1 | 0 | 0 | hfaccshbath___1 | Numeric | Do you share any of the following rooms with people you do not consider to be a part of your household? You may select more than one answer. If you do not share any of the rooms listed, please select None. (choice=Kitchen) |
| 372 | 4 | 1 | 0 | 0 | hfaccshbath___2 | Numeric | Do you share any of the following rooms with people you do not consider to be a part of your household? You may select more than one answer. If you do not share any of the rooms listed, please select None. (choice=Bathroom) |
| 373 | 4 | 1 | 0 | 0 | hfaccshbath___3 | Numeric | Do you share any of the following rooms with people you do not consider to be a part of your household? You may select more than one answer. If you do not share any of the rooms listed, please select None. (choice=Living room, sitting room or dining area) |
| 374 | 4 | 1 | 0 | 0 | hfaccshbath___99 | Numeric | Do you share any of the following rooms with people you do not consider to be a part of your household? You may select more than one answer. If you do not share any of the rooms listed, please select None. (choice=Prefer not to answer) |
| 375 | 4 | 1 | 0 | 0 | hf_share | Numeric | Does your accommodation include shared communal areas such as hallways, stairwells or lifts? |
| 376 | 4 | 1 | 0 | 0 | hfgarden | Numeric | Does your accommodation have a safe outdoor space (e.g., a garden or yard) where you can exercise or relax? |
| 377 | 4 | 1 | 0 | 0 | hfgardenpriv | Numeric | Is your garden/yard shared with other households or private? |
| 378 | 5 | 0 | 0 | 1 | Section5_Raw_FriendsAndSocialNetwork_______________Raw | Numeric | |
| 379 | 5 | 1 | 0 | 0 | socncommnowrem | Numeric | Remotely (e.g. over the phone, social media or via video media) |

| | | | | | | | |
|---|---|---|---|---|---|---|---|
| 380 | 5 | 1 | 0 | 0 | socncommnowftfx | Numeric | Face-to-face with social distancing |
| 381 | 5 | 1 | 0 | 0 | socncommnowpcx | Numeric | With physical contact (e.g. handshake/hug/kiss, etc) |
| 382 | 5 | 1 | 0 | 0 | soceth2 | Numeric | What proportion of your friends are of the same ethnic group as yourself? |
| 383 | 6 | 0 | 0 | 1 | Section6_Raw_HarassmentAndDiscn_____________Raw | Numeric | |
| 384 | 6 | 1 | 0 | 0 | harasedds_1 | Numeric | You are treated with less courtesy than other people are. |
| 385 | 6 | 1 | 0 | 0 | harasedds_2 | Numeric | You are treated with less respect than other people are. |
| 386 | 6 | 1 | 0 | 0 | harasedds_3 | Numeric | You receive poorer service than other people at restaurants or shops. |
| 387 | 6 | 1 | 0 | 0 | harasedds_4 | Numeric | People act as if they think you are not smart. |
| 388 | 6 | 1 | 0 | 0 | harasedds_5 | Numeric | People act as if they are afraid of you. |
| 389 | 6 | 1 | 0 | 0 | harasedds_6 | Numeric | People act as if they think you are dishonest. |
| 390 | 6 | 1 | 0 | 0 | harasedds_7 | Numeric | People act as if theyre better than you are. |
| 391 | 6 | 1 | 0 | 0 | harasedds_8 | Numeric | You are called names or insulted. |
| 392 | 6 | 1 | 0 | 0 | harasedds_9 | Numeric | You are threatened or harassed. |
| 393 | 6 | 1 | 0 | 0 | harascaus___1 | Numeric | What do you think are the reasons for these experiences? Please select all that apply.Â (choice=Your national origins) |
| 394 | 6 | 1 | 0 | 0 | harascaus___2 | Numeric | What do you think are the reasons for these experiences? Please select all that apply.Â (choice=Your gender) |
| 395 | 6 | 1 | 0 | 0 | harascaus___3 | Numeric | What do you think are the reasons for these experiences? Please select all that apply.Â (choice=Your ethnicity) |
| 396 | 6 | 1 | 0 | 0 | harascaus___4 | Numeric | What do you think are the reasons for these experiences? Please select all that apply.Â (choice=Your age) |
| 397 | 6 | 1 | 0 | 0 | harascaus___5 | Numeric | What do you think are the reasons for these experiences? Please select all that apply.Â (choice=Your religion) |
| 398 | 6 | 1 | 0 | 0 | harascaus___6 | Numeric | What do you think are the reasons for these experiences? Please select all that apply.Â (choice=Your height) |
| 399 | 6 | 1 | 0 | 0 | harascaus___7 | Numeric | What do you think are the reasons for these experiences? Please select all that apply.Â (choice=Your weight) |
| 400 | 6 | 1 | 0 | 0 | harascaus___8 | Numeric | What do you think are the reasons for these experiences? Please select all that apply.Â (choice=Your health or disability) |
| 401 | 6 | 1 | 0 | 0 | harascaus___9 | Numeric | What do you think are the reasons for these experiences? Please select all that apply.Â (choice=Your dress) |
| 402 | 6 | 1 | 0 | 0 | harascaus___10 | Numeric | What do you think are the reasons for these experiences? Please select all that apply.Â (choice=Some other aspect of your physical appearance) |
| 403 | 6 | 1 | 0 | 0 | harascaus___11 | Numeric | What do you think are the reasons for these experiences? Please select all that apply.Â (choice=Your sexual orientation) |
| 404 | 6 | 1 | 0 | 0 | harascaus___12 | Numeric | What do you think are the reasons for these experiences? Please select all that apply.Â (choice=Your education or income level) |
| 405 | 6 | 1 | 0 | 0 | harascaus___13 | Numeric | What do you think are the reasons for these experiences? Please select all that apply.Â (choice=Your language or accent) |
| 406 | 6 | 1 | 0 | 0 | harascaus___14 | Numeric | What do you think are the reasons for these experiences? Please select all that apply.Â (choice=Your social class) |
| 407 | 6 | 1 | 0 | 0 | harascaus___15 | Numeric | What do you think are the reasons for these experiences? Please select all that apply.Â (choice=Other (please specify)) |
| 408 | 6 | 1 | 0 | 0 | harascaus___99 | Numeric | What do you think are the reasons for these experiences? Please select all that apply.Â (choice=Prefer not to answer) |
| 409 | 6 | 1 | 0 | 0 | haraswork___1 | Numeric | In the last 12 months have you personally experienced discrimination at work from any of the following? Select all that apply. (choice=Patients / service users, their relatives or other members of the public) |
| 410 | 6 | 1 | 0 | 0 | haraswork___2 | Numeric | In the last 12 months have you personally experienced discrimination at work from any of the following? Select all that apply. (choice=Manager / team leader or other colleagues) |
| 411 | 6 | 1 | 0 | 0 | haraswork___3 | Numeric | In the last 12 months have you personally experienced discrimination at work from any of the following? Select all that apply. (choice=I have not experienced discrimination at work in the last 12 months) |
| 412 | 6 | 1 | 0 | 0 | haraswork___4 | Numeric | In the last 12 months have you personally experienced discrimination at work from any of the following? Select all that apply. (choice=I have not worked in the last 12 months) |
| 413 | 6 | 1 | 0 | 0 | haraswork___99 | Numeric | In the last 12 months have you personally experienced discrimination at work from any of the following? Select all that apply. (choice=Prefer not to answer) |
| 414 | 6 | 1 | 0 | 0 | harasworkgr___1 | Numeric | On what grounds have you experienced discrimination at work? (choice=Your national origins) |
| 415 | 6 | 1 | 0 | 0 | harasworkgr___2 | Numeric | On what grounds have you experienced discrimination at work? (choice=Your gender) |
| 416 | 6 | 1 | 0 | 0 | harasworkgr___3 | Numeric | On what grounds have you experienced discrimination at work? (choice=Your ethnicity) |
| 417 | 6 | 1 | 0 | 0 | harasworkgr___4 | Numeric | On what grounds have you experienced discrimination at work? (choice=Your age) |
| 418 | 6 | 1 | 0 | 0 | harasworkgr___5 | Numeric | On what grounds have you experienced discrimination at work? (choice=Your religion) |
| 419 | 6 | 1 | 0 | 0 | harasworkgr___6 | Numeric | On what grounds have you experienced discrimination at work? (choice=Your height) |
| 420 | 6 | 1 | 0 | 0 | harasworkgr___7 | Numeric | On what grounds have you experienced discrimination at work? (choice=Your weight) |
| 421 | 6 | 1 | 0 | 0 | harasworkgr___8 | Numeric | On what grounds have you experienced discrimination at work? (choice=Your health or disability) |
| 422 | 6 | 1 | 0 | 0 | harasworkgr___9 | Numeric | On what grounds have you experienced discrimination at work? (choice=Your dress) |
| 423 | 6 | 1 | 0 | 0 | harasworkgr___10 | Numeric | On what grounds have you experienced discrimination at work? (choice=Some other aspect of your physical appearance) |
| 424 | 6 | 1 | 0 | 0 | harasworkgr___11 | Numeric | On what grounds have you experienced discrimination at work? (choice=Your sexual orientation) |
| 425 | 6 | 1 | 0 | 0 | harasworkgr___12 | Numeric | On what grounds have you experienced discrimination at work? (choice=Your education or income level) |
| 426 | 6 | 1 | 0 | 0 | harasworkgr___13 | Numeric | On what grounds have you experienced discrimination at work? (choice=Your language or accent) |
| 427 | 6 | 1 | 0 | 0 | harasworkgr___14 | Numeric | On what grounds have you experienced discrimination at work? (choice=Your social class) |
| 428 | 6 | 1 | 0 | 0 | harasworkgr___15 | Numeric | On what grounds have you experienced discrimination at work? (choice=Other (please specify)) |
| 429 | 6 | 1 | 0 | 0 | harasworkgr___99 | Numeric | On what grounds have you experienced discrimination at work? (choice=Prefer not to answer) |
| 430 | 6 | 1 | 0 | 0 | harasworkrs | String | Please specify the grounds on which you have experienced discrimination at work: |
| 431 | 6 | 1 | 0 | 0 | haras_comp | Numeric | Did you make a complaint about the discrimination at work?Â |
| 432 | 6 | 1 | 0 | 0 | harascaus_other | String | |
| 433 | 6 | 1 | 0 | 0 | harascaus_other_temp_Length | Numeric | |
| 434 | 7 | 0 | 0 | 1 | Section7_Raw_Health_____________Raw | Numeric | |
| 435 | 7 | 1 | 0 | 0 | hlthht | Numeric | What is your current height? |
| 436 | 7 | 1 | 0 | 0 | hlthhtcm | Numeric | Please enter your current height to the nearest centimetre: |
| 437 | 7 | 1 | 0 | 0 | hlthhtft | Numeric | Feet |
| 438 | 7 | 1 | 0 | 0 | hlthhtinch | Numeric | Inches |
| 439 | 7 | 1 | 0 | 0 | hlthwt | Numeric | What is your current weight? |
| 440 | 7 | 1 | 0 | 0 | hlthwtkg | Numeric | Please enter your current weight in kilograms: |
| 441 | 7 | 1 | 0 | 0 | hlthweightst | Numeric | Stones |
| 442 | 7 | 1 | 0 | 0 | hlthwtlb | Numeric | Pounds |
| 443 | 7 | 1 | 0 | 0 | hlthsmk | Numeric | Do you or have you ever smoked tobacco? |

| | | | | | | | |
|---|---|---|---|---|---|---|---|
| 444 | 7 | 1 | 0 | 0 | **hlthvape** | Numeric | Do you currently use an e-cigarette or vape? |
| 445 | 7 | 1 | 0 | 0 | **hlthalca** | Numeric | How often do you have a drink containing alcohol? |
| 446 | 7 | 1 | 0 | 0 | **hlthalcb** | Numeric | How many units of alcohol do you drink in a typical week? If you are unsure, see the guide below. Â Pint of standard strength (3.6%) lager/beer/cider 2 units Pint of higher strength (5.2%) lager/beer/cider 3 units Medium (175ml) glass of |
| 447 | 7 | 1 | 0 | 0 | **hlthgppaq1** | Numeric | Think about a typical week at work over the past month. Please consider the type and amount of physical activity involved in your work.Please select one option only. |
| 448 | 7 | 1 | 0 | 0 | **hlthgppaq2_1** | Numeric | Physical exercise such as swimming, jogging, aerobics, football, tennis, gym workout etc. |
| 449 | 7 | 1 | 0 | 0 | **hlthgppaq2_2** | Numeric | Cycling, including cycling to work and during leisure time |
| 450 | 7 | 1 | 0 | 0 | **hlthgppaq2_3** | Numeric | Walking, including walking to work, shopping, for pleasure etc. |
| 451 | 7 | 1 | 0 | 0 | **hlthgppaq2_4** | Numeric | Housework/Childcare |
| 452 | 7 | 1 | 0 | 0 | **hlthgppaq2_5** | Numeric | Gardening/DIY |
| 453 | 7 | 1 | 0 | 0 | **hlthgppaq3** | Numeric | How would you describe your usual walking pace? Please select one option only. |
| 454 | 7 | 1 | 0 | 0 | **hlthchange_1** | Numeric | Smoking |
| 455 | 7 | 1 | 0 | 0 | **hlthchange_2** | Numeric | Drinking alcohol |
| 456 | 7 | 1 | 0 | 0 | **hlthchange_3** | Numeric | Eating healthy food |
| 457 | 7 | 1 | 0 | 0 | **hlthchange_4** | Numeric | Physical activity (including walking and cycling) |
| 458 | 7 | 1 | 0 | 0 | **hlthvstgp** | Numeric | Last year, in 2019, how many times did you have a consultation with your GP about your own health? |
| 459 | 7 | 1 | 0 | 0 | **hlthinpt** | Numeric | Last year, in 2019, how many days did you spend as a hospital inpatient? |
| 460 | 7 | 1 | 0 | 0 | **hlthflujab** | Numeric | Did you have a flu vaccine last winter (2019-2020)? |
| 461 | 7 | 1 | 0 | 0 | **hlthflujab_2** | Numeric | Have you had a flu vaccine for this winter (2020-2021)? |
| 462 | 7 | 1 | 0 | 0 | **hlthshield** | Numeric | Have you been contacted by letter or text message to say you are at severe risk from COVID-19 due to an underlying health condition and should be shielding? |
| 463 | 7 | 1 | 0 | 0 | **hlthmeds___4** | Numeric | Do you currently take any of these medications/supplements?Please select all that apply. If you do not take any of these, please select None of these. (choice=Ibuprofen / Nurofen, any other type of non-steroidal anti-inflammatory) |
| 464 | 7 | 1 | 0 | 0 | **hlthmeds___5** | Numeric | Do you currently take any of these medications/supplements?Please select all that apply. If you do not take any of these, please select None of these. (choice=Vitamin D) |
| 465 | 7 | 1 | 0 | 0 | **hlthmeds___1** | Numeric | Do you currently take any of these medications/supplements?Please select all that apply. If you do not take any of these, please select None of these. (choice=ACE-inhibitor (e.g. ramipril, lisinopril)) |
| 466 | 7 | 1 | 0 | 0 | **hlthmeds___2** | Numeric | Do you currently take any of these medications/supplements?Please select all that apply. If you do not take any of these, please select None of these. (choice=Sartan (e.g. losartan, valsartan, candesartan)) |
| 467 | 7 | 1 | 0 | 0 | **hlthmeds___3** | Numeric | Do you currently take any of these medications/supplements?Please select all that apply. If you do not take any of these, please select None of these. (choice=Entresto (sucubitril/valsartan)) |
| 468 | 7 | 1 | 0 | 0 | **hlthmeds___6** | Numeric | Do you currently take any of these medications/supplements?Please select all that apply. If you do not take any of these, please select None of these. (choice=Metformin) |
| 469 | 7 | 1 | 0 | 0 | **hlthmeds___7** | Numeric | Do you currently take any of these medications/supplements?Please select all that apply. If you do not take any of these, please select None of these. (choice=None of these) |
| 470 | 7 | 1 | 0 | 0 | **hlthmeds___99** | Numeric | Do you currently take any of these medications/supplements?Please select all that apply. If you do not take any of these, please select None of these. (choice=Prefer not to answer) |
| 471 | 7 | 1 | 0 | 0 | **hlthcomorb___0** | Numeric | Are you, or do you, currently have any of the following?Please select all that apply. If none apply to you, please select None of the above. (choice=Pregnant) |
| 472 | 7 | 1 | 0 | 0 | **hlthcomorb___1** | Numeric | Are you, or do you, currently have any of the following?Please select all that apply. If none apply to you, please select None of the above. (choice=Organ transplant) |
| 473 | 7 | 1 | 0 | 0 | **hlthcomorb___2** | Numeric | Are you, or do you, currently have any of the following?Please select all that apply. If none apply to you, please select None of the above. (choice=Diabetes (Type I or II)) |
| 474 | 7 | 1 | 0 | 0 | **hlthcomorb___3** | Numeric | Are you, or do you, currently have any of the following?Please select all that apply. If none apply to you, please select None of the above. (choice=Heart disease or heart problems) |
| 475 | 7 | 1 | 0 | 0 | **hlthcomorb___4** | Numeric | Are you, or do you, currently have any of the following?Please select all that apply. If none apply to you, please select None of the above. (choice=Hypertension) |
| 476 | 7 | 1 | 0 | 0 | **hlthcomorb___5** | Numeric | Are you, or do you, currently have any of the following?Please select all that apply. If none apply to you, please select None of the above. (choice=Overweight) |
| 477 | 7 | 1 | 0 | 0 | **hlthcomorb___6** | Numeric | Are you, or do you, currently have any of the following?Please select all that apply. If none apply to you, please select None of the above. (choice=Stroke) |
| 478 | 7 | 1 | 0 | 0 | **hlthcomorb___7** | Numeric | Are you, or do you, currently have any of the following?Please select all that apply. If none apply to you, please select None of the above. (choice=Kidney disease) |
| 479 | 7 | 1 | 0 | 0 | **hlthcomorb___8** | Numeric | Are you, or do you, currently have any of the following?Please select all that apply. If none apply to you, please select None of the above. (choice=Liver disease) |
| 480 | 7 | 1 | 0 | 0 | **hlthcomorb___9** | Numeric | Are you, or do you, currently have any of the following?Please select all that apply. If none apply to you, please select None of the above. (choice=Anaemia) |
| 481 | 7 | 1 | 0 | 0 | **hlthcomorb___10** | Numeric | Are you, or do you, currently have any of the following?Please select all that apply. If none apply to you, please select None of the above. (choice=Asthma) |
| 482 | 7 | 1 | 0 | 0 | **hlthcomorb___11** | Numeric | Are you, or do you, currently have any of the following?Please select all that apply. If none apply to you, please select None of the above. (choice=Other lung condition such as COPD, bronchitis or emphysema) |
| 483 | 7 | 1 | 0 | 0 | **hlthcomorb___12** | Numeric | Are you, or do you, currently have any of the following?Please select all that apply. If none apply to you, please select None of the above. (choice=Cancer) |
| 484 | 7 | 1 | 0 | 0 | **hlthcomorb___13** | Numeric | Are you, or do you, currently have any of the following?Please select all that apply. If none apply to you, please select None of the above. (choice=Condition affecting the brain and nerves (e.g. Dementia, Parkinsons, Multiple Sclerosis)) |
| 485 | 7 | 1 | 0 | 0 | **hlthcomorb___14** | Numeric | Are you, or do you, currently have any of the following?Please select all that apply. If none apply to you, please select None of the above. (choice=A weakened immune system or reduced ability to deal with infections (as a result of a disease or treatment) |
| 486 | 7 | 1 | 0 | 0 | **hlthcomorb___15** | Numeric | Are you, or do you, currently have any of the following?Please select all that apply. If none apply to you, please select None of the above. (choice=Depression) |
| 487 | 7 | 1 | 0 | 0 | **hlthcomorb___16** | Numeric | Are you, or do you, currently have any of the following?Please select all that apply. If none apply to you, please select None of the above. (choice=Anxiety) |
| 488 | 7 | 1 | 0 | 0 | **hlthcomorb___17** | Numeric | Are you, or do you, currently have any of the following?Please select all that apply. If none apply to you, please select None of the above. (choice=Psychiatric disorder) |
| 489 | 7 | 1 | 0 | 0 | **hlthcomorb___18** | Numeric | Are you, or do you, currently have any of the following?Please select all that apply. If none apply to you, please select None of the above. (choice=None of the above) |
| 490 | 7 | 1 | 0 | 0 | **hlthcomorb___99** | Numeric | Are you, or do you, currently have any of the following?Please select all that apply. If none apply to you, please select None of the above. (choice=Prefer not to answer) |
| 491 | 7 | 1 | 0 | 0 | **healtheq5d_mob** | Numeric | MOBILITY |
| 492 | 7 | 1 | 0 | 0 | **hltheq5d_sc** | Numeric | SELF-CARE |
| 493 | 7 | 1 | 0 | 0 | **hltheq5d_ua** | Numeric | USUAL ACTIVITIESÂ (e.g. work, study, housework, family or leisure activities) |
| 494 | 7 | 1 | 0 | 0 | **hltheq5d_pd** | Numeric | Â Â Â PAIN / DISCOMFORT |
| 495 | 7 | 1 | 0 | 0 | **htlheq5d_ad** | Numeric | ANXIETY / DEPRESSION |
| 496 | 7 | 1 | 0 | 0 | **htltheq5d_t** | Numeric | We would like to know how good or bad your health is TODAY.This scale is numbered from 0 to 100. 100 means the best health you can imagine.0 means the worst health you can imagine.Using the slider, please indicate how your health is TODAY. |
| 497 | 7 | 1 | 0 | 0 | **hlthgad2_1x** | Numeric | Feeling nervous, anxious or on edge? |
| 498 | 7 | 1 | 0 | 0 | **hlthgad2_2x** | Numeric | Not being able to stop or control worrying? |
| 499 | 7 | 1 | 0 | 0 | **hlthphq2_pleasure** | Numeric | Little interest or pleasure in doing things |
| 500 | 7 | 1 | 0 | 0 | **hlthphq2_down** | Numeric | Feeling down, depressed, or hopeless? |
| 501 | 7 | 1 | 0 | 0 | **hlthfinance** | Numeric | How worried are you about your future financial situation? |
| 502 | 7 | 1 | 0 | 0 | **hlthptsd_1** | Numeric | Repeated, disturbing memories, thoughts, or images of a stressful experience from the past? |
| 503 | 7 | 1 | 0 | 0 | **hlthptsd_2** | Numeric | Feeling very upset when something reminded you of a stressful experience from the past? |
| 504 | 7 | 1 | 0 | 0 | **hlthptsd_3** | Numeric | Avoided activities or situations because they reminded you of a stressful experience from the past? |
| 505 | 7 | 1 | 0 | 0 | **hlthuclalonely_1** | Numeric | How often do you feel you lack companionship? |
| 506 | 7 | 1 | 0 | 0 | **hlthuclalonely_2** | Numeric | How often do you feel left out? |
| 507 | 7 | 1 | 0 | 0 | **hlthuclalonely_3** | Numeric | How often do you feel isolated from others? |

| | | | | | | | |
|---|---|---|---|---|---|---|---|
| 508 | 7 | 1 | 0 | 0 | **hlthsat_1** | Numeric | Overall, how satisfied are you with your life nowadays?Please give an answer on a scale of 0 to 10, where 0 is not at all and 10 is completely. |
| 509 | 8 | 0 | 0 | 1 | **Section8_Raw_Covid_______________Raw** | Numeric | |
| 510 | 8 | 1 | 0 | 0 | **c19contact** | Numeric | Have you been in close contact with anyone with COVID-19 outside of your work in the last two weeks? |
| 511 | 8 | 1 | 0 | 0 | **c19_behav_1___1** | Numeric | Cancelling my usual social activities (choice=Between January 2020and March 2020) |
| 512 | 8 | 1 | 0 | 0 | **c19_behav_1___2** | Numeric | Cancelling my usual social activities (choice=Past few weeks) |
| 513 | 8 | 1 | 0 | 0 | **c19_behav_1___99** | Numeric | Cancelling my usual social activities (choice=Prefer not to answer) |
| 514 | 8 | 1 | 0 | 0 | **c19_behav_2___1** | Numeric | Not going to work (choice=Between January 2020and March 2020) |
| 515 | 8 | 1 | 0 | 0 | **c19_behav_2___2** | Numeric | Not going to work (choice=Past few weeks) |
| 516 | 8 | 1 | 0 | 0 | **c19_behav_2___99** | Numeric | Not going to work (choice=Prefer not to answer) |
| 517 | 8 | 1 | 0 | 0 | **c19_behav_3___1** | Numeric | Only going shopping for essential things (choice=Between January 2020and March 2020) |
| 518 | 8 | 1 | 0 | 0 | **c19_behav_3___2** | Numeric | Only going shopping for essential things (choice=Past few weeks) |
| 519 | 8 | 1 | 0 | 0 | **c19_behav_3___99** | Numeric | Only going shopping for essential things (choice=Prefer not to answer) |
| 520 | 8 | 1 | 0 | 0 | **c19_behav_4___1** | Numeric | Not going to a grocery store or pharmacy (choice=Between January 2020and March 2020) |
| 521 | 8 | 1 | 0 | 0 | **c19_behav_4___2** | Numeric | Not going to a grocery store or pharmacy (choice=Past few weeks) |
| 522 | 8 | 1 | 0 | 0 | **c19_behav_4___99** | Numeric | Not going to a grocery store or pharmacy (choice=Prefer not to answer) |
| 523 | 8 | 1 | 0 | 0 | **c19_behav_5___1** | Numeric | Not leaving the house (choice=Between January 2020and March 2020) |
| 524 | 8 | 1 | 0 | 0 | **c19_behav_5___2** | Numeric | Not leaving the house (choice=Past few weeks) |
| 525 | 8 | 1 | 0 | 0 | **c19_behav_5___99** | Numeric | Not leaving the house (choice=Prefer not to answer) |
| 526 | 8 | 1 | 0 | 0 | **c19_behav_6___1** | Numeric | Wearing a face mask outside my home (choice=Between January 2020and March 2020) |
| 527 | 8 | 1 | 0 | 0 | **c19_behav_6___2** | Numeric | Wearing a face mask outside my home (choice=Past few weeks) |
| 528 | 8 | 1 | 0 | 0 | **c19_behav_6___99** | Numeric | Wearing a face mask outside my home (choice=Prefer not to answer) |
| 529 | 8 | 1 | 0 | 0 | **c19_behav_7___1** | Numeric | Trying to avoid physical contact with people (choice=Between January 2020and March 2020) |
| 530 | 8 | 1 | 0 | 0 | **c19_behav_7___2** | Numeric | Trying to avoid physical contact with people (choice=Past few weeks) |
| 531 | 8 | 1 | 0 | 0 | **c19_behav_7___99** | Numeric | Trying to avoid physical contact with people (choice=Prefer not to answer) |
| 532 | 8 | 1 | 0 | 0 | **c19_behav_8___1** | Numeric | Following handwashing recommendations (choice=Between January 2020and March 2020) |
| 533 | 8 | 1 | 0 | 0 | **c19_behav_8___2** | Numeric | Following handwashing recommendations (choice=Past few weeks) |
| 534 | 8 | 1 | 0 | 0 | **c19_behav_8___99** | Numeric | Following handwashing recommendations (choice=Prefer not to answer) |
| 535 | 8 | 1 | 0 | 0 | **c19_behav_9___1** | Numeric | Using hand sanitiser more than usual (choice=Between January 2020and March 2020) |
| 536 | 8 | 1 | 0 | 0 | **c19_behav_9___2** | Numeric | Using hand sanitiser more than usual (choice=Past few weeks) |
| 537 | 8 | 1 | 0 | 0 | **c19_behav_9___99** | Numeric | Using hand sanitiser more than usual (choice=Prefer not to answer) |
| 538 | 8 | 1 | 0 | 0 | **c19_behav_10___1** | Numeric | Following coughing and sneezing recommendations (choice=Between January 2020and March 2020) |
| 539 | 8 | 1 | 0 | 0 | **c19_behav_10___2** | Numeric | Following coughing and sneezing recommendations (choice=Past few weeks) |
| 540 | 8 | 1 | 0 | 0 | **c19_behav_10___99** | Numeric | Following coughing and sneezing recommendations (choice=Prefer not to answer) |
| 541 | 8 | 1 | 0 | 0 | **c19_behav_11___1** | Numeric | Using tissues more than usual (choice=Between January 2020and March 2020) |
| 542 | 8 | 1 | 0 | 0 | **c19_behav_11___2** | Numeric | Using tissues more than usual (choice=Past few weeks) |
| 543 | 8 | 1 | 0 | 0 | **c19_behav_11___99** | Numeric | Using tissues more than usual (choice=Prefer not to answer) |
| 544 | 8 | 1 | 0 | 0 | **c19_behav_12___1** | Numeric | Wearing gloves while going out of my home (choice=Between January 2020and March 2020) |
| 545 | 8 | 1 | 0 | 0 | **c19_behav_12___2** | Numeric | Wearing gloves while going out of my home (choice=Past few weeks) |
| 546 | 8 | 1 | 0 | 0 | **c19_behav_12___99** | Numeric | Wearing gloves while going out of my home (choice=Prefer not to answer) |
| 547 | 8 | 1 | 0 | 0 | **c19_behav_13___1** | Numeric | Avoiding public transport (choice=Between January 2020and March 2020) |
| 548 | 8 | 1 | 0 | 0 | **c19_behav_13___2** | Numeric | Avoiding public transport (choice=Past few weeks) |
| 549 | 8 | 1 | 0 | 0 | **c19_behav_13___99** | Numeric | Avoiding public transport (choice=Prefer not to answer) |
| 550 | 8 | 1 | 0 | 0 | **c19_behav_14___1** | Numeric | Avoiding going to restaurants/bars/pubs (choice=Between January 2020and March 2020) |
| 551 | 8 | 1 | 0 | 0 | **c19_behav_14___2** | Numeric | Avoiding going to restaurants/bars/pubs (choice=Past few weeks) |
| 552 | 8 | 1 | 0 | 0 | **c19_behav_14___99** | Numeric | Avoiding going to restaurants/bars/pubs (choice=Prefer not to answer) |
| 553 | 8 | 1 | 0 | 0 | **c19_behav_15___1** | Numeric | Avoiding going for walks or exercise outside (choice=Between January 2020and March 2020) |
| 554 | 8 | 1 | 0 | 0 | **c19_behav_15___2** | Numeric | Avoiding going for walks or exercise outside (choice=Past few weeks) |
| 555 | 8 | 1 | 0 | 0 | **c19_behav_15___99** | Numeric | Avoiding going for walks or exercise outside (choice=Prefer not to answer) |
| 556 | 8 | 1 | 0 | 0 | **c19_behav_16___1** | Numeric | Avoiding taking my children out of my home (if applicable) (choice=Between January 2020and March 2020) |
| 557 | 8 | 1 | 0 | 0 | **c19_behav_16___2** | Numeric | Avoiding taking my children out of my home (if applicable) (choice=Past few weeks) |
| 558 | 8 | 1 | 0 | 0 | **c19_behav_16___99** | Numeric | Avoiding taking my children out of my home (if applicable) (choice=Prefer not to answer) |
| 559 | 8 | 1 | 0 | 0 | **c19_behav_17___1** | Numeric | Prefer not to answer (choice=Between January 2020and March 2020) |
| 560 | 8 | 1 | 0 | 0 | **c19_behav_17___2** | Numeric | Prefer not to answer (choice=Past few weeks) |
| 561 | 8 | 1 | 0 | 0 | **c19_behav_17___99** | Numeric | Prefer not to answer (choice=Prefer not to answer) |
| 562 | 8 | 1 | 0 | 0 | **lockdownresponse** | Numeric | Thinking back to the months of UK national lockdown which began on 23rd March, which of these is closest to your view? |
| 563 | 8 | 1 | 0 | 0 | **c19typetest___1** | Numeric | Have you ever had a test to see if you have or have had COVID-19? Select all that apply. (choice=No) |
| 564 | 8 | 1 | 0 | 0 | **c19typetest___2** | Numeric | Have you ever had a test to see if you have or have had COVID-19? Select all that apply. (choice=Yes, A swab test (swab of your throat and/or nose) which tests for active infection) |
| 565 | 8 | 1 | 0 | 0 | **c19typetest___3** | Numeric | Have you ever had a test to see if you have or have had COVID-19? Select all that apply. (choice=Yes, An antibody test for COVID-19 (a blood test, or a drop of blood from your finger) which tests for past infection) |
| 566 | 8 | 1 | 0 | 0 | **c19typetest___4** | Numeric | Have you ever had a test to see if you have or have had COVID-19? Select all that apply. (choice=Do not know) |
| 567 | 8 | 1 | 0 | 0 | **c19typetest___99** | Numeric | Have you ever had a test to see if you have or have had COVID-19? Select all that apply. (choice=Prefer not to answer) |
| 568 | 8 | 1 | 0 | 0 | **c19evertest___1** | Numeric | What was the reason that you had the swab test? Please select all that apply. (choice=Because I had symptoms) |
| 569 | 8 | 1 | 0 | 0 | **c19evertest___2** | Numeric | What was the reason that you had the swab test? Please select all that apply. (choice=Because I have been in contact with someone who had COVID-19) |
| 570 | 8 | 1 | 0 | 0 | **c19evertest___3** | Numeric | What was the reason that you had the swab test? Please select all that apply. (choice=Because of my job) |
| 571 | 8 | 1 | 0 | 0 | **c19evertest___4** | Numeric | What was the reason that you had the swab test? Please select all that apply. (choice=Before going into hospital as a patient (e.g. for surgery)) |

| | | | | | | | | | | | |
|---|---|---|---|---|---|---|---|---|---|---|---|
| 572 | 8 | 1 | 0 | 0 | c19evertest___5 | | | | | Numeric | What was the reason that you had the swab test? Please select all that apply. (choice=It was offered by my local council (e.g. posted through my door)) |
| 573 | 8 | 1 | 0 | 0 | c19evertest___6 | | | | | Numeric | What was the reason that you had the swab test? Please select all that apply. (choice=For another reason (please specify)) |
| 574 | 8 | 1 | 0 | 0 | c19evertest___99 | | | | | Numeric | What was the reason that you had the swab test? Please select all that apply. (choice=Prefer not to answer) |
| 575 | 8 | 1 | 0 | 0 | c19evertest_other | | | | | String | Please specify the reason you had the swab test: |
| 576 | 8 | 1 | 0 | 0 | c19swabres | | | | | Numeric | Have you ever had a positive result from a swab test (i.e. showing that you had coronavirus when the swab was taken)? |
| 577 | 8 | 1 | 0 | 0 | c19swabdate | | | | | String | When was the sample taken for the test that came back positive?Give the latest date if you have had more than one |
| 578 | 8 | 1 | 0 | 0 | c19abres | | | | | Numeric | Have you had a positive result from an antibody test (i.e. showing that you had coronavirus at some time previously)? |
| 579 | 8 | 1 | 0 | 0 | c19abdate | | | | | String | When was the sample taken for the test that came back positive?Give the latest date if you have had more than one |
| 580 | 8 | 1 | 0 | 0 | c19sympt___1 | | | | | Numeric | Have you experienced any of the following symptoms in the past two weeks?Please select all that apply. (choice=Fever) |
| 581 | 8 | 1 | 0 | 0 | c19sympt___2 | | | | | Numeric | Have you experienced any of the following symptoms in the past two weeks?Please select all that apply. (choice=Cough - dry) |
| 582 | 8 | 1 | 0 | 0 | c19sympt___3 | | | | | Numeric | Have you experienced any of the following symptoms in the past two weeks?Please select all that apply. (choice=Cough - mucus or phlegm) |
| 583 | 8 | 1 | 0 | 0 | c19sympt___4 | | | | | Numeric | Have you experienced any of the following symptoms in the past two weeks?Please select all that apply. (choice=Sore throat) |
| 584 | 8 | 1 | 0 | 0 | c19sympt___5 | | | | | Numeric | Have you experienced any of the following symptoms in the past two weeks?Please select all that apply. (choice=Chest tightness) |
| 585 | 8 | 1 | 0 | 0 | c19sympt___6 | | | | | Numeric | Have you experienced any of the following symptoms in the past two weeks?Please select all that apply. (choice=Shortness of breath) |
| 586 | 8 | 1 | 0 | 0 | c19sympt___7 | | | | | Numeric | Have you experienced any of the following symptoms in the past two weeks?Please select all that apply. (choice=Runny nose) |
| 587 | 8 | 1 | 0 | 0 | c19sympt___8 | | | | | Numeric | Have you experienced any of the following symptoms in the past two weeks?Please select all that apply. (choice=Nasal congestion) |
| 588 | 8 | 1 | 0 | 0 | c19sympt___9 | | | | | Numeric | Have you experienced any of the following symptoms in the past two weeks?Please select all that apply. (choice=Sneezing) |
| 589 | 8 | 1 | 0 | 0 | c19sympt___10 | | | | | Numeric | Have you experienced any of the following symptoms in the past two weeks?Please select all that apply. (choice=Muscle or body aches) |
| 590 | 8 | 1 | 0 | 0 | c19sympt___11 | | | | | Numeric | Have you experienced any of the following symptoms in the past two weeks?Please select all that apply. (choice=Fatigue) |
| 591 | 8 | 1 | 0 | 0 | c19sympt___12 | | | | | Numeric | Have you experienced any of the following symptoms in the past two weeks?Please select all that apply. (choice=Unusual loose motions or diarrhoea) |
| 592 | 8 | 1 | 0 | 0 | c19sympt___13 | | | | | Numeric | Have you experienced any of the following symptoms in the past two weeks?Please select all that apply. (choice=Vomiting) |
| 593 | 8 | 1 | 0 | 0 | c19sympt___14 | | | | | Numeric | Have you experienced any of the following symptoms in the past two weeks?Please select all that apply. (choice=Loss of smell) |
| 594 | 8 | 1 | 0 | 0 | c19sympt___15 | | | | | Numeric | Have you experienced any of the following symptoms in the past two weeks?Please select all that apply. (choice=Loss of taste) |
| 595 | 8 | 1 | 0 | 0 | c19sympt___16 | | | | | Numeric | Have you experienced any of the following symptoms in the past two weeks?Please select all that apply. (choice=Skin rash) |
| 596 | 8 | 1 | 0 | 0 | c19sympt___17 | | | | | Numeric | Have you experienced any of the following symptoms in the past two weeks?Please select all that apply. (choice=None of these) |
| 597 | 8 | 1 | 0 | 0 | c19sympt___99 | | | | | Numeric | Have you experienced any of the following symptoms in the past two weeks?Please select all that apply. (choice=Prefer not to answer) |
| 598 | 8 | 1 | 0 | 0 | c19ever | | | | | Numeric | Do you think that you currently have or have had COVID-19? |
| 599 | 8 | 1 | 0 | 0 | c19date_month | | | | | Numeric | Month |
| 600 | 8 | 1 | 0 | 0 | c19date_year | | | | | Numeric | Year |
| 601 | 8 | 1 | 0 | 0 | c19hosp | | | | | Numeric | Were you hospitalised for treatment of your COVID-19 disease? |
| 602 | 8 | 1 | 0 | 0 | c19icu | | | | | Numeric | Were you admitted to the intensive care unit (ITU/ICU)? |
| 603 | 8 | 1 | 0 | 0 | c19_long | | | | | Numeric | How long have you been unwell since having COVID-19? |
| 604 | 8 | 1 | 0 | 0 | c19_long_diag | | | | | Numeric | Has any health professional told you that your symptoms are likely to be the ongoing effect of COVID-19? |
| 605 | 8 | 1 | 0 | 0 | c19crnsl | | | | | Numeric | How concerned are you that you will get COVID-19 and require hospitalisation? |
| 606 | 8 | 1 | 0 | 0 | c19crnot | | | | | Numeric | How concerned are you that you might unknowingly spread COVID-19 to others? |
| 607 | 8 | 1 | 0 | 0 | c19died___1 | | | | | Numeric | Do you personally know anyone who has died from COVID-19 (not including patients you have cared for as part of your work)?Select all that apply. (choice=Yes, family member(s)) |
| 608 | 8 | 1 | 0 | 0 | c19died___2 | | | | | Numeric | Do you personally know anyone who has died from COVID-19 (not including patients you have cared for as part of your work)?Select all that apply. (choice=Yes, friend(s)) |
| 609 | 8 | 1 | 0 | 0 | c19died___3 | | | | | Numeric | Do you personally know anyone who has died from COVID-19 (not including patients you have cared for as part of your work)?Select all that apply. (choice=Yes, colleague(s)) |
| 610 | 8 | 1 | 0 | 0 | c19died___4 | | | | | Numeric | Do you personally know anyone who has died from COVID-19 (not including patients you have cared for as part of your work)?Select all that apply. (choice=Yes, someone else) |
| 611 | 8 | 1 | 0 | 0 | c19died___5 | | | | | Numeric | Do you personally know anyone who has died from COVID-19 (not including patients you have cared for as part of your work)?Select all that apply. (choice=No) |
| 612 | 8 | 1 | 0 | 0 | c19died___99 | | | | | Numeric | Do you personally know anyone who has died from COVID-19 (not including patients you have cared for as part of your work)?Select all that apply. (choice=Prefer not to answer) |
| 613 | 8 | 1 | 0 | 0 | c19info___1 | | | | | Numeric | Where do you get information about COVID-19?Select all that apply. (choice=Friends, family, neighbours) |
| 614 | 8 | 1 | 0 | 0 | c19info___2 | | | | | Numeric | Where do you get information about COVID-19?Select all that apply. (choice=Colleagues) |
| 615 | 8 | 1 | 0 | 0 | c19info___3 | | | | | Numeric | Where do you get information about COVID-19?Select all that apply. (choice=Employer or manager) |
| 616 | 8 | 1 | 0 | 0 | c19info___4 | | | | | Numeric | Where do you get information about COVID-19?Select all that apply. (choice=Television) |
| 617 | 8 | 1 | 0 | 0 | c19info___5 | | | | | Numeric | Where do you get information about COVID-19?Select all that apply. (choice=Radio) |
| 618 | 8 | 1 | 0 | 0 | c19info___6 | | | | | Numeric | Where do you get information about COVID-19?Select all that apply. (choice=Newspapers or magazines) |
| 619 | 8 | 1 | 0 | 0 | c19info___7 | | | | | Numeric | Where do you get information about COVID-19?Select all that apply. (choice=Government or NHS posters, adverts or leaflets) |
| 620 | 8 | 1 | 0 | 0 | c19info___8 | | | | | Numeric | Where do you get information about COVID-19?Select all that apply. (choice=Twitter) |
| 621 | 8 | 1 | 0 | 0 | c19info___9 | | | | | Numeric | Where do you get information about COVID-19?Select all that apply. (choice=Other social media (e.g. Facebook, Instagram)) |
| 622 | 8 | 1 | 0 | 0 | c19info___10 | | | | | Numeric | Where do you get information about COVID-19?Select all that apply. (choice=UK Government website) |
| 623 | 8 | 1 | 0 | 0 | c19info___11 | | | | | Numeric | Where do you get information about COVID-19?Select all that apply. (choice=Welsh, Scottish or NI government website) |
| 624 | 8 | 1 | 0 | 0 | c19info___12 | | | | | Numeric | Where do you get information about COVID-19?Select all that apply. (choice=NHS website) |
| 625 | 8 | 1 | 0 | 0 | c19info___13 | | | | | Numeric | Where do you get information about COVID-19?Select all that apply. (choice=WHO website) |
| 626 | 8 | 1 | 0 | 0 | c19info___14 | | | | | Numeric | Where do you get information about COVID-19?Select all that apply. (choice=Other websites) |
| 627 | 8 | 1 | 0 | 0 | c19info___15 | | | | | Numeric | Where do you get information about COVID-19?Select all that apply. (choice=Local council) |
| 628 | 8 | 1 | 0 | 0 | c19info___16 | | | | | Numeric | Where do you get information about COVID-19?Select all that apply. (choice=My own GP or other healthcare workers) |
| 629 | 8 | 1 | 0 | 0 | c19info___17 | | | | | Numeric | Where do you get information about COVID-19?Select all that apply. (choice=Scientific journals) |
| 630 | 8 | 1 | 0 | 0 | c19info___18 | | | | | Numeric | Where do you get information about COVID-19?Select all that apply. (choice=Not applicable) |
| 631 | 8 | 1 | 0 | 0 | c19info___99 | | | | | Numeric | Where do you get information about COVID-19?Select all that apply. (choice=Prefer not to answer) |
| 632 | 8 | 1 | 0 | 0 | c19riskpersmonth | | | | | Numeric | What do you think is your personal chance of catching the coronavirus in the next month?Please enter a value on a scale from 0 to 100, where 0 means there is no possibility that you will and 100 means that you definitely will. |
| 633 | 8 | 1 | 0 | 0 | c19riskpers6months | | | | | Numeric | What do you think is your personal chance of catching the coronavirus in the next six months?Please enter a value on a scale from 0 to 100, where 0 means there is no possibility that you will and 100 means that you definitely will. |
| 634 | 8 | 1 | 0 | 0 | c19riskpershosp | 1 | 1 | 7 | 51 | Numeric | If you do catch coronavirus, what do you think areÂ yourÂ chances of needing hospital treatment? Please enter a value on a scale from 0 to 100, where 0 means there is no possibility that you will and 100 means that you definitely will. |
| 635 | 8 | 1 | 0 | 0 | c19riskukdeath | | | | | Numeric | What percentage of people in the UK who are hospitalised with coronavirus do you think will end up dying as a result of the disease? Please enter a value on a scale from 0 to 100, where 0 means nobody hospitalised will die and 100 means everybody hospitali |

| | | | | | | | | | |
|---|---|---|---|---|---|---|---|---|---|
| 636 | 8 | 1 | 0 | 0 | c19vaccn1 | | | Numeric | Have you taken part inÂ a trialÂ of a COVID-19 vaccine? |
| 637 | 8 | 1 | 0 | 0 | c19vaccn1txt | | | String | If yes, which one? |
| 638 | 8 | 1 | 0 | 0 | c19vaccn2 | | | Numeric | Have youÂ had, or are youÂ going to have,Â a vaccination against COVID-19? |
| 639 | 8 | 1 | 0 | 0 | c19vaccn2dose | | | Numeric | How many doses have you had? |
| 640 | 8 | 1 | 0 | 0 | c19vaccn2loc | | | Numeric | Was the vaccination: |
| 641 | 8 | 1 | 0 | 0 | c19vaccn2loctxt | | | String | Please specify: |
| 642 | 8 | 1 | 0 | 0 | c19vaccn2date | | | String | What was the date when you had your first vaccination? (if you are unsure please give your best estimate) |
| 643 | 8 | 1 | 0 | 0 | c19vaccn2which | | | Numeric | Which vaccine did you receive? |
| 644 | 8 | 1 | 0 | 0 | c19vaccn2whichtxt | | | String | Please specify: |
| 645 | 8 | 1 | 0 | 0 | c19vaccn2cons | | | Numeric | Did you considerÂ notÂ having the vaccination? |
| 646 | 8 | 1 | 0 | 0 | c19vaccn2reas___1 | | | Numeric | What would have been your reason(s) for not having the vaccination? Please select all that apply. (choice=I have allergies, needle-phobia, am immuno-compromised, or have other clinical reasons not to be vaccinated) |
| 647 | 8 | 1 | 0 | 0 | c19vaccn2reas___2 | | | Numeric | What would have been your reason(s) for not having the vaccination? Please select all that apply. (choice=I am concerned about the safety or potential side-effects of a COVID-19 vaccine) |
| 648 | 8 | 1 | 0 | 0 | c19vaccn2reas___3 | | | Numeric | What would have been your reason(s) for not having the vaccination? Please select all that apply. (choice=I am not convinced that COVID-19 vaccines will be effective) |
| 649 | 8 | 1 | 0 | 0 | c19vaccn2reas___4 | | | Numeric | What would have been your reason(s) for not having the vaccination? Please select all that apply. (choice=Vaccines may not have been tested thoroughly in all ethnic groups) |
| 650 | 8 | 1 | 0 | 0 | c19vaccn2reas___5 | | | Numeric | What would have been your reason(s) for not having the vaccination? Please select all that apply. (choice=I have had COVID-19 and therefore do not feel I need the vaccine) |
| 651 | 8 | 1 | 0 | 0 | c19vaccn2reas___6 | | | Numeric | What would have been your reason(s) for not having the vaccination? Please select all that apply. (choice=I am taking part in a clinical trial of a COVID-19 vaccine) |
| 652 | 8 | 1 | 0 | 0 | c19vaccn2reas___7 | | | Numeric | What would have been your reason(s) for not having the vaccination? Please select all that apply. (choice=I would prefer one of the other COVID-19 vaccines that are being developed) |
| 653 | 8 | 1 | 0 | 0 | c19vaccn2reas___8 | | | Numeric | What would have been your reason(s) for not having the vaccination? Please select all that apply. (choice=I would prefer to wait until many other people have received a COVID-19 vaccine) |
| 654 | 8 | 1 | 0 | 0 | c19vaccn2reas___9 | | | Numeric | What would have been your reason(s) for not having the vaccination? Please select all that apply. (choice=I do not feel that I personally am at risk from COVID-19) |
| 655 | 8 | 1 | 0 | 0 | c19vaccn2reas___10 | | | Numeric | What would have been your reason(s) for not having the vaccination? Please select all that apply. (choice=I would rather the vaccine were used for other people who need it more than I do) |
| 656 | 8 | 1 | 0 | 0 | c19vaccn2reas___11 | | | Numeric | What would have been your reason(s) for not having the vaccination? Please select all that apply. (choice=I do not believe in vaccinations in general) |
| 657 | 8 | 1 | 0 | 0 | c19vaccn2reas___12 | | | Numeric | What would have been your reason(s) for not having the vaccination? Please select all that apply. (choice=Other reason) |
| 658 | 8 | 1 | 0 | 0 | c19vaccn2reas___99 | | | Numeric | What would have been your reason(s) for not having the vaccination? Please select all that apply. (choice=Prefer not to answer) |
| 659 | 8 | 1 | 0 | 0 | c19vaccn2reastxt | | | String | Please specify: |
| 660 | 8 | 1 | 0 | 0 | c19vaccn2when | | | Numeric | When is the vaccination likely to be? |
| 661 | 8 | 1 | 0 | 0 | c19vaccn2whentxt | | | String | Please specify: |
| 662 | 8 | 1 | 0 | 0 | c19vaccn2loc2 | | | Numeric | Will this vaccination be: |
| 663 | 8 | 1 | 0 | 0 | c19vaccn2loc2txt | | | String | Please specify: |
| 664 | 8 | 1 | 0 | 0 | c19vaccn2cons_2 | | | Numeric | Are you considering not having the vaccination? |
| 665 | 8 | 1 | 0 | 0 | c19vaccn2reas2___1 | | | Numeric | What might be your reason(s) for not having the vaccination? Please select all that apply. (choice=I have allergies, needle-phobia, am immuno-compromised, or have other clinical reasons not to be vaccinated) |
| 666 | 8 | 1 | 0 | 0 | c19vaccn2reas2___2 | | | Numeric | What might be your reason(s) for not having the vaccination? Please select all that apply. (choice=I am concerned about the safety or potential side-effects of a COVID-19 vaccine) |
| 667 | 8 | 1 | 0 | 0 | c19vaccn2reas2___3 | | | Numeric | What might be your reason(s) for not having the vaccination? Please select all that apply. (choice=I am not convinced that COVID-19 vaccines will be effective) |
| 668 | 8 | 1 | 0 | 0 | c19vaccn2reas2___4 | | | Numeric | What might be your reason(s) for not having the vaccination? Please select all that apply. (choice=Vaccines may not have been tested thoroughly in all ethnic groups) |
| 669 | 8 | 1 | 0 | 0 | c19vaccn2reas2___5 | | | Numeric | What might be your reason(s) for not having the vaccination? Please select all that apply. (choice=I have had COVID-19 and therefore do not feel I need the vaccine) |
| 670 | 8 | 1 | 0 | 0 | c19vaccn2reas2___6 | | | Numeric | What might be your reason(s) for not having the vaccination? Please select all that apply. (choice=I am taking part in a clinical trial of a COVID-19 vaccine) |
| 671 | 8 | 1 | 0 | 0 | c19vaccn2reas2___7 | | | Numeric | What might be your reason(s) for not having the vaccination? Please select all that apply. (choice=I would prefer one of the other COVID-19 vaccines that are being developed) |
| 672 | 8 | 1 | 0 | 0 | c19vaccn2reas2___8 | | | Numeric | What might be your reason(s) for not having the vaccination? Please select all that apply. (choice=I would prefer to wait until many other people have received a COVID-19 vaccine) |
| 673 | 8 | 1 | 0 | 0 | c19vaccn2reas2___9 | | | Numeric | What might be your reason(s) for not having the vaccination? Please select all that apply. (choice=I do not feel that I personally am at risk from COVID-19) |
| 674 | 8 | 1 | 0 | 0 | c19vaccn2reas2___10 | | | Numeric | What might be your reason(s) for not having the vaccination? Please select all that apply. (choice=I would rather the vaccine were used for other people who need it more than I do) |
| 675 | 8 | 1 | 0 | 0 | c19vaccn2reas2___11 | | | Numeric | What might be your reason(s) for not having the vaccination? Please select all that apply. (choice=I do not believe in vaccinations in general) |
| 676 | 8 | 1 | 0 | 0 | c19vaccn2reas2___12 | | | Numeric | What might be your reason(s) for not having the vaccination? Please select all that apply. (choice=Other reason) |
| 677 | 8 | 1 | 0 | 0 | c19vaccn2reas2___99 | | | Numeric | What might be your reason(s) for not having the vaccination? Please select all that apply. (choice=Prefer not to answer) |
| 678 | 8 | 1 | 0 | 0 | c19vaccn2reas2txt | | | String | Please specify: |
| 679 | 8 | 1 | 0 | 0 | c19vaccn2loc3 | | | Numeric | Was the vaccination offered by: |
| 680 | 8 | 1 | 0 | 0 | c19vaccn2loc3txt | | | String | Please specify:Â |
| 681 | 8 | 1 | 0 | 0 | c19vaccn2reas3___1 | | | Numeric | What were your reason(s) for not having the vaccination? Please select all that apply. (choice=I have allergies, needle-phobia, am immuno-compromised, or have other clinical reasons not to be vaccinated) |
| 682 | 8 | 1 | 0 | 0 | c19vaccn2reas3___2 | | | Numeric | What were your reason(s) for not having the vaccination? Please select all that apply. (choice=I am concerned about the safety or potential side-effects of a COVID-19 vaccine) |
| 683 | 8 | 1 | 0 | 0 | c19vaccn2reas3___3 | | | Numeric | What were your reason(s) for not having the vaccination? Please select all that apply. (choice=I am not convinced that COVID-19 vaccines will be effective) |
| 684 | 8 | 1 | 0 | 0 | c19vaccn2reas3___4 | | | Numeric | What were your reason(s) for not having the vaccination? Please select all that apply. (choice=Vaccines may not have been tested thoroughly in all ethnic groups) |
| 685 | 8 | 1 | 0 | 0 | c19vaccn2reas3___5 | | | Numeric | What were your reason(s) for not having the vaccination? Please select all that apply. (choice=I have had COVID-19 and therefore do not feel I need the vaccine) |
| 686 | 8 | 1 | 0 | 0 | c19vaccn2reas3___6 | | | Numeric | What were your reason(s) for not having the vaccination? Please select all that apply. (choice=I am taking part in a clinical trial of a COVID-19 vaccine) |
| 687 | 8 | 1 | 0 | 0 | c19vaccn2reas3___7 | | | Numeric | What were your reason(s) for not having the vaccination? Please select all that apply. (choice=I would prefer one of the other COVID-19 vaccines that are being developed) |
| 688 | 8 | 1 | 0 | 0 | c19vaccn2reas3___8 | | | Numeric | What were your reason(s) for not having the vaccination? Please select all that apply. (choice=I would prefer to wait until many other people have received a COVID-19 vaccine) |
| 689 | 8 | 1 | 0 | 0 | c19vaccn2reas3___9 | | | Numeric | What were your reason(s) for not having the vaccination? Please select all that apply. (choice=I do not feel that I personally am at risk from COVID-19) |
| 690 | 8 | 1 | 0 | 0 | c19vaccn2reas3___10 | | | Numeric | What were your reason(s) for not having the vaccination? Please select all that apply. (choice=I would rather the vaccine were used for other people who need it more than I do) |
| 691 | 8 | 1 | 0 | 0 | c19vaccn2reas3___11 | | | Numeric | What were your reason(s) for not having the vaccination? Please select all that apply. (choice=I do not believe in vaccinations in general) |
| 692 | 8 | 1 | 0 | 0 | c19vaccn2reas3___12 | | | Numeric | What were your reason(s) for not having the vaccination? Please select all that apply. (choice=Other reason) |
| 693 | 8 | 1 | 0 | 0 | c19vaccn2reas3___99 | | | Numeric | What were your reason(s) for not having the vaccination? Please select all that apply. (choice=Prefer not to answer) |
| 694 | 8 | 1 | 0 | 0 | c19vaccn2reas3txt | | | String | Please specify: |
| 695 | 8 | 1 | 0 | 0 | c19vaccn2cons_4 | | | Numeric | When you are offered the vaccine, is there anything that might make you consider not having it? |
| 696 | 8 | 1 | 0 | 0 | c19vaccn2reas4___1 | | | Numeric | What are your reason(s) for considering not having the vaccine? Please select all that apply. (choice=I have allergies, needle-phobia, am immuno-compromised, or have other clinical reasons not to be vaccinated) |
| 697 | 8 | 1 | 0 | 0 | c19vaccn2reas4___2 | | | Numeric | What are your reason(s) for considering not having the vaccine? Please select all that apply. (choice=I am concerned about the safety or potential side-effects of a COVID-19 vaccine) |
| 698 | 8 | 1 | 0 | 0 | c19vaccn2reas4___3 | | | Numeric | What are your reason(s) for considering not having the vaccine? Please select all that apply. (choice=I am not convinced that COVID-19 vaccines will be effective) |
| 699 | 8 | 1 | 0 | 0 | c19vaccn2reas4___4 | | | Numeric | What are your reason(s) for considering not having the vaccine? Please select all that apply. (choice=Vaccines may not have been tested thoroughly in all ethnic groups) |

| | | | | | | | | | |
|---|---|---|---|---|---|---|---|---|---|
| 700 | 8 | 1 | 0 | 0 | c19vaccn2reas4___5 | | | Numeric | What are your reason(s) for considering not having the vaccine? Please select all that apply. (choice=I have had COVID-19 and therefore do not feel I need the vaccine) |
| 701 | 8 | 1 | 0 | 0 | c19vaccn2reas4___6 | | | Numeric | What are your reason(s) for considering not having the vaccine? Please select all that apply. (choice=I am taking part in a clinical trial of a COVID-19 vaccine) |
| 702 | 8 | 1 | 0 | 0 | c19vaccn2reas4___7 | | | Numeric | What are your reason(s) for considering not having the vaccine? Please select all that apply. (choice=I would prefer one of the other COVID-19 vaccines that are being developed) |
| 703 | 8 | 1 | 0 | 0 | c19vaccn2reas4___8 | | | Numeric | What are your reason(s) for considering not having the vaccine? Please select all that apply. (choice=I would prefer to wait until many other people have received a COVID-19 vaccine) |
| 704 | 8 | 1 | 0 | 0 | c19vaccn2reas4___9 | | | Numeric | What are your reason(s) for considering not having the vaccine? Please select all that apply. (choice=I do not feel that I personally am at risk from COVID-19) |
| 705 | 8 | 1 | 0 | 0 | c19vaccn2reas4___10 | | | Numeric | What are your reason(s) for considering not having the vaccine? Please select all that apply. (choice=I would rather the vaccine were used for other people who need it more than I do) |
| 706 | 8 | 1 | 0 | 0 | c19vaccn2reas4___11 | | | Numeric | What are your reason(s) for considering not having the vaccine? Please select all that apply. (choice=I do not believe in vaccinations in general) |
| 707 | 8 | 1 | 0 | 0 | c19vaccn2reas4___12 | | | Numeric | What are your reason(s) for considering not having the vaccine? Please select all that apply. (choice=Other reason) |
| 708 | 8 | 1 | 0 | 0 | c19vaccn2reas4___99 | | | Numeric | What are your reason(s) for considering not having the vaccine? Please select all that apply. (choice=Prefer not to answer) |
| 709 | 8 | 1 | 0 | 0 | c19vaccn2reas4txt | | | String | Please specify: |
| 710 | 8 | 1 | 0 | 0 | c19vaccn2reas5___1 | | | Numeric | What are your reason(s) for not having the vaccination? Please select all that apply. (choice=I have allergies, needle-phobia, am immuno-compromised, or have other clinical reasons not to be vaccinated) |
| 711 | 8 | 1 | 0 | 0 | c19vaccn2reas5___2 | | | Numeric | What are your reason(s) for not having the vaccination? Please select all that apply. (choice=I am concerned about the safety or potential side-effects of a COVID-19 vaccine) |
| 712 | 8 | 1 | 0 | 0 | c19vaccn2reas5___3 | | | Numeric | What are your reason(s) for not having the vaccination? Please select all that apply. (choice=I am not convinced that COVID-19 vaccines will be effective) |
| 713 | 8 | 1 | 0 | 0 | c19vaccn2reas5___4 | | | Numeric | What are your reason(s) for not having the vaccination? Please select all that apply. (choice=Vaccines may not have been tested thoroughly in all ethnic groups) |
| 714 | 8 | 1 | 0 | 0 | c19vaccn2reas5___5 | | | Numeric | What are your reason(s) for not having the vaccination? Please select all that apply. (choice=I have had COVID-19 and therefore do not feel I need the vaccine) |
| 715 | 8 | 1 | 0 | 0 | c19vaccn2reas5___6 | | | Numeric | What are your reason(s) for not having the vaccination? Please select all that apply. (choice=I am taking part in a clinical trial of a COVID-19 vaccine) |
| 716 | 8 | 1 | 0 | 0 | c19vaccn2reas5___7 | | | Numeric | What are your reason(s) for not having the vaccination? Please select all that apply. (choice=I would prefer one of the other COVID-19 vaccines that are being developed) |
| 717 | 8 | 1 | 0 | 0 | c19vaccn2reas5___8 | | | Numeric | What are your reason(s) for not having the vaccination? Please select all that apply. (choice=I would prefer to wait until many other people have received a COVID-19 vaccine) |
| 718 | 8 | 1 | 0 | 0 | c19vaccn2reas5___9 | | | Numeric | What are your reason(s) for not having the vaccination? Please select all that apply. (choice=I do not feel that I personally am at risk from COVID-19) |
| 719 | 8 | 1 | 0 | 0 | c19vaccn2reas5___10 | | | Numeric | What are your reason(s) for not having the vaccination? Please select all that apply. (choice=I would rather the vaccine were used for other people who need it more than I do) |
| 720 | 8 | 1 | 0 | 0 | c19vaccn2reas5___11 | | | Numeric | What are your reason(s) for not having the vaccination? Please select all that apply. (choice=I do not believe in vaccinations in general) |
| 721 | 8 | 1 | 0 | 0 | c19vaccn2reas5___12 | | | Numeric | What are your reason(s) for not having the vaccination? Please select all that apply. (choice=Other reason) |
| 722 | 8 | 1 | 0 | 0 | c19vaccn2reas5___99 | | | Numeric | What are your reason(s) for not having the vaccination? Please select all that apply. (choice=Prefer not to answer) |
| 723 | 8 | 1 | 0 | 0 | c19vaccn2reas5txt | | | String | Please specify: |
| 724 | 8 | 1 | 0 | 0 | c19vaccn3_rely | | | Numeric | I can rely on vaccines to stop serious infectious diseases |
| 725 | 8 | 1 | 0 | 0 | c19vaccn3_safe | | | Numeric | Although most vaccines appear to be safe, there may be problems that we have not yet discovered |
| 726 | 8 | 1 | 0 | 0 | c19vaccn3_gain | | | Numeric | Authorities promote vaccination for financial gain, not for peoples health |
| 727 | 8 | 1 | 0 | 0 | c19vaccn3_nat | | | Numeric | Being exposed to diseases naturally is safer for the immune system than being exposed through vaccination |
| 728 | 8 | 1 | 0 | 0 | c19vacchap | | | Numeric | When, if at all, do you think it will be possible to vaccinate most of the population against coronavirus? |
| 729 | 8 | 1 | 0 | 0 | c19effectscrisis_1 | | | Numeric | Effects on children and their education |
| 730 | 8 | 1 | 0 | 0 | c19effectscrisis_2 | | | Numeric | Effects on the economy and jobs |
| 731 | 8 | 1 | 0 | 0 | c19effectscrisis_3 | | | Numeric | Increasing deaths as a direct result of catching coronavirus |
| 732 | 8 | 1 | 0 | 0 | c19effectscrisis_4 | | | Numeric | Increased deaths due to fewer healthcare resources to identify and treat medical conditions other than coronavirus |
| 733 | 8 | 1 | 0 | 0 | c19effectscrisis_5 | | | Numeric | Increased mental health issues |
| 734 | 8 | 1 | 0 | 0 | c19vaccacc | | | Numeric | When a vaccine becomes available, would you wish to receive it? |
| 735 | 8 | 1 | 0 | 0 | c19inequality | | | Numeric | Do you think the coronavirus crisis will increase or decrease the level of inequality in the UK, compared with before the pandemic? |
| 736 | 8 | 1 | 0 | 0 | c19conspiracies_1 | | | Numeric | A person can be infected twice with coronavirus |
| 737 | 8 | 1 | 0 | 0 | c19conspiracies_2 | | | Numeric | Coronavirus is less infectious than the influenza virus |
| 738 | 8 | 1 | 0 | 0 | c19conspiracies_3 | | | Numeric | Coronavirus was created in a laboratory |
| 739 | 8 | 1 | 0 | 0 | c19conspiracies_4 | | | Numeric | Infection with coronavirus is equally likely in men and women |
| 740 | 8 | 1 | 0 | 0 | c19conspiracies_5 | | | Numeric | Mortality from coronavirus is higher in men than women |
| 741 | 8 | 1 | 0 | 0 | c19conspiracies_6 | | | Numeric | Most people in the UK have already had coronavirus without realising it |
| 742 | 8 | 1 | 0 | 0 | c19conspiracies_7 | | | Numeric | The current pandemic is part of a global effort to force everyone to be vaccinated whether they want to or not |
| 743 | 8 | 1 | 0 | 0 | c19conspiracies_8 | | | Numeric | The genetic material in a coronavirus is RNA, unlike that of humans which is DNA |
| 744 | 8 | 1 | 0 | 0 | c19conspiracies_9 | | | Numeric | The number of people reported as dying from coronavirus is being deliberately reduced or hidden by the authorities |
| 745 | 8 | 1 | 0 | 0 | c19conspiracies_10 | | | Numeric | The symptoms that most people blame on coronavirus appear to be linked to 5G network radiation |
| 746 | 8 | 1 | 0 | 0 | c19conspiracies_11 | | | Numeric | There is no hard evidence that coronavirus really exists |
| 747 | 9 | 0 | 0 | 1 | Section9_Raw_PersonalityEtc______________ | Raw | | Numeric | |
| 748 | 9 | 1 | 0 | 0 | lifebig5_rude | | | Numeric | Is sometimes rude to others |
| 749 | 9 | 1 | 0 | 0 | lifebig5_thorough | | | Numeric | Does a thorough job |
| 750 | 9 | 1 | 0 | 0 | lifebig5_talkative | | | Numeric | Is talkative |
| 751 | 9 | 1 | 0 | 0 | lifebig5_worries | | | Numeric | Worries a lot |
| 752 | 9 | 1 | 0 | 0 | lifebig5_orig | | | Numeric | Is original, comes up with new ideas |
| 753 | 9 | 1 | 0 | 0 | lifebig5_forgiving | | | Numeric | Has a forgiving nature |
| 754 | 9 | 1 | 0 | 0 | lifebig5_lazy | | | Numeric | Tends to be lazy |
| 755 | 9 | 1 | 0 | 0 | lifebig5_outgoing | | | Numeric | Is outgoing, sociable |
| 756 | 9 | 1 | 0 | 0 | lifebig5_nervous | | | Numeric | Gets nervous easily |
| 757 | 9 | 1 | 0 | 0 | lifebig5_art | | | Numeric | Values artistic, aesthetic experiences |
| 758 | 9 | 1 | 0 | 0 | lifebig5_kind | | | Numeric | Is considerate and kind to almost everyone |
| 759 | 9 | 1 | 0 | 0 | lifebig5_eff | | | Numeric | Does things efficiently |
| 760 | 9 | 1 | 0 | 0 | lifebig5_res | | | Numeric | Is reserved |
| 761 | 9 | 1 | 0 | 0 | lifebig5_stress | | | Numeric | Is relaxed, handles stress well |
| 762 | 9 | 1 | 0 | 0 | lifebig5_imag | | | Numeric | Has an active imagination |
| 763 | 9 | 1 | 0 | 0 | lifeloc_1 | | | Numeric | My life is determined by my own actions |

| | | | | | | | | | | | | | |
|---|---|---|---|---|---|---|---|---|---|---|---|---|---|
| 764 | 9 | 1 | 0 | 0 | lifeloc_2 | | | | | | | Numeric | I am usually able to protect my personal interests |
| 765 | 9 | 1 | 0 | 0 | lifeloc_3 | | | | | | | Numeric | I can pretty much determine what will happen in my life |
| 766 | 9 | 1 | 0 | 0 | lifeloc_4 | | | | | | | Numeric | To a great extent, my life is controlled by accidental happenings |
| 767 | 9 | 1 | 0 | 0 | lifeloc_5 | | | | | | | Numeric | Often there is no chance of protecting my personal interest from bad luck happenings |
| 768 | 9 | 1 | 0 | 0 | lifeloc_6 | | | | | | | Numeric | When I get what I want, its usually because Im lucky |
| 769 | 9 | 1 | 0 | 0 | lifeloc_7 | | | | | | | Numeric | People like myself have very little chance of protecting our personal interests where they conflict with those of strong pressure groups |
| 770 | 9 | 1 | 0 | 0 | lifeloc_8 | | | | | | | Numeric | My life is chiefly controlled by powerful others |
| 771 | 9 | 1 | 0 | 0 | lifeloc_9 | | | | | | | Numeric | I feel like what happens in my life is mostly determined by powerful people |
| 772 | 9 | 1 | 0 | 0 | lifefatlh_1 | | | | | | | Numeric | If someone is meant to have a serious disease, they will get that disease. |
| 773 | 9 | 1 | 0 | 0 | lifefatlh_2 | | | | | | | Numeric | My health is determined by fate. |
| 774 | 9 | 1 | 0 | 0 | lifefatlh_3 | | | | | | | Numeric | My health is determined by something greater than myself. |
| 775 | 9 | 1 | 0 | 0 | lifefatlh_4 | | | | | | | Numeric | I will stay healthy if I am lucky. |
| 776 | 9 | 1 | 0 | 0 | lifenfcc_1 | | | | | | | Numeric | I like having a clear and structured mode of life |
| 777 | 9 | 1 | 0 | 0 | lifenfcc_2 | | | | | | | Numeric | I would quickly become impatient and irritated if I could not find a solution to a problem immediately |
| 778 | 9 | 1 | 0 | 0 | lifefatlh_5 | | | | | | | Numeric | Genes are more important than ones own behaviour in determining ones health |
| 779 | 9 | 1 | 0 | 0 | lifegnrisk | | | | | | | Numeric | How do you see yourself: are you generally a person who is fully prepared to take risks or do you try to avoid taking risks? Please select a number on the scale, where the value 0 means: not at all willing to take risks and the value 10 means: very willing |
| 780 | 9 | 1 | 0 | 0 | pastworkwcqf_1 | | | | | | | Numeric | There was a real opportunity for me to choose the particular things I worked on |
| 781 | 9 | 1 | 0 | 0 | pastworkwcqf_2 | | | | | | | Numeric | My work colleagues really tried hard to get to know one another |
| 782 | 9 | 1 | 0 | 0 | pastworkwcqf_3 | | | | | | | Numeric | I had a lot of choice about the work I did |
| 783 | 9 | 1 | 0 | 0 | pastworkwcqf_4 | | | | | | | Numeric | I was required to do too many different things |
| 784 | 9 | 1 | 0 | 0 | pastworkwcqf_5 | | | | | | | Numeric | My coworkers were supportive and friendly towards me |
| 785 | 9 | 1 | 0 | 0 | pastworkwcqf_6 | | | | | | | Numeric | There seemed to be too much work to get through |
| 786 | 9 | 1 | 0 | 0 | workengbout_1 | | | | | | | Numeric | I was exhausted in the morning at the thought of another day at work |
| 787 | 9 | 1 | 0 | 0 | workengbout_2 | | | | | | | Numeric | When I was working I forgot everything else around me |
| 788 | 9 | 1 | 0 | 0 | workengbout_3 | | | | | | | Numeric | I didnt have enough energy for family and friends during leisure time |
| 789 | 9 | 1 | 0 | 0 | workengbout_4 | | | | | | | Numeric | I was proud of the work that I did |
| 790 | 9 | 1 | 0 | 0 | workengbout_5 | | | | | | | Numeric | At work I felt bursting with energy |
| 791 | 10 | 0 | 0 | 1 | Section10_Raw_FinalQuestions_____________Raw | | | | | | | Numeric | |
| 792 | 10 | 1 | 0 | 0 | exbamec19 | | | | | | | String | What are your thoughts on why people from ethnic minorities working in health and careÂ have been more severely affected by COVID-19? |
| 793 | 10 | 1 | 0 | 0 | exsocc19 | | | | | | | String | How do you see society changing as a result of COVID-19? |
| 794 | 10 | 1 | 0 | 0 | exyouc19 | | | | | | | String | How do you see your own future changing as a result of COVID-19? |
| 795 | 10 | 1 | 0 | 0 | exlength | | | | | | | Numeric | We know that the questionnaire was long. Did you think: |
| 796 | 10 | 1 | 0 | 0 | exutility | | | | | | | Numeric | How useful do you think this questionnaire might be for researching and understanding COVID-19 in ethnic minorities? |
| 797 | 10 | 0 | 0 | 1 | End_of_Raw_Questions_____________Raw | | | | | | | Numeric | |
| 798 | 0 | 0 | 0 | 1 | Section0_RegistrationEtc_____________Derived | | | | | | | Numeric | |
| 799 | 0 | 0 | 1 | 0 | ID2 | | | | | | | Numeric | |
| 800 | 0 | 0 | 1 | 0 | Questionnaire_Completed | | | | | | | Numeric | |
| 801 | 0 | 0 | 1 | 0 | OddEven | | | | | | | Numeric | ID split on Odd-Even to assess replication of results |
| 801.5 | 0 | 0 | 1 | 0 | age | 1 | 1 | 2 | Older | 2 | 14 | Numeric | Age in years on date of registration [NB differs from 2021-01-19 version which used date of download] |
| 802 | 0 | 0 | 1 | 0 | age10_5to4 | | | | | | | Numeric | |
| 803 | 0 | 0 | 1 | 0 | Age10_5to4_5decades | | | | | | | Numeric | Age in decades <= 34 to >= 65 |
| 804 | 0 | 0 | 1 | 0 | age10_0to9 | | | | | | | Numeric | |
| 805 | 0 | 0 | 1 | 0 | Age_ge_45 | | | | | | | Numeric | |
| 806 | 0 | 0 | 1 | 0 | AgeDecades | | | | | | | Numeric | |
| 807 | 0 | 0 | 1 | 0 | Regulator_2 | 1 | | | | | | Numeric | Regulator: recoded for clarity |
| 808 | 0 | 0 | 1 | 0 | Index.of.Multiple.Deprivation.quintile | | | | | | | Numeric | |
| 809 | 0 | 0 | 1 | 0 | Income.quintile | | | | | | | Numeric | |
| 810 | 0 | 0 | 1 | 0 | Employment.quintile | | | | | | | Numeric | |
| 811 | 0 | 0 | 1 | 0 | Education.and.Skills.quintile | | | | | | | Numeric | |
| 812 | 0 | 0 | 1 | 0 | Health.and.Disability.quintile | | | | | | | Numeric | |
| 813 | 0 | 0 | 1 | 0 | Crime.quintile | | | | | | | Numeric | |
| 814 | 0 | 0 | 1 | 0 | Barriers.to.Housing.and.Services.quintile | | | | | | | Numeric | |
| 815 | 0 | 0 | 1 | 0 | Living.Environment.quintile | | | | | | | Numeric | |
| 816 | 0 | 0 | 1 | 0 | IDACI.quintile | | | | | | | Numeric | |
| 817 | 0 | 0 | 1 | 0 | IDAOPI.quintile | | | | | | | Numeric | |
| 818 | 0 | 0 | 1 | 0 | DayOfConsent | 1 | 1 | 3 | Later consen | 3 | 92 | Numeric | From Redcap date.of.consent but probably DayOfStart |
| 819 | 0 | 0 | 1 | 0 | DayOfCompletion | 1 | 1 | 3 | Later comple | 3 | 91 | Numeric | From Redcap date_quest_complete |
| 820 | 0 | 0 | 1 | 0 | DaysConsentToCompletion | | | | | | | Numeric | Days from Consent to Completion |
| 821 | 0 | 0 | 1 | 0 | DayOfConsent_flag | | | | | | | Numeric | Date of Consent present |
| 822 | 0 | 0 | 1 | 0 | DayOfCompletion_flag | | | | | | | Numeric | Date of Completion present |
| 823 | 0 | 0 | 1 | 0 | DayOfCompletion_2 | | | | | | | Numeric | From Redcap date_quest_complete |
| 824 | 0 | 0 | 1 | 0 | DaysConsentToCompletion_2 | | | | | | | Numeric | Days from Consent to Completion |
| 825 | 1 | 0 | 0 | 1 | Section1_DemographicsEtc_____________Derived | | | | | | | Numeric | |
| 826 | 1 | 0 | 1 | 0 | Woman | 1 | 1 | 4 | Female | 4 | 254 | Numeric | Self-describes as Woman (rather than Man) |

| | | | | | | | | | | | | | |
|---|---|---|---|---|---|---|---|---|---|---|---|---|---|
| 827 | 1 | 0 | 1 | 0 | FemaleSexAtBirth | | | | | | | Numeric | |
| 828 | 1 | 0 | 1 | 0 | Single | | | | | | | Numeric | Single -- not in or has been in a relationship |
| 829 | 2 | 0 | 0 | 1 | Section2_Job______________Derived | | | | | | | Numeric | |
| 830 | 2 | 0 | 1 | 0 | jobcode | 4 | | | | | | Numeric | job from jobrole |
| 831 | 2 | 0 | 1 | 0 | Jobcode2 | | | | | | | Numeric | Job categories -- n=5 |
| 832 | 2 | 0 | 1 | 0 | Job_doctor | | 1 | 3 | Doctor | 3 | 183 | Numeric | |
| 833 | 2 | 0 | 1 | 0 | Job_nurse | | 1 | | | 3 | 184 | Numeric | |
| 834 | 2 | 0 | 1 | 0 | Job_AHPsEtc | | 1 | | | 7 | 181 | Numeric | |
| 835 | 2 | 0 | 1 | 0 | Job_dental | | 1 | 3 | Dental | 7 | 182 | Numeric | |
| 836 | 2 | 0 | 1 | 0 | Job_AdminEtc | | 1 | | | 7 | 180 | Numeric | |
| 837 | 2 | 0 | 1 | 0 | Working_Now | | | | | | | Numeric | Working when Quest completed |
| 838 | 2 | 0 | 1 | 0 | Working_Lockdown1 | | | | | | | Numeric | Working when in Lockdown 1 |
| 839 | 2 | 0 | 1 | 0 | Sector_Now_NHS | 1 | 1 | 3 | Employed in | 3 | 232 | Numeric | Working in NHSnow (or private used by NHS) |
| 840 | 2 | 0 | 1 | 0 | Sector_Lockown_NHS | | | | | | | Numeric | |
| 841 | 2 | 0 | 1 | 0 | Sector_Lockdown_NHS | 1 | 1 | | | 7 | 231 | Numeric | Working in NHS lockdown (or private used by NHS) |
| 842 | 2 | 0 | 1 | 0 | PostCodeArea | | | | | | | String | |
| 843 | 2 | 0 | 1 | 0 | UKregion_work | 4 | | | | | | String | |
| 844 | 2 | 0 | 1 | 0 | UKregion_EastOfEngland | | 1 | | | 7 | 240 | Numeric | |
| 845 | 2 | 0 | 1 | 0 | UKregion_EastMidlands | | 1 | | | 7 | 239 | Numeric | |
| 846 | 2 | 0 | 1 | 0 | UKregion_OuterLondon | | 1 | | | 7 | 245 | Numeric | |
| 847 | 2 | 0 | 1 | 0 | UKregion_InnerLondon | | 1 | | | 7 | 241 | Numeric | |
| 848 | 2 | 0 | 1 | 0 | UKregion_NorthEast | | 1 | | | 7 | 242 | Numeric | |
| 849 | 2 | 0 | 1 | 0 | UKregion_NorthWestIncIoM | | 1 | | | 7 | 244 | Numeric | |
| 850 | 2 | 0 | 1 | 0 | UKregion_NorthernIreland | | 1 | | | 7 | 243 | Numeric | |
| 851 | 2 | 0 | 1 | 0 | UKregion_Scotland | | 1 | | | 7 | 246 | Numeric | |
| 852 | 2 | 0 | 1 | 0 | UKregion_SouthEast | | 1 | | | 7 | 247 | Numeric | |
| 853 | 2 | 0 | 1 | 0 | UKregion_SouthWestIncCI | | 1 | | | 7 | 248 | Numeric | |
| 854 | 2 | 0 | 1 | 0 | UKregion_SouthWest | | | | | | | Numeric | |
| 855 | 2 | 0 | 1 | 0 | UKregion_WestMidlands | | 1 | | | 7 | 249 | Numeric | |
| 856 | 2 | 0 | 1 | 0 | JobBand_Now_Quartiles | | | | | | | Numeric | |
| 857 | 2 | 0 | 1 | 0 | JobBandIncDr_Now_Quartiles | | | | | | | Numeric | |
| 858 | 2 | 0 | 1 | 0 | JobBand_Lockdown_Quartiles | | | | | | | Numeric | |
| 859 | 2 | 0 | 1 | 0 | JobBandIncDr_Lockdown_Quartiles | | | | | | | Numeric | |
| 860 | 2 | 0 | 1 | 0 | job_areas_answered_all | | | | | | | Numeric | |
| 861 | 2 | 0 | 1 | 0 | Practice_Hospital | 1 | 1 | | | 7 | 215 | Numeric | Any hospital location mentioned [areas 5 to 14 + 17] |
| 862 | 2 | 0 | 1 | 0 | Practice_General_Practice | 1 | 1 | | | 7 | 214 | Numeric | Community clinical/primary setting [code 3] |
| 863 | 2 | 0 | 1 | 0 | Practice_Community | 1 | 1 | | | 7 | 213 | Numeric | Ambulance; Armed Forces; Comm non-clin; Nursing/care home; Prison |
| 864 | 2 | 0 | 1 | 0 | Practice_Other | | | | | | | Numeric | University/home/other |
| 865 | 2 | 0 | 1 | 0 | HoursWork_Now | 1 | 1 | | | 7 | 175 | Numeric | |
| 866 | 2 | 0 | 1 | 0 | HoursWork_Lockdown | 1 | 1 | | | 7 | 174 | Numeric | |
| 867 | 2 | 0 | 1 | 0 | NightWork_Now | 1 | 1 | 5 | Night work c | 5 | 208 | Numeric | |
| 868 | 2 | 0 | 1 | 0 | NightWork_Lockdown | 1 | 1 | | | 7 | 207 | Numeric | |
| 869 | 2 | 0 | 1 | 0 | Covid_Exposure | 1 | 1 | 4 | Covid-19 Exp | 4 | 70 | Numeric | |
| 870 | 2 | 0 | 1 | 0 | Covid_Exposure_past | 1 | 1 | | | 7 | 71 | Numeric | |
| 872 | 2 | 0 | 1 | 0 | Covid_Exp_mean_past | | | | | | | Numeric | |
| 873 | 2 | 0 | 1 | 0 | Covid_ExposureFtFsocdisPtsNow | | | | | | | Numeric | |
| 874 | 2 | 0 | 1 | 0 | Covid_ExposurePhysContPtsNow | | | | | | | Numeric | |
| 875 | 2 | 0 | 1 | 0 | Covid_Exposure_Pts_Now | | | | | | | Numeric | Covid exposure from patients now |
| 876 | 2 | 0 | 1 | 0 | Covid_ExposureFtFsocdisPtsLD | | | | | | | Numeric | |
| 877 | 2 | 0 | 1 | 0 | Covid_ExposurePhysContPtsLD | | | | | | | Numeric | |
| 878 | 2 | 0 | 1 | 0 | Covid_Exposure_Pts_LD | | | | | | | Numeric | Covid exposure from patients in first lockdown |
| 879 | 2 | 0 | 1 | 0 | Covid_Exposure_Pts_NowBIN | | | | | | | Numeric | |
| 880 | 2 | 0 | 1 | 0 | Covid_Exposure_Pts_LDBIN | | | | | | | Numeric | |
| 881 | 2 | 0 | 1 | 0 | TravelTime_Now | 1 | 1 | | | 7 | 237 | Numeric | |
| 882 | 2 | 0 | 1 | 0 | TravelTime_Lockdown | 1 | 1 | | | 7 | 236 | Numeric | |
| 883 | 2 | 0 | 1 | 0 | PPEaccess_now | 1 | 1 | | | 7 | 212 | Numeric | |
| 884 | 2 | 0 | 1 | 0 | PPEaccess_ld | 1 | 1 | | | 5 | 211 | Numeric | |
| 885 | 2 | 0 | 1 | 0 | AerosolGPs_Now | 1 | 1 | | | 7 | 13 | Numeric | |
| 886 | 2 | 0 | 1 | 0 | AerosolGPs_Lockdown | 1 | 1 | | | 7 | 12 | Numeric | |
| 887 | 2 | 0 | 1 | 0 | RiskAssessmentDone | 1 | 1 | | | 7 | 229 | Numeric | |
| 888 | 2 | 0 | 1 | 0 | RiskAssessmentChangesMade | | | | | | | Numeric | |
| 889 | 2 | 0 | 1 | 0 | Concerns_FeelSecureRaising | 1 | 1 | | | 7 | 59 | Numeric | |
| 890 | 2 | 0 | 1 | 0 | Concerns_ConfidentAddressed | 1 | 1 | | | 7 | 58 | Numeric | |
| 891 | 2 | 0 | 1 | 0 | Redeployed_RoleChange | 1 | 1 | | | 5 | 218 | Numeric | |

| | | | | | | | | | | | | | |
|---|---|---|---|---|---|---|---|---|---|---|---|---|---|
| 892 | 2 | 0 | 1 | 0 | Redeployed_MorePtContact | | | | | | | Numeric | |
| 893 | 3 | 0 | 0 | 1 | Section3_EthnicityEtc_____________Derived | | | | | | | Numeric | |
| 894 | 3 | 0 | 1 | 0 | Ethnic18 | 4 | | | | | | Numeric | All 18 ONS ethnic categories |
| 895 | 3 | 0 | 1 | 0 | Ethnic18_Indian | | 1 | | | 7 | 106 | Numeric | |
| 896 | 3 | 0 | 1 | 0 | Ethnic18_Pakistani | | 1 | 5 | ONS18: Pakis | 5 | 116 | Numeric | |
| 897 | 3 | 0 | 1 | 0 | Ethnic18_Bangladeshi | | 1 | | | 7 | 102 | Numeric | |
| 898 | 3 | 0 | 1 | 0 | Ethnic18_Chinese | | 1 | 5 | ONS18: Chin | 7 | 105 | Numeric | |
| 899 | 3 | 0 | 1 | 0 | Ethnic18_OtherAsian | | 1 | | | 7 | 113 | Numeric | |
| 900 | 3 | 0 | 1 | 0 | Ethnic18_African | | 1 | 6 | ONS18: Afric | 6 | 100 | Numeric | |
| 901 | 3 | 0 | 1 | 0 | Ethnic18_Caribbean | | 1 | | | 7 | 104 | Numeric | |
| 902 | 3 | 0 | 1 | 0 | Ethnic18_OtherBlack | | 1 | | | 7 | 114 | Numeric | |
| 903 | 3 | 0 | 1 | 0 | Ethnic18_Mixed_White_Caribbean | | 1 | | | 7 | 111 | Numeric | |
| 904 | 3 | 0 | 1 | 0 | Ethnic18_Mixed_White_African | | 1 | | | 7 | 109 | Numeric | |
| 905 | 3 | 0 | 1 | 0 | Ethnic18_Mixed_White_Asian | | 1 | | | 7 | 110 | Numeric | |
| 906 | 3 | 0 | 1 | 0 | Ethnic18_Mixed_Other | | 1 | | | 7 | 108 | Numeric | |
| 907 | 3 | 0 | 1 | 0 | Ethnic18_British | | 1 | | | 7 | 103 | Numeric | |
| 908 | 3 | 0 | 1 | 0 | Ethnic18_Irish | | 1 | | | 7 | 107 | Numeric | |
| 909 | 3 | 0 | 1 | 0 | Ethnic18_OtherIncTraveller | | 1 | | | 7 | 115 | Numeric | |
| 910 | 3 | 0 | 1 | 0 | Ethnic18_Arab | | 1 | | | 7 | 101 | Numeric | |
| 911 | 3 | 0 | 1 | 0 | Ethnic18_Other_Other | | 1 | | | 7 | 112 | Numeric | |
| 912 | 3 | 0 | 1 | 0 | ethnic5 | | | | | | | Numeric | Ethnicity: Five categories |
| 913 | 3 | 0 | 1 | 0 | BAME | | | | | | | Numeric | Black and Minority Ethnic |
| 914 | 3 | 0 | 1 | 0 | Ethnic5_White | | 1 | 4 | ONS5: White | 4 | 121 | Numeric | |
| 915 | 3 | 0 | 1 | 0 | Ethnic5_Asian_Chinese | | 1 | | | 5 | 117 | Numeric | |
| 916 | 3 | 0 | 1 | 0 | Ethnic5_Black | | 1 | | | 7 | 118 | Numeric | |
| 917 | 3 | 0 | 1 | 0 | Ethnic5_Other | | 1 | | | 7 | 120 | Numeric | |
| 918 | 3 | 0 | 1 | 0 | Ethnic5_Mixed | | 1 | | | 7 | 119 | Numeric | |
| 918.5 | 3 | 0 | 1 | 0 | Ethnic18_Mother | 4 | | | | | | | Ethnicity Mother - 18 categories |
| 919 | 3 | 0 | 1 | 0 | Ethnic5_Mother | | | | | | | Numeric | Ethnicity Mother: Five categories |
| 920 | 3 | 0 | 1 | 0 | BAME_Mother | | | | | | | Numeric | |
| 920.5 | 3 | 0 | 1 | 0 | Ethnic18_Father | 4 | | | | | | | Ethnicity Father - 18 categories |
| 921 | 3 | 0 | 1 | 0 | Ethnic5_Father | | | | | | | Numeric | Ethnicity Father: Five categories |
| 922 | 3 | 0 | 1 | 0 | BAME_Father | | | | | | | Numeric | |
| 922.5 | 3 | 0 | 1 | 0 | Ethnic18_Partner | 4 | | | | | | | Ethnicity artner - 18 categories |
| 923 | 3 | 0 | 1 | 0 | ethnic5_Partner | | | | | | | Numeric | Ethnicity Partner: Five categories |
| 924 | 3 | 0 | 1 | 0 | BAME_partner | | | | | | | Numeric | |
| 925 | 3 | 0 | 1 | 0 | UKnational | 1 | 1 | | | 7 | 238 | Numeric | UK national from UK_nat_1 to 3 |
| 926 | 3 | 0 | 1 | 0 | CountryOfTrainingUK | 1 | 1 | 4 | UK Training | 4 | 60 | Numeric | Primary qualification in UK; from langedpmq |
| 927 | 3 | 0 | 1 | 0 | CountryOfTrainingUK_2 | | | | | | | Numeric | Primary qualification in UK; from langedpmq inc no PMQ |
| 928 | 3 | 0 | 1 | 0 | BornUK | 1 | 1 | | | 7 | 26 | Numeric | Born in the UK - from ethbornUK |
| 929 | 3 | 0 | 1 | 0 | BornUKparents | | | | | | | Numeric | Number of parents born in UK |
| 930 | 3 | 0 | 1 | 0 | BornUKparents_oneplus | 1 | 1 | | | 7 | 28 | Numeric | |
| 931 | 3 | 0 | 1 | 0 | BornUKgrandparents | | | | | | | Numeric | Number of grandparents born in UK |
| 932 | 3 | 0 | 1 | 0 | BornUKgrandparents_oneplus | 1 | 1 | | | 7 | 27 | Numeric | |
| 933 | 3 | 0 | 1 | 0 | EnglishSpokenLate | | | | | | | Numeric | Age English spoken 1=Early 5 = Late |
| 934 | 3 | 0 | 1 | 0 | EnglishSpoken_6plus | 1 | 1 | | | 7 | 98 | Numeric | |
| 935 | 3 | 0 | 1 | 0 | Enghome1 | | | | | | | Numeric | |
| 936 | 3 | 0 | 1 | 0 | Enghome2 | | | | | | | Numeric | |
| 937 | 3 | 0 | 1 | 0 | LanguageAtHome | | | | | | | Numeric | |
| 938 | 3 | 0 | 1 | 0 | EnglishSpokenAtHome | 1 | 1 | | | 7 | 99 | Numeric | English spoken at home |
| 939 | 3 | 0 | 1 | 0 | EngAtHomeAsChild | 1 | 1 | | | 7 | 97 | Numeric | English at home as a child: langedch___11 = 1 |
| 940 | 3 | 0 | 1 | 0 | LangedChildSum | | | | | | | Numeric | Number of languages spoken as a child |
| 941 | 3 | 0 | 1 | 0 | BilingualAsChild | 1 | 1 | | | 7 | 23 | Numeric | 2+ languages as a child |
| 942 | 3 | 0 | 1 | 0 | HigherEducationLevel | 1 | 1 | 3 | Higher Educa | 3 | 170 | Numeric | |
| 943 | 3 | 0 | 1 | 0 | HigherEducationLevel_Mother | 1 | 1 | | | 7 | 172 | Numeric | |
| 944 | 3 | 0 | 1 | 0 | HigherEducationLevel_Father | 1 | 1 | | | 7 | 171 | Numeric | |
| 945 | 3 | 0 | 1 | 0 | religion | 4 | | | | | | Numeric | Religion from Census 2011 -- raw variable relig |
| 946 | 3 | 0 | 1 | 0 | relig_None | | 1 | | | 7 | 224 | Numeric | |
| 947 | 3 | 0 | 1 | 0 | relig_Christian | | 1 | | | 7 | 220 | Numeric | |
| 948 | 3 | 0 | 1 | 0 | relig_Buddhist | | 1 | | | 7 | 219 | Numeric | |
| 949 | 3 | 0 | 1 | 0 | relig_Hindu | | 1 | | | 7 | 221 | Numeric | |
| 950 | 3 | 0 | 1 | 0 | relig_Jewish | | 1 | | | 7 | 222 | Numeric | |
| 951 | 3 | 0 | 1 | 0 | relig_Muslim | | 1 | 4 | ONS: Muslim | 4 | 223 | Numeric | |
| 952 | 3 | 0 | 1 | 0 | relig_Sikh | | 1 | | | 7 | 226 | Numeric | |

| | | | | | | | | | | | | | |
|---|---|---|---|---|---|---|---|---|---|---|---|---|---|
| 953 | 3 | 0 | 1 | 0 | relig_Other | | 1 | | | 7 | 225 | Numeric | |
| 954 | 3 | 0 | 1 | 0 | religious_bin | | | | | | | Numeric | Responded as any religion vs no religion |
| 955 | 3 | 0 | 1 | 0 | ReligionImportantEveryday_Raw | | | | | | | Numeric | |
| 956 | 3 | 0 | 1 | 0 | ReligionImportantEveryday | 1 | 1 | 4 | Religion Impo | 4 | 227 | Numeric | |
| 957 | 3 | 0 | 1 | 0 | ReligionImportantUpbringing | 1 | 1 | | | 7 | 228 | Numeric | From relimp2 |
| 958 | 3 | 0 | 1 | 0 | HolyPlaceOutsideHome_Raw | | | | | | | Numeric | |
| 959 | 3 | 0 | 1 | 0 | HolyPlaceOutsideHome | 1 | 1 | | | 7 | 173 | Numeric | |
| 960 | 3 | 0 | 1 | 0 | CultureImpToIdentity | 1 | 1 | | | 7 | 90 | Numeric | Culture important to identity 0 Low 10 High; from ethimp |
| 961 | 3 | 0 | 1 | 0 | CultureImpOfMother | 1 | 1 | | | 7 | 88 | Numeric | Culture important to identity 0 Low 10 High; from ethimpm |
| 962 | 3 | 0 | 1 | 0 | CultureImpOfFather | 1 | 1 | | | 7 | 87 | Numeric | Culture important to identity 0 Low 10 High; from ethimpf |
| 963 | 3 | 0 | 1 | 0 | CultureImpOfPartner | 1 | 1 | | | 7 | 89 | Numeric | Culture important to identity 0 Low 10 High; from ethimpf |
| 964 | 4 | 0 | 0 | 1 | Section4_HomeAndFamilyLife____________ | Derived | | | | | | Numeric | |
| 965 | 4 | 0 | 1 | 0 | SupportBubble | 1 | 1 | | | 7 | 234 | Numeric | |
| 966 | 4 | 0 | 1 | 0 | ChildSupportBubble | 1 | 1 | | | 7 | 52 | Numeric | |
| 967 | 4 | 0 | 1 | 0 | TotalHouseholdSize | 1 | 1 | | | 7 | 235 | Numeric | Includes respondent: hf_nhhold+1 |
| 968 | 4 | 0 | 1 | 0 | TempAgeRespondent | | | | | | | Numeric | |
| 969 | 4 | 0 | 1 | 0 | N_Household_0to1 | 1 | 1 | | | 7 | 190 | Numeric | |
| 970 | 4 | 0 | 1 | 0 | N_Household_2to4 | 1 | 1 | | | 7 | 195 | Numeric | |
| 971 | 4 | 0 | 1 | 0 | N_Household_5to10 | 1 | 1 | | | 7 | 199 | Numeric | |
| 972 | 4 | 0 | 1 | 0 | N_Household_11to16 | 1 | 1 | | | 7 | 191 | Numeric | |
| 973 | 4 | 0 | 1 | 0 | N_Household_17to18 | 1 | 1 | | | 7 | 192 | Numeric | |
| 974 | 4 | 0 | 1 | 0 | N_Household_19to24 | 1 | 1 | | | 7 | 193 | Numeric | |
| 975 | 4 | 0 | 1 | 0 | N_Household_25to34 | 1 | 1 | | | 7 | 194 | Numeric | |
| 976 | 4 | 0 | 1 | 0 | N_Household_35to44 | 1 | 1 | | | 7 | 196 | Numeric | |
| 977 | 4 | 0 | 1 | 0 | N_Household_45to54 | 1 | 1 | | | 7 | 197 | Numeric | |
| 978 | 4 | 0 | 1 | 0 | N_Household_55to64 | 1 | 1 | | | 7 | 198 | Numeric | |
| 979 | 4 | 0 | 1 | 0 | N_Household_65to74 | 1 | 1 | | | 7 | 200 | Numeric | |
| 980 | 4 | 0 | 1 | 0 | N_Household_75to84 | 1 | 1 | | | 7 | 201 | Numeric | |
| 981 | 4 | 0 | 1 | 0 | N_Household_85to94 | 1 | 1 | | | 7 | 202 | Numeric | |
| 982 | 4 | 0 | 1 | 0 | N_Household_95plus | 1 | 1 | | | 7 | 203 | Numeric | |
| 983 | 4 | 0 | 1 | 0 | N_Household_UsePublicTransport | | | | | | | Numeric | Household: Public Transport from hfriskjob |
| 984 | 4 | 0 | 1 | 0 | N_Household_JobsCloseContact | | | | | | | Numeric | Household: Close Phys Contact from hfriskjob_2 |
| 985 | 4 | 0 | 1 | 0 | N_Household_PubTransBIN | 1 | 1 | | | 7 | 205 | Numeric | Household: Pub Trans BIN |
| 986 | 4 | 0 | 1 | 0 | N_Household_JobsCloseContactBIN | 1 | 1 | | | 7 | 204 | Numeric | Household: Close Phys Contact BIN |
| 987 | 4 | 0 | 1 | 0 | Address_YearMoved | | | | | | | Numeric | |
| 988 | 4 | 0 | 1 | 0 | Address_Year_Moved | | | | | | | Numeric | year moved to address from hfcuradd |
| 989 | 4 | 0 | 1 | 0 | Accomn_Type | 4 | | | | | | Numeric | Accomn type -- reduced from hfacctype |
| 989.1 | 4 | 0 | 1 | 0 | Accomn_Detached | | 1 | | | 7 | 2 | | |
| 989.2 | 4 | 0 | 1 | 0 | Accomn_SemiDetached | | 1 | | | 7 | 10 | | |
| 989.3 | 4 | 0 | 1 | 0 | Accomn_Terraced | | 1 | | | 7 | 11 | | |
| 989.4 | 4 | 0 | 1 | 0 | Accomn_Flat | | 1 | | | 7 | 4 | | |
| 989.5 | 4 | 0 | 1 | 0 | Accomn_Other | | 1 | | | 7 | 7 | | |
| 990 | 4 | 0 | 1 | 0 | Accomn_EmployerProvided | 1 | 1 | | | 7 | 3 | Numeric | |
| 991 | 4 | 0 | 1 | 0 | Accomn_Nrooms | 1 | 1 | | | 7 | 6 | Numeric | N rooms: from hfaccrooms; > 16 made missing e.g. 103 |
| 992 | 4 | 0 | 1 | 0 | Accomn_PeoplePerRoom | 1 | 1 | | | 7 | 8 | Numeric | TotalHousehold/Rooms -- >2 set at 2 |
| 993 | 4 | 0 | 1 | 0 | Accomn_RoomsShared | 1 | 1 | | | 7 | 9 | Numeric | Shared Rooms: 1=yes, 0 = no, from hfaccshbath___4 |
| 994 | 4 | 0 | 1 | 0 | Accomn_CommunalAreas | 1 | 1 | | | 7 | 1 | Numeric | Comumnal Areas: from hf_share |
| 995 | 4 | 0 | 1 | 0 | Accomn_Garden | 1 | 1 | | | 7 | 5 | Numeric | Garden: from hfgarden |
| 996 | 5 | 0 | 0 | 1 | Section5_FriendsAndSocialNetwork____________ | Derived | | | | | | Numeric | |
| 997 | 5 | 0 | 1 | 0 | Commn_Remotely | 1 | 1 | | | 7 | 55 | Numeric | Comm Remotely: from socncommnowrem |
| 998 | 5 | 0 | 1 | 0 | Commn_FaceToFace | 1 | 1 | | | 7 | 53 | Numeric | Comm FaceToFace: from socncommnowftfx |
| 999 | 5 | 0 | 1 | 0 | Commn_PhysicalContact | 1 | 1 | | | 7 | 54 | Numeric | Comm Physical Contact: from socncommnowpcx |
| 1000 | 5 | 0 | 1 | 0 | FriendsSameEthnicity | 1 | 1 | | | 7 | 127 | Numeric | Friends of same ethnicity as oneself: from soceth2 |
| 1001 | 6 | 0 | 0 | 1 | Section6_HarassmentAndDiscn____________ | Derived | | | | | | Numeric | |
| 1002 | 6 | 0 | 1 | 0 | Harassment_LowRespect | 1 | 1 | | | 7 | 130 | Numeric | Harassment Low Respect -- items harasseds_1,2,3,4,7; low=1 High=6 |
| 1003 | 6 | 0 | 1 | 0 | Harassment_FearLowTrust | 1 | 1 | | | 7 | 128 | Numeric | Harassment Fear Low Trust -- items harasseds_5,6; low=1 High=6 |
| 1004 | 6 | 0 | 1 | 0 | Harassment_InsultsThreats | 1 | 1 | | | 7 | 129 | Numeric | Harassment Insults Threats -- items harasseds_8,9; low=1 High=6 |
| 1005 | 6 | 0 | 1 | 0 | DiscriminationAtWork | 1 | 1 | | | 7 | 94 | Numeric | |
| 1006 | 6 | 0 | 1 | 0 | DiscAtWork_EthNatRel | 1 | 1 | | | 7 | 93 | Numeric | |
| 1007 | 7 | 0 | 0 | 1 | Section7_Health______________Derived | | | | | | | Numeric | |
| 1008 | 7 | 0 | 1 | 0 | HeightCms | | | | | | | Numeric | |
| 1009 | 7 | 0 | 1 | 0 | WeightKgms | | | | | | | Numeric | |
| 1010 | 7 | 0 | 1 | 0 | HeightInches | | | | | | | Numeric | |
| 1011 | 7 | 0 | 1 | 0 | WeightPounds | | | | | | | Numeric | |

| | | | | | | | | | | | | | |
|---|---|---|---|---|---|---|---|---|---|---|---|---|---|
| 1012 | 7 | 0 | 1 | 0 | **BMI** | 1 | 1 | | | 7 | 24 | Numeric | |
| 1013 | 7 | 0 | 1 | 0 | **BMI_group** | | 1 | | | 7 | 25 | Numeric | |
| 1014 | 7 | 0 | 1 | 0 | **SmokeEver** | 1 | 1 | | | 7 | 233 | Numeric | Current or Ex-Smoker: from hlthsmk |
| 1015 | 7 | 0 | 1 | 0 | **Vape** | 1 | 1 | | | 7 | 252 | Numeric | Vapes: from hlthvape |
| 1016 | 7 | 0 | 1 | 0 | **Alcohol_Rate** | 1 | 1 | | | 7 | 15 | Numeric | Alcohol rate: from hlthalc |
| 1017 | 7 | 0 | 1 | 0 | **Alcohol_UnitsPerWeek** | 1 | 1 | | | 7 | 16 | Numeric | |
| 1018 | 7 | 0 | 1 | 0 | **PhysicalExertion_Work** | 1 | 1 | 5 | Physical exer | 7 | 210 | Numeric | Physical effort at work; from hlthgppaq1 |
| 1019 | 7 | 0 | 1 | 0 | **PhysicalExertion_Home** | 1 | 1 | | | 5 | 209 | Numeric | Sum of five activities; hlthgppaq2_1 to 5 |
| 1020 | 7 | 0 | 1 | 0 | **WalkingPace** | 1 | 1 | | | 7 | 253 | Numeric | Walking Pace: hlthgppaq3 |
| 1021 | 7 | 0 | 1 | 0 | **Lockdown_Unhealthy** | 1 | 1 | | | 7 | 186 | Numeric | Unhealthy behaviours in lockdown: hlthchange_1 to 4 |
| 1022 | 7 | 0 | 1 | 0 | **HealthCare_GP** | 1 | 1 | | | 7 | 145 | Numeric | Visits to GP: hlthvstgp |
| 1023 | 7 | 0 | 1 | 0 | **HealthCare_Inpatient** | 1 | 1 | | | 7 | 147 | Numeric | Days as inpatient in hospital: hlthinpt |
| 1024 | 7 | 0 | 1 | 0 | **hlthflujabM** | | | | | | | Numeric | |
| 1025 | 7 | 0 | 1 | 0 | **hlthflujab_2M** | | | | | | | Numeric | |
| 1026 | 7 | 0 | 1 | 0 | **FluVaccines_Ever** | | | | | | | Numeric | Flu jab ever -- including Yes on one measure and missing on the other, Chris Martin style |
| 1027 | 7 | 0 | 1 | 0 | **FluVaccines** | | | | | | | Numeric | N Flu vaccines this year and last year; from hlthflujab and _2 |
| 1028 | 7 | 0 | 1 | 0 | **FluVaccines_AtLeastOnce** | | | | | | | Numeric | |
| 1029 | 7 | 0 | 1 | 0 | **FluVaccines_BothYears** | | | | | | | Numeric | |
| 1030 | 7 | 0 | 1 | 0 | **FluVaccine_2019_20** | 1 | 1 | 2 | Had Flu Vacc | 2 | 126 | Numeric | |
| 1031 | 7 | 0 | 1 | 0 | **FluVaccine_2020_21** | | | | | | | Numeric | |
| 1032 | 7 | 0 | 1 | 0 | **HealthCare_Shielding** | 1 | 1 | | | 7 | 150 | Numeric | Health Care -Shielding: hlthshield |
| 1033 | 7 | 0 | 1 | 0 | **HealthCare_Ibuprofen** | 1 | 1 | | | 7 | 146 | Numeric | |
| 1034 | 7 | 0 | 1 | 0 | **HealthCare_VitaminD** | 1 | 1 | | | 7 | 151 | Numeric | |
| 1035 | 7 | 0 | 1 | 0 | **HealthCare_ACEinhibitor** | 1 | 1 | | | 7 | 144 | Numeric | |
| 1036 | 7 | 0 | 1 | 0 | **HealthCare_Sartan** | 1 | 1 | | | 7 | 149 | Numeric | |
| 1037 | 7 | 0 | 1 | 0 | **HealthCare_Entresto** | | | | | | | Numeric | |
| 1038 | 7 | 0 | 1 | 0 | **HealthCare_Metformin** | 1 | 1 | | | 7 | 148 | Numeric | |
| 1039 | 7 | 0 | 1 | 0 | **Comorbidities** | | | | | | | Numeric | Comorbidities -- Any reported |
| 1040 | 7 | 0 | 1 | 0 | **HealthComorbidity_Pregnant** | 1 | 1 | 2 | Pregnant | 2 | 166 | Numeric | |
| 1041 | 7 | 0 | 1 | 0 | **HealthComorbidity_Transplant** | 1 | 1 | | | 7 | 169 | Numeric | |
| 1042 | 7 | 0 | 1 | 0 | **HealthComorbidity_Diabetes** | 1 | 1 | | | 7 | 157 | Numeric | |
| 1043 | 7 | 0 | 1 | 0 | **HealthComorbidity_HeartDisease** | 1 | 1 | | | 7 | 158 | Numeric | |
| 1044 | 7 | 0 | 1 | 0 | **HealthComorbidity_Hypertension** | 1 | 1 | | | 7 | 159 | Numeric | |
| 1045 | 7 | 0 | 1 | 0 | **HealthComorbidity_Overweight** | 1 | 1 | | | 7 | 165 | Numeric | |
| 1046 | 7 | 0 | 1 | 0 | **HealthComorbidity_Stroke** | 1 | 1 | | | 7 | 168 | Numeric | |
| 1047 | 7 | 0 | 1 | 0 | **HealthComorbidity_KidneyDisease** | 1 | 1 | | | 7 | 161 | Numeric | |
| 1048 | 7 | 0 | 1 | 0 | **HealthComorbidity_LiverDisease** | 1 | 1 | | | 7 | 162 | Numeric | |
| 1049 | 7 | 0 | 1 | 0 | **HealthComorbidity_Anaemia** | 1 | 1 | | | 7 | 152 | Numeric | |
| 1050 | 7 | 0 | 1 | 0 | **HealthComorbidity_Asthma** | 1 | 1 | | | 7 | 154 | Numeric | |
| 1051 | 7 | 0 | 1 | 0 | **HealthComorbidity_LungDisease** | 1 | 1 | | | 7 | 163 | Numeric | |
| 1052 | 7 | 0 | 1 | 0 | **HealthComorbidity_Cancer** | 1 | 1 | | | 7 | 155 | Numeric | |
| 1053 | 7 | 0 | 1 | 0 | **HealthComorbidity_NeuralDisorder** | 1 | 1 | | | 7 | 164 | Numeric | |
| 1054 | 7 | 0 | 1 | 0 | **HealthComorbidity_ImmuneDisorder** | 1 | 1 | | | 7 | 160 | Numeric | |
| 1055 | 7 | 0 | 1 | 0 | **HealthComorbidity_Depression** | 1 | 1 | | | 7 | 156 | Numeric | |
| 1056 | 7 | 0 | 1 | 0 | **HealthComorbidity_Anxiety** | 1 | 1 | | | 7 | 153 | Numeric | |
| 1057 | 7 | 0 | 1 | 0 | **HealthComorbidity_PsychiatricDisorder** | 1 | 1 | | | 7 | 167 | Numeric | |
| 1058 | 7 | 0 | 1 | 0 | **HealthComorbidity_None** | | | | | | | Numeric | |
| 1059 | 7 | 0 | 1 | 0 | **ComorbiditySumExcPregnantOverweight** | | | | | | | Numeric | |
| 1060 | 7 | 0 | 1 | 0 | **ComorbiditySum2** | | 1 | | | 7 | 56 | Numeric | |
| 1061 | 7 | 0 | 1 | 0 | **ComorbiditySumPsych** | | 1 | | | 7 | 57 | | |
| 1061 | 7 | 0 | 1 | 0 | **Health_EQ5Dproblem_Mobility** | 1 | 1 | | | 7 | 136 | Numeric | |
| 1062 | 7 | 0 | 1 | 0 | **Health_EQ5Dproblem_SelfCare** | 1 | 1 | | | 7 | 138 | Numeric | |
| 1063 | 7 | 0 | 1 | 0 | **Health_EQ5Dproblem_UsualActivities** | 1 | 1 | | | 7 | 139 | Numeric | |
| 1064 | 7 | 0 | 1 | 0 | **Health_EQ5Dproblem_PainDiscomfort** | 1 | 1 | | | 7 | 137 | Numeric | |
| 1065 | 7 | 0 | 1 | 0 | **Health_EQ5Dproblem_AnxietyDepression** | 1 | 1 | | | 7 | 135 | Numeric | |
| 1066 | 7 | 0 | 1 | 0 | **Health_EQ5D_HealthGoodToday** | 1 | 1 | | | 7 | 134 | Numeric | EQ5D thermometer scale; 100 excellent Health 0 Worst Health |
| 1067 | 7 | 0 | 1 | 0 | **Health_EQ5Dproblem_Total** | | | | | | | Numeric | EQ5D -- simple sum of five main scales NB Not in manual |
| 1068 | 7 | 0 | 1 | 0 | **Health_AnxietyGAD2** | 1 | 1 | | | 7 | 132 | Numeric | Anxiety - sum of two GAD2 scores. High = anxiety |
| 1069 | 7 | 0 | 1 | 0 | **Health_DepressionPHQ2** | 1 | 1 | | | 7 | 133 | Numeric | Depression - sum of two PHQ2 scores. High = depression |
| 1070 | 7 | 0 | 1 | 0 | **Health_FinancialWorries** | 1 | 1 | 4 | Financial Cor | 4 | 140 | Numeric | Health Financial Worries: from hlthfiance |
| 1071 | 7 | 0 | 1 | 0 | **Health_PTSD** | 1 | 1 | 5 | PTSD | 5 | 143 | Numeric | Health PTSD: from healthptsd1/2/3 |
| 1072 | 7 | 0 | 1 | 0 | **Health_Loneliness** | 1 | 1 | | | 7 | 142 | Numeric | Health UCLA Loneliness scale: from hlthuclalonely_1/2/3 |
| 1073 | 7 | 0 | 1 | 0 | **Health_LifeSatisfaction** | 1 | 1 | 5 | Life Satisfact | 5 | 141 | Numeric | Health Life Satisfaction (ONS): from hlthsat |
| 1074 | 8 | 0 | 0 | 1 | **Section8_Covid______________Derived** | | | | | | | Numeric | |

| | | | | | | | | | | | | | |
|---|---|---|---|---|---|---|---|---|---|---|---|---|---|
| 1075 | 8 | 0 | 1 | 0 | Covid_Contact2wks | 1 | 1 | | | 7 | 64 | Numeric | Covid contact: from c19contract |
| 1076 | 8 | 0 | 1 | 0 | Covid_Prevention_JantoMar20 | 1 | 1 | | | 7 | 81 | Numeric | Covid Preventive measures Pre-Lockdown: c19_behav_1___1, etc |
| 1077 | 8 | 0 | 1 | 0 | Covid_Prevention_Now | 1 | 1 | 5 | Current COV | 5 | 82 | Numeric | Covid Preventive measures Now: c19_behav_1___2, etc |
| 1078 | 8 | 0 | 1 | 0 | Covid_LockdownEnjoyed | 1 | 1 | | | 7 | 75 | Numeric | Enjoyed aspects of lockdown: lockdownresponse |
| 1079 | 8 | 0 | 1 | 0 | c19typetest | | | | | | | Numeric | col 1 (10000s) = missing; col 2 PNTA; col3 Had Ab test; 4 Had Swab; 5 No tests; 2=ticked 1 = not ticked |
| 1080 | 8 | 0 | 1 | 0 | c19abresM | | | | | | | Numeric | |
| 1081 | 8 | 0 | 1 | 0 | c19swabresM | | | | | | | Numeric | |
| 1082 | 8 | 0 | 1 | 0 | Covid_AntibodyTest_ChrisMartin | 4 | | | | | | Numeric | |
| 1083 | 8 | 0 | 1 | 0 | Covid_SwabTest_ChrisMartin | 4 | | | | | | Numeric | |
| 1084 | 8 | 0 | 1 | 0 | Covid_AntibodyTest | | | | | | | Numeric | Ever had covid antibody test -- from Covid_AntibodyTest_ChrisMartin |
| 1085 | 8 | 0 | 1 | 0 | Covid_SwabTest | | | | | | | Numeric | Ever had covid swab test -- from Covid_SwabTest_ChrisMartin |
| 1086 | 8 | 0 | 1 | 0 | Covid_AntibodyPositive | 1 | 1 | | | 7 | 61 | Numeric | Positive antibody; from Covid_AntibodyTest_ChrisMartin |
| 1087 | 8 | 0 | 1 | 0 | Covid_SwabPositive | 1 | 1 | | | 7 | 85 | Numeric | Positive swab; from Covid_SwabTest_ChrisMartin |
| 1088 | 8 | 0 | 1 | 0 | Covid_SwabTest_ChrisMartinM | | | | | | | Numeric | |
| 1089 | 8 | 0 | 1 | 0 | Covid_AntibodyTest_ChrisMartinM | | | | | | | Numeric | |
| 1090 | 8 | 0 | 1 | 0 | Covid_Positive | | 1 | | | 7 | 80 | Numeric | |
| 1091 | 8 | 0 | 1 | 0 | Covid_Nsymptoms2weeks | 1 | 1 | | | 6 | 77 | Numeric | Covid N symptoms 2 weeks; from c19sympt___1 etc |
| 1092 | 8 | 0 | 1 | 0 | Covid_AnySymptoms2weeks | | | 6 | Symptoms in past two weeks | | | Numeric | Covid any symptoms 2 weeks; from c19sympt___1 etc |
| 1093 | 8 | 0 | 1 | 0 | Covid_ProbablyHad | 1 | 1 | | | 7 | 83 | Numeric | 1=suspected etc 0 = No, unsure; from c19ever |
| 1094 | 8 | 0 | 1 | 0 | CovidInfection_Date | | | | | | | Numeric | Date Covid Infection 1=Jan2020: c19date_year + month |
| 1095 | 8 | 0 | 1 | 0 | Covid_Hospitalised | 1 | 1 | | | 3 | 73 | Numeric | |
| 1096 | 8 | 0 | 1 | 0 | Covid_LongSymptoms | 1 | 1 | | | 7 | 76 | Numeric | Lasting symptoms; see c19_long & c19long_diag |
| 1097 | 8 | 0 | 1 | 0 | Covid_ConcernedUnknowinglySpread | 1 | 1 | 4 | Concerned al | 4 | 62 | Numeric | Concerned get covid; Quite or Very concerned; c19crnot |
| 1098 | 8 | 0 | 1 | 0 | Covid_KnowPeopleWhoHaveDied | 1 | 1 | | | 7 | 74 | Numeric | From c19died 1 to 4: Personally know anyone who has died from Covid |
| 1099 | 8 | 0 | 1 | 0 | Covid_Info_Nsources | | | | | | | Numeric | |
| 1100 | 8 | 0 | 1 | 0 | c19info1_friends | 1 | 1 | | | 7 | 30 | Numeric | |
| 1101 | 8 | 0 | 1 | 0 | c19info2_colleagues | 1 | 1 | | | 7 | 43 | Numeric | |
| 1102 | 8 | 0 | 1 | 0 | c19info3_employer | 1 | 1 | | | 7 | 44 | Numeric | |
| 1103 | 8 | 0 | 1 | 0 | c19info4_TV | 1 | 1 | | | 7 | 45 | Numeric | |
| 1104 | 8 | 0 | 1 | 0 | c19info5_radio | 1 | 1 | | | 7 | 46 | Numeric | |
| 1105 | 8 | 0 | 1 | 0 | c19info6_Newspapers | 1 | 1 | | | 7 | 47 | Numeric | |
| 1106 | 8 | 0 | 1 | 0 | c19info7_Govt_NHS | 1 | 1 | | | 7 | 48 | Numeric | |
| 1107 | 8 | 0 | 1 | 0 | c19info8_Twitter | 1 | 1 | | | 7 | 49 | Numeric | |
| 1108 | 8 | 0 | 1 | 0 | c19info9_SocialMedia | 1 | 1 | 3 | Info: Social M | 3 | 50 | Numeric | |
| 1109 | 8 | 0 | 1 | 0 | c19info10_UKGovt_website | 1 | 1 | | | 7 | 35 | Numeric | |
| 1110 | 8 | 0 | 1 | 0 | c19info11_Welsh_Scot_NI_website | 1 | 1 | | | 7 | 36 | Numeric | |
| 1111 | 8 | 0 | 1 | 0 | c19info12_NHSwebsite | 1 | 1 | | | 7 | 37 | Numeric | |
| 1112 | 8 | 0 | 1 | 0 | c19info13_WHOwebsite | 1 | 1 | | | 7 | 38 | Numeric | |
| 1113 | 8 | 0 | 1 | 0 | c19info14_OtherWebsites | 1 | 1 | | | 7 | 39 | Numeric | |
| 1114 | 8 | 0 | 1 | 0 | c19info15_LocalCouncil | 1 | 1 | | | 7 | 40 | Numeric | |
| 1115 | 8 | 0 | 1 | 0 | c19info16_GPetc | 1 | 1 | | | 7 | 41 | Numeric | |
| 1116 | 8 | 0 | 1 | 0 | c19info17_SciJournals | 1 | 1 | | | 7 | 42 | Numeric | |
| 1117 | 8 | 0 | 1 | 0 | Covid_Info_Official | | | | | | | Numeric | Info on c19 from official websites e.g. govt NHS |
| 1118 | 8 | 0 | 1 | 0 | Covid_Info_Friends | | | | | | | Numeric | Info on c19 from friends e.g. colleagues, social media |
| 1119 | 8 | 0 | 1 | 0 | Covid_Info_Media | | | | | | | Numeric | Info on c19 from media e.g. TV, radio, newspapers |
| 1120 | 8 | 0 | 1 | 0 | Covid_Info_Science | | | | | | | Numeric | Info on c19 from science sources e.g. journals, WHO, other websites |
| 1121 | 8 | 0 | 1 | 0 | c19info_Official | | 1 | | | 7 | 32 | Numeric | |
| 1122 | 8 | 0 | 1 | 0 | c19info_Friends | | 1 | | | 7 | 30 | Numeric | |
| 1123 | 8 | 0 | 1 | 0 | c19info_Media | | 1 | | | 7 | 31 | Numeric | |
| 1124 | 8 | 0 | 1 | 0 | c19info_Science | | 1 | | | 7 | 33 | Numeric | |
| 1125 | 8 | 0 | 1 | 0 | Covid_PerceivedRisks_averaged | 1 | 1 | 3 | Perceived ris | 3 | 79 | Numeric | Averaged c19 risks -- c19riskspermonth etc -- 0 optimistic 100 pessimistic |
| 1126 | 8 | 0 | 1 | 0 | Vaccine_Hesitant | 1 | 1 | 1 | Vaccine hesit | 1 | 250 | Numeric | |
| 1127 | 8 | 0 | 1 | 0 | Vaccine_Refusing | 1 | | | | | | Numeric | |
| 1128 | 8 | 0 | 1 | 0 | VaccQ_Jan | 1 | 1 | 2 | VaccQ Jan ve | 2 | 251 | Numeric | January (not Dec) version of vaccination questions |
| 1129 | 8 | 0 | 1 | 0 | Vaccinated1_Yes | | | | | | | Numeric | |
| 1130 | 8 | 0 | 1 | 0 | Vaccinated2_Vaccinated_NearFuture | | | | | | | Numeric | |
| 1131 | 8 | 0 | 1 | 0 | Vaccinated3_OfferedButRefused | | | | | | | Numeric | |
| 1132 | 8 | 0 | 1 | 0 | Vaccinated4_IntendToHaveVaccine | | | | | | | Numeric | |
| 1133 | 8 | 0 | 1 | 0 | Vaccinated5_NoOfferButWillRefuse | | | | | | | Numeric | |
| 1134 | 8 | 0 | 1 | 0 | VaccHes1_NeedlesEtc | | | | | | | Numeric | |
| 1135 | 8 | 0 | 1 | 0 | VaccHes2_SideEffects | | | | | | | Numeric | |
| 1136 | 8 | 0 | 1 | 0 | VaccHes3_NotEffective | | | | | | | Numeric | |
| 1137 | 8 | 0 | 1 | 0 | VaccHes4_EthnicTesting | | | | | | | Numeric | |
| 1138 | 8 | 0 | 1 | 0 | VaccHes5_HadCovid | | | | | | | Numeric | |

| | | | | | | | | | | | | | |
|---|---|---|---|---|---|---|---|---|---|---|---|---|---|
| 1139 | 8 | 0 | 1 | 0 | VaccHes6_InTrial | | | | | | | Numeric | |
| 1140 | 8 | 0 | 1 | 0 | VaccHes7_WantDiffVaccine | | | | | | | Numeric | |
| 1141 | 8 | 0 | 1 | 0 | VaccHes8_WaitForOthers | | | | | | | Numeric | |
| 1142 | 8 | 0 | 1 | 0 | VaccHes9_NotAtRisk | | | | | | | Numeric | |
| 1143 | 8 | 0 | 1 | 0 | VaccHes10_UsedForOthers | | | | | | | Numeric | |
| 1144 | 8 | 0 | 1 | 0 | VaccHes11_DontBelieveInVaccines | | | | | | | Numeric | |
| 1145 | 8 | 0 | 1 | 0 | VaccHes12_OtherReason | | | | | | | Numeric | |
| 1146 | 8 | 0 | 1 | 0 | VaccHes_Nreasons | | | | | | | Numeric | N reasons for vaccine hesitancy -- only if questionnaire complete |
| 1147 | 8 | 0 | 1 | 0 | Covid_Provaccine | 1 | 1 | 2 | VAX scale: Pr | 2 | 84 | Numeric | VAXX sub-scales combined: rely-safe-gain-nat ; 20 high provaccination 4 low antivaccination |
| 1148 | 8 | 0 | 1 | 0 | Covid_HappyToBeInTrial | 1 | | | | | | Numeric | Either in trial (3%) or willing to be in trial (29%) |
| 1149 | 8 | 0 | 1 | 0 | Covid_OptimisticAboutPopnVaccination | 1 | 1 | 2 | Optimism ab | 2 | 78 | Numeric | How soon popn vaccinated 8 = soon (optimistic) 1 = A long while (pessimistic); from c19vacchap |
| 1150 | 8 | 0 | 1 | 0 | Covid_Effects1_ChildrenAndEducation | 1 | 1 | | | 7 | 65 | Numeric | C19 effects on children and education |
| 1151 | 8 | 0 | 1 | 0 | Covid_Effects2_Economy | 1 | 1 | | | 7 | 66 | Numeric | C19 effects on economy |
| 1152 | 8 | 0 | 1 | 0 | Covid_Effects3_DeathsFromC19 | 1 | 1 | | | 7 | 67 | Numeric | C19 effects of C19 deaths |
| 1153 | 8 | 0 | 1 | 0 | Covid_Effects4_IndirectDeaths | 1 | 1 | | | 7 | 68 | Numeric | C19 indirect deaths |
| 1154 | 8 | 0 | 1 | 0 | Covid_Effects5_MentalHealthIssues | 1 | 1 | 6 | Concerns on | 6 | 69 | Numeric | C19 effects on Mental Health |
| 1155 | 8 | 0 | 1 | 0 | Covid_Conspir_Knowledge_Nanswers | | | | | | | Numeric | |
| 1156 | 8 | 0 | 1 | 0 | Covid_Conspiracies | 1 | 1 | 2 | Covid-19 Cor | 2 | 63 | Numeric | Total score on six conspiracy items; high = conspiratorial beliefs |
| 1157 | 8 | 0 | 1 | 0 | Covid_Knowledge | 1 | 1 | 3 | Covid Knowle | 2 | 72 | Numeric | Total score on five knowledge items: High = Good knowledge |
| 1158 | 8 | 0 | 1 | 0 | Doctor | | | | | | | Numeric | Is a doctor -- GMC or self-classified on jobrole |
| 1159 | 8 | 0 | 1 | 0 | Doctor_Source | | | | | | | Numeric | Source of Doctor -- 1 GMC 2 Jobrole |
| 1160 | 8 | 0 | 1 | 0 | nurse | | | | | | | Numeric | Is a nurse -- GMC or self-classified on jobrole |
| 1161 | 8 | 0 | 1 | 0 | nurse_Source | | | | | | | Numeric | Source of nurse -- 1 GMC 2 Jobrole |
| 1162 | 9 | 0 | 0 | 1 | Section9_PersonalityEtc_____________Derived | | | | | | | Numeric | |
| 1163 | 9 | 0 | 1 | 0 | Big5_Extraversion | 1 | 1 | 5 | Big 5: Extrave | 5 | 20 | Numeric | |
| 1164 | 9 | 0 | 1 | 0 | Big5_Neuroticism | 1 | 1 | 5 | Big 5: Neurot | 5 | 21 | Numeric | |
| 1165 | 9 | 0 | 1 | 0 | Big5_Openness | 1 | 1 | | | 5 | 22 | Numeric | |
| 1166 | 9 | 0 | 1 | 0 | Big5_Agreeableness | 1 | 1 | 5 | Big 5: Agreea | 6 | 18 | Numeric | |
| 1167 | 9 | 0 | 1 | 0 | Big5_Conscientiousness | 1 | 1 | 5 | Big 5: Consci | 5 | 19 | Numeric | |
| 1168 | 9 | 0 | 1 | 0 | LocusOfControl_Internal | 1 | 1 | | | 7 | 189 | Numeric | Internal locus of control |
| 1169 | 9 | 0 | 1 | 0 | LocusOfControl_Chance | 1 | 1 | 4 | LoC: Chance | 4 | 187 | Numeric | Chance locus of control |
| 1170 | 9 | 0 | 1 | 0 | LocusOfControl_External | 1 | 1 | 4 | LoC: External | 4 | 188 | Numeric | External locus of control |
| 1171 | 9 | 0 | 1 | 0 | Fatalism | 1 | 1 | 3 | Fatalism | 3 | 125 | Numeric | How do you see your own future changing as a result of COVID-19? |
| 1172 | 9 | 0 | 1 | 0 | NeedForCognitiveClosure | 1 | 1 | 4 | Need for Clo: | 4 | 206 | Numeric | From Pagnini et al JAMA Open 2020 -- sum of two items |
| 1173 | 9 | 0 | 1 | 0 | RiskTaking | 1 | 1 | | | 7 | 230 | Numeric | 10 high risk taker; 0 low risk taker; from lifegnrisk |
| 1174 | 9 | 0 | 1 | 0 | Burnout_Past | 1 | 1 | | | 7 | 29 | Numeric | Mean Burnout 2019 Copenhagen GENHQ202 + GENHQ204 Never=1 Every Day=7 |
| 1175 | 9 | 0 | 1 | 0 | WorkEngagement_Past_Absorption | 1 | 1 | | | 7 | 258 | Numeric | Absorption - Work Engagement - Past = workengbout2 7 = Every Day 1=Never |
| 1176 | 9 | 0 | 1 | 0 | WorkEngagement_Past_Dedication | 1 | 1 | | | 7 | 259 | Numeric | Dedication - Work Engagement - Past = workengbout4 7 = Every Day 1=Never |
| 1177 | 9 | 0 | 1 | 0 | WorkEngagement_Past_Vigor | 1 | 1 | | | 7 | 260 | Numeric | Vigor - Work Engagement - Past = workengbout5 7 = Every Day 1=Never |
| 1178 | 9 | 0 | 1 | 0 | WorkClim_ChoiceInd_Past | 1 | 1 | | | 7 | 255 | Numeric | Workplace Climate - Past - Choice&Independence -- High=agree |
| 1179 | 9 | 0 | 1 | 0 | WorkClim_Workload_Past | 1 | 1 | | | 7 | 257 | Numeric | Workplace Climate - Past - Workload -- High score= high workload |
| 1180 | 9 | 0 | 1 | 0 | WorkClim_Supportive_Past | 1 | 1 | | | 7 | 256 | Numeric | Workplace Climate - Past - Supportive-Receptive - High=supportive |
| 1181 | 10 | 0 | 0 | 1 | Section10_FinalQuestions_____________Derived | | | | | | | Numeric | |
| 1182 | 10 | 0 | 1 | 0 | exbamec19_Length | | | | | | | Numeric | Open-ended text on BAME risk -- number of words written |
| 1183 | 10 | 0 | 1 | 0 | exbamec19_Wrote | 1 | 1 | | | 7 | 122 | Numeric | Open-ended text on BAME risk: something written |
| 1184 | 10 | 0 | 1 | 0 | exsocc19_Length | | | | | | | Numeric | Open-ended text on BAME C19 & society -- number of words written |
| 1185 | 10 | 0 | 1 | 0 | exsocc19_Wrote | 1 | 1 | | | 7 | 123 | Numeric | Open-ended text on c19 & society: something written |
| 1186 | 10 | 0 | 1 | 0 | exyouc19_Length | | | | | | | Numeric | How do you see your own future changing as a result of COVID-19? |
| 1187 | 10 | 0 | 1 | 0 | exyouc19_Wrote | 1 | 1 | | | 7 | 124 | Numeric | How do you see your own future changing as a result of COVID-19? |
| 1188 | 10 | 0 | 1 | 0 | Q_TooLong | | | | | | | Numeric | |
| 1189 | 10 | 0 | 1 | 0 | Q_Useful | | | | | | | Numeric | |
| 1190 | 10 | 0 | 1 | 0 | Questionnaire_Useful | 1 | 1 | 5 | Questionnair | 7 | 217 | Numeric | Questionnaire useful 5 = extremely 1 = not at all |
| 1191 | 10 | 0 | 1 | 0 | Questionnaire_TooLong | 1 | 1 | | | 7 | 216 | Numeric | Questionnaire length 5 = Much shorter 1 = asked more things |
| 1192 | 10 | 0 | 0 | 1 | SectionEndfDerivedVariables_____________Derived | | | | | | | Numeric | |
| 1193 | 10 | 0 | 0 | 1 | VariablesAwaitingProperPlacement_____________Derived | | | | | | | Numeric | |
